# Phenotype-guided prophylactic vasopressor selection for post-induction hypotension in older adults: a systematic review and network meta-analysis

**DOI:** 10.64898/2026.06.15.26355638

**Authors:** ZiWen Zhang, Dongxu Wang, Chun Ling Duan, XuanYi Di, Yan Ran Wang, HongHai Zhang

## Abstract

**Background:** Post-induction hypotension is a predictable haemodynamic hazard in older adults undergoing general anaesthesia. Prevention remains divided among volume optimisation, anaesthetic dose reduction, rescue treatment after hypotension occurs and proactive vasoactive support.

**Methods:** We searched PubMed, Embase, Web of Science, CENTRAL, CNKI, Wanfang and VIP from inception to 30 March 2026. Eligible studies were randomised trials of prophylactic vasoactive drugs given before, during or immediately after induction in older adults. The primary outcome was post-induction hypotension. Secondary outcomes were post-induction mean arterial pressure (MAP), systolic arterial pressure (SBP), heart rate (HR) and reported haemodynamic adverse events. Random-effects network meta-analysis was used, and confidence in network estimates was assessed using CINeMA principles.

**Results:** Thirty-one trials including 2,821 participants were included in the revised network. Compared with placebo/control, all active agents favoured lower post-induction hypotension. The most favourable point estimates were observed for phenylephrine (odds ratio [OR] 0.17, 95% confidence interval [CI] 0.01 to 2.16) and metaraminol (OR 0.19, 95% CI 0.02 to 1.53), although both were imprecise. More precise reductions were observed for methoxamine (OR 0.23, 95% CI 0.13 to 0.43), norepinephrine (OR 0.25, 95% CI 0.13 to 0.47) and ephedrine (OR 0.34, 95% CI 0.19 to 0.63). Phenylephrine ranked highest for MAP support, norepinephrine ranked highest for SBP support, and ephedrine ranked highest for HR preservation. Global inconsistency was detected for SBP but not for hypotension incidence, MAP or HR, supporting cautious profile-based interpretation.

**Conclusions:** Prophylactic vasopressor choice during induction should be guided by haemodynamic phenotype rather than ranking alone. In the revised network, active prophylaxis consistently favoured lower hypotension, but sparse nodes produced uncertainty. Norepinephrine retained a comparatively balanced profile when vasodilatory post-induction hypotension is anticipated, phenylephrine and related alpha-agonists provided stronger pressure support when HR and cardiac-output reserve are preserved, and ephedrine was most relevant when chronotropic support is desired.

**Key points:**

- This network meta-analysis compares prophylactic vasoactive agents for preventing post-induction hypotension in older adults undergoing general anaesthesia.
- Across 31 randomised trials, active prophylaxis generally reduced hypotension in the revised network, but agents separated into different haemodynamic profiles: alpha-agonists for pressure support, norepinephrine for a comparatively balanced pressure-and-heart-rate profile, and ephedrine for heart-rate preservation

## Introduction

Surgery is a major exposure for ageing health systems. An estimated 312.9 million operations were performed worldwide in 2012, up from 226.4 million in 2004 ^[1]^. Older adults account for an increasing share of this burden, with English projections showing rapid growth among patients aged ≥75 yr ^[2]^. These patients frequently carry frailty, multimorbidity and reduced physiological reserve. In SNAP-3, frailty was associated with higher postoperative morbidity, delirium, length of stay and mortality ^[3]^. Frailty is therefore not chronological age alone, but a multidimensional loss of resilience linked to postoperative complications and death ^[4,5]^.

Induction of general anaesthesia is a short period in which this vulnerability becomes clinically visible. Before surgical stimulation begins, propofol- and opioid-based induction and positive-pressure ventilation can reduce sympathetic tone, venous return, mean systemic filling pressure and systemic vascular resistance ^[6-12]^. Effects on cardiac output vary. In older patients, vascular ageing, arterial stiffness, frailty, altered drug sensitivity, impaired autonomic compensation and reduced baroreflex reserve may limit rapid pressure recovery^[6,13-21]^. Intraoperative hypotension is common but definition-dependent ^[22]^, and greater depth and duration are associated with acute kidney injury, myocardial injury and mortality ^[23-28]^. Postoperative troponin elevation and myocardial injury after noncardiac surgery strongly predict 30-day mortality^[29,30]^. POQI linked mean arterial pressures below approximately 60-70 mm Hg with myocardial injury, acute kidney injury and death, while its 2024 consensus emphasised individualised pressure management ^[31,32]^.

The unresolved clinical question is not whether post-induction hypotension matters, but which mechanism should be prevented. Current practice variably emphasises hypnotic-dose reduction, preload assessment, fluid optimisation, rescue vasopressors after hypotension develops or prophylactic vasopressors before arterial pressure falls ^[17,33,34]^. This variation reflects three competing targets: preload, anaesthetic dose and vascular tone. No single target is likely to dominate in every older patient. Fluid therapy is logical when preload deficit predominates, and ultrasound-guided fluid administration may reduce induction hypotension in preload-responsive patients ^[34]^. Yet fluid alone may not correct the vasodilatory and sympatholytic physiology of propofol-opioid induction ^[7-12,33]^.

Prophylactic vasoactive drugs offer a biologically plausible prevention strategy, but useful selection depends on phenotype. The relevant phenotype includes anticipated vasodilatation, HR reserve, cardiac-output dependence and susceptibility to chronotropic or afterload-related harm. Randomised trials have evaluated alpha-adrenergic agents, mixed or indirect sympathomimetics and catecholamine-based strategies during induction, but the evidence remains fragmented. Many trials are small, most compare one active strategy with saline or placebo, and active-agent comparisons are limited. Clinicians therefore lack a comparative hierarchy that also respects mechanism. Network meta-analysis can integrate direct and indirect randomised evidence across interventions that have not all been compared head to head ^[35]^. We conducted this systematic review and network meta-analysis to compare prophylactic vasoactive agents for preventing post-induction hypotension in older adults undergoing general anaesthesia and to characterise haemodynamic profiles for phenotype-guided selection.

## Methods

### Protocol and reporting

This systematic review and network meta-analysis was designed according to PRISMA 2020 and PRISMA-NMA guidance.[36,37] The protocol was registered in PROSPERO (CRD420261355544). Because only aggregate data from published randomised trials were used, ethics approval and patient consent were not required.

### Eligibility criteria

Eligible studies were randomised controlled trials enrolling older adults undergoing induction of general anaesthesia. Older-adult eligibility was defined a priori as trials explicitly enrolling older, elderly or higher-age surgical patients, or as age-restricted cohorts with a mean or median age of ≥60 yr. Unselected adult trials were eligible only when older-adult subgroup data were extractable. Interventions were prophylactic norepinephrine, phenylephrine, ephedrine, metaraminol or methoxamine, given before, during or immediately after induction. Comparators were placebo, saline, no active prophylactic vasopressor or another eligible agent. We excluded non-randomised studies, non-general-anaesthesia settings, rescue-only treatment, dopamine-only prophylaxis, combined vasoactive regimens not attributable to a single eligible network node and studies without extractable induction-period outcomes.

### Information sources and search strategy

We searched PubMed, Embase, Web of Science, the Cochrane Central Register of Controlled Trials, CNKI, Wanfang and VIP from inception to 30 March 2026. Search terms combined concepts for older adults, general anaesthesia, induction, hypotension, prophylaxis, vasoactive or vasoconstrictor agents and individual drug names. Reference lists of included trials and relevant reviews were screened manually. Full search strategies are provided in Supplementary Appendix 1.

### Study selection and data extraction

Two reviewers independently screened titles, abstracts and potentially eligible full texts. Disagreements were resolved by consensus or adjudication by a third reviewer. Using a standardised form, two reviewers independently extracted study identifier, participant age, sample size, body mass index, baseline mean arterial pressure, randomised treatments, vasopressor agent, dose, route, timing, comparator, hypotension definition, hypotension incidence, post-induction mean arterial pressure and bradycardia. Multiple dose arms of the same agent were extracted separately and assigned to the same pharmacological node when clinically appropriate. Multi-arm trials were handled as such in the network analysis.

### Outcomes

The primary outcome was incidence of post-induction hypotension, using each trial’s original definition. This pragmatic approach preserved randomised comparisons but introduced definitional heterogeneity. Secondary outcomes were post-induction MAP, SBP and HR. For continuous outcomes, we used the post-induction time point closest to completion of induction or the earliest clinically relevant post-induction measurement.

### Risk of bias and certainty of evidence

Risk of bias was assessed independently by two reviewers using RoB 2, covering randomisation, deviations from intended interventions, missing outcome data, outcome measurement and selection of the reported result.[38] Disagreements were resolved by consensus. Confidence in network estimates was assessed using CINeMA principles, considering within-study bias, reporting bias, indirectness, imprecision, heterogeneity and incoherence^[39,40]^.

### Data synthesis

Network meta-analysis was performed for outcomes with a connected treatment network. Dichotomous outcomes were summarised as odds ratios (ORs) with 95% confidence intervals (CIs). Continuous outcomes were summarised as mean differences (MDs) with 95% CIs. For hypotension incidence, ORs less than 1 favoured the active prophylactic vasopressor over placebo. For MAP and SBP, positive MDs indicated higher post-induction pressure. For HR, positive MDs indicated higher post-induction HR. Random-effects models were used because clinical heterogeneity was expected across age thresholds, comorbidity, induction regimen, surgical setting, hypotension definition, vasopressor dose, route and timing. Network plots displayed geometry, with node size proportional to participants and edge width proportional to direct comparisons ^[41]^. Treatment ranking was summarised using SUCRA values, but rankings were interpreted alongside effect estimates, uncertainty, inconsistency and certainty of evidence ^[42]^.

### Inconsistency, heterogeneity and subgroup handling

Global inconsistency was assessed using a design-by-treatment interaction model or an equivalent global test. Local inconsistency was assessed using node-splitting comparisons between direct and indirect evidence, as implemented by Stata network sidesplit. Between-study heterogeneity was addressed with random-effects models and interpreted in relation to clinical and methodological diversity across trials. We prespecified that formal network subgroup analysis or meta-regression would be reported only when the subgroup formed a sufficiently connected network and did not leave treatment contrasts informed by single studies only. This criterion was important for induction hypnotic subgrouping. Propofol-predominant studies represented most evidence, whereas etomidate-predominant and mixed-regimen studies were sparse and unevenly distributed across drug nodes. We therefore used hypnotic regimen primarily for transitivity assessment and interpretation, and did not present a definitive separate hierarchy for etomidate induction.

## Results

### Study selection

The search identified 1,676 records. After duplicate removal, 1,130 records were screened by title and abstract, and 1,021 were excluded as clearly ineligible. Full texts were assessed for 109 reports; 78 were excluded in the revised eligibility process. Common reasons were PICOS mismatch, ineligible age group, combined vasoactive interventions, absence of an eligible comparator, dopamine-only prophylaxis, missing outcome data and non-randomised design. Thirty-one randomised controlled trials involving 2,821 participants were included. The study-selection process is shown in Figure 1.

**Figure 1.**
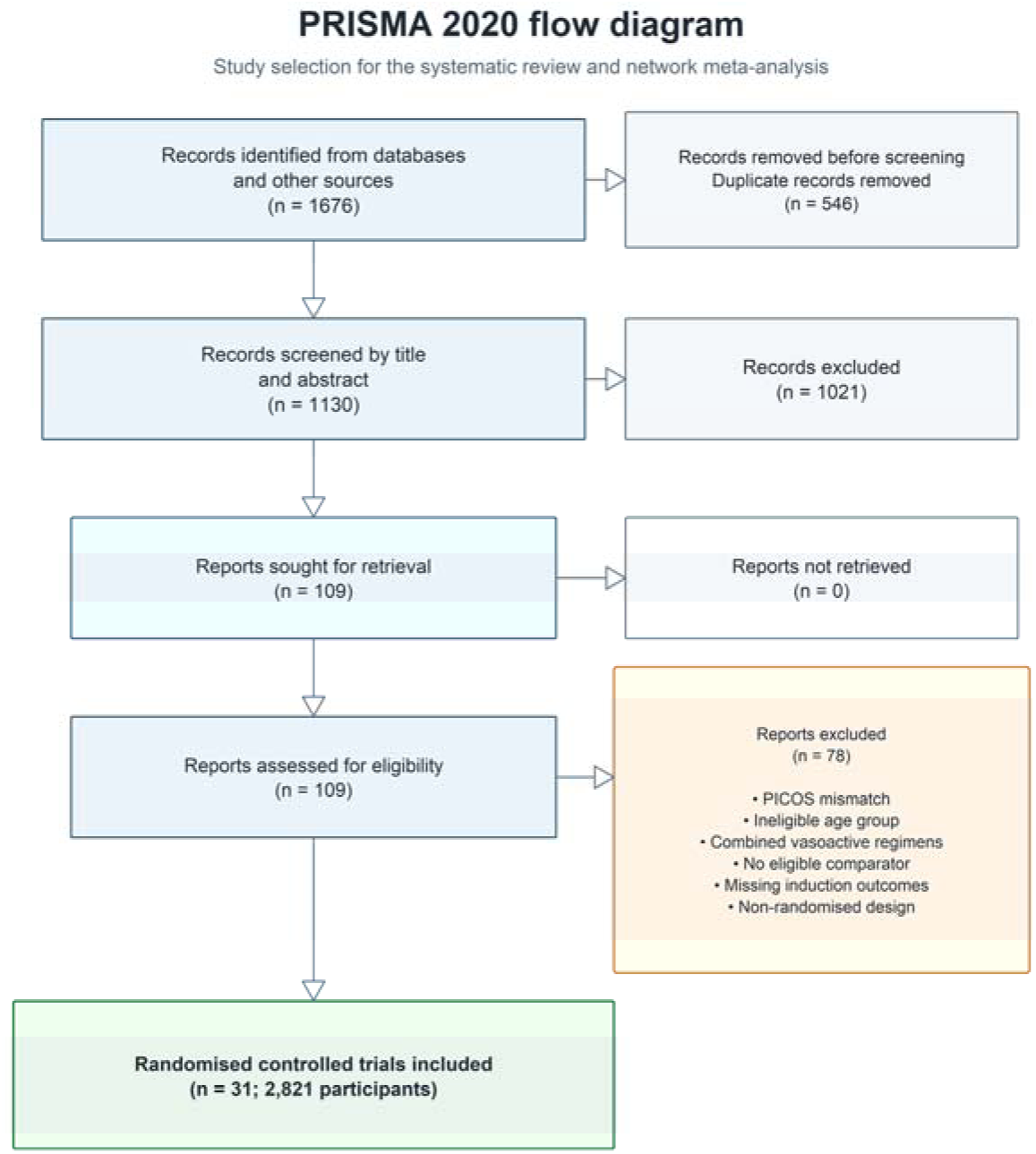
Study selection. PRISMA 2020 flow diagram showing record identification, duplicate removal, screening, full-text assessment and inclusion for the revised no-dopamine network meta-analysis.

### Study characteristics and network geometry

The included trials evaluated five active agents in the revised network: norepinephrine, phenylephrine, ephedrine, metaraminol and methoxamine. Placebo, saline and no active prophylactic vasopressor were grouped as the common inactive-control node. Methoxamine, ephedrine and norepinephrine were the most frequently represented active nodes; phenylephrine and metaraminol were supported by fewer trials and smaller participant numbers. Most trials used propofol-based or propofol-containing induction, usually with an opioid and, in surgical trials, a neuromuscular blocking drug. Fewer trials used etomidate-predominant induction or propofol-etomidate combinations. The overall network therefore estimates drug effects most directly in propofol-based or propofol-containing induction. Etomidate-predominant trials informed transitivity but did not support a stable independent hierarchy across active agents. Detailed treatment-arm characteristics, including age, randomised treatment, sample size, administration route, dose or infusion rate, BMI and baseline MAP, are provided in Supplementary Table S2.

#### Risk of bias

Risk of bias varied across trials. Missing outcome data and outcome measurement were generally less problematic than incomplete reporting of allocation concealment, blinding and deviations from intended interventions. Several Chinese-language trials reported random-number-table allocation but provided limited information on concealment, blinding or prespecified outcomes. This reporting pattern reduced confidence for some treatment nodes, particularly methoxamine and smaller alpha-agonist nodes, despite their contribution to network connectivity. Full study-level assessments are provided in Supplementary Appendix 4. The domain-level risk-of-bias summary is shown in Figure 2.

**Figure 2.**
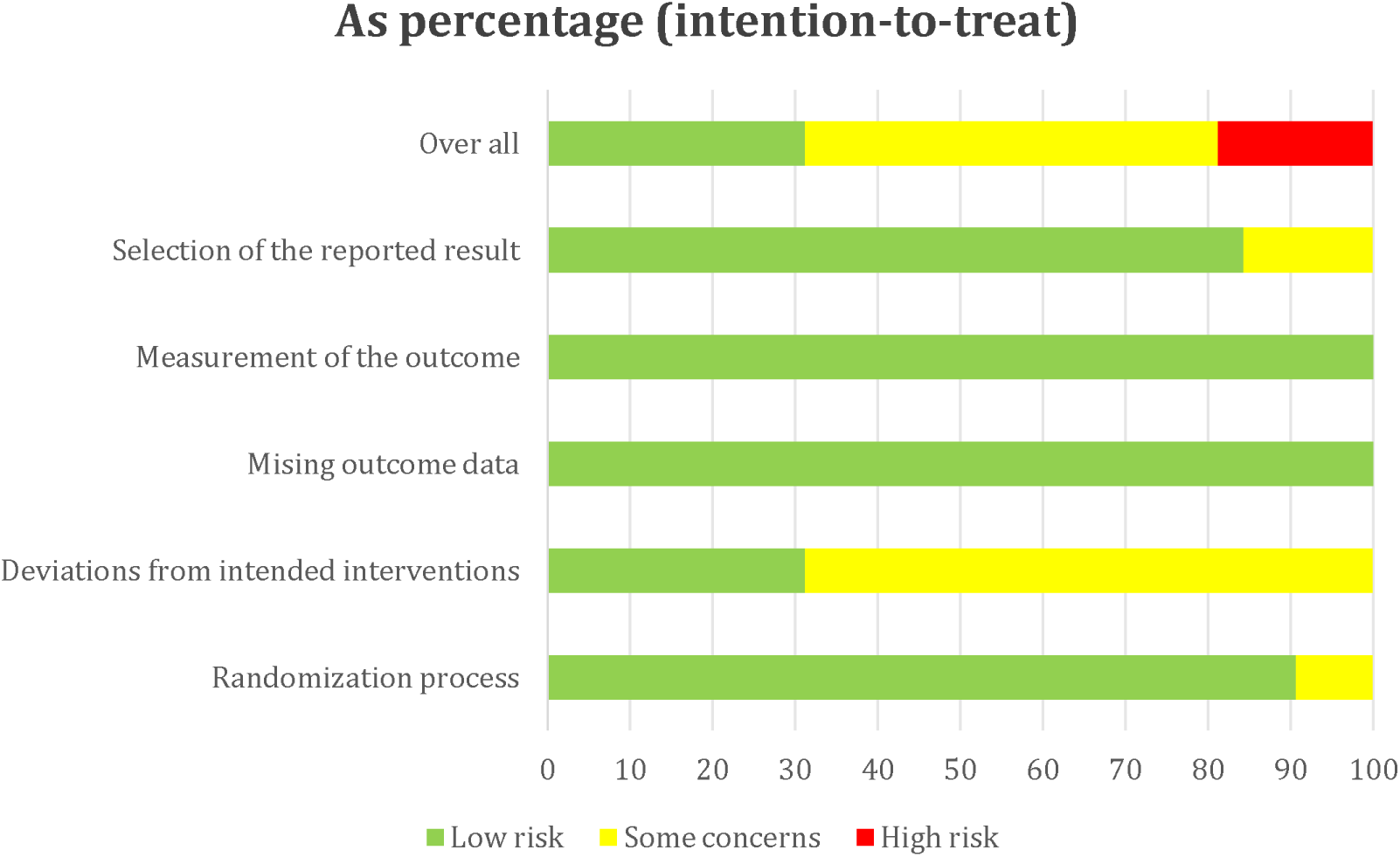
Risk of bias across included randomised controlled trials. Domain-level percentages were calculated from the study-level RoB 2 assessments in Supplementary Appendix 4. Green indicates low risk of bias, yellow indicates some concerns, and red indicates high risk of bias.

#### Incidence of post-induction hypotension

Compared with the inactive-control node, all estimable active agents favoured lower post-induction hypotension in the revised network. Phenylephrine had the most favourable point estimate (OR 0.17, 95% CI 0.01 to 2.16), followed by metaraminol (OR 0.19, 95% CI 0.02 to 1.53), methoxamine (OR 0.23, 95% CI 0.13 to 0.43), norepinephrine (OR 0.25, 95% CI 0.13 to 0.47) and ephedrine (OR 0.34, 95% CI 0.19 to 0.63). The phenylephrine and metaraminol estimates were directionally favourable but imprecise, whereas methoxamine, norepinephrine and ephedrine had CIs excluding no effect. Favourable SUCRA values for hypotension prevention were 66.8 for phenylephrine, 65.2 for metaraminol, 64.1 for methoxamine, 61.6 for norepinephrine, 39.4 for ephedrine and 2.9 for placebo/control. Integrated primary-outcome forest plots are shown in Figure 3. Across drug subgroups, most study-level estimates favoured active prophylaxis over placebo/control, although precision varied according to the number and size of contributing trials.

**Figure 3.**
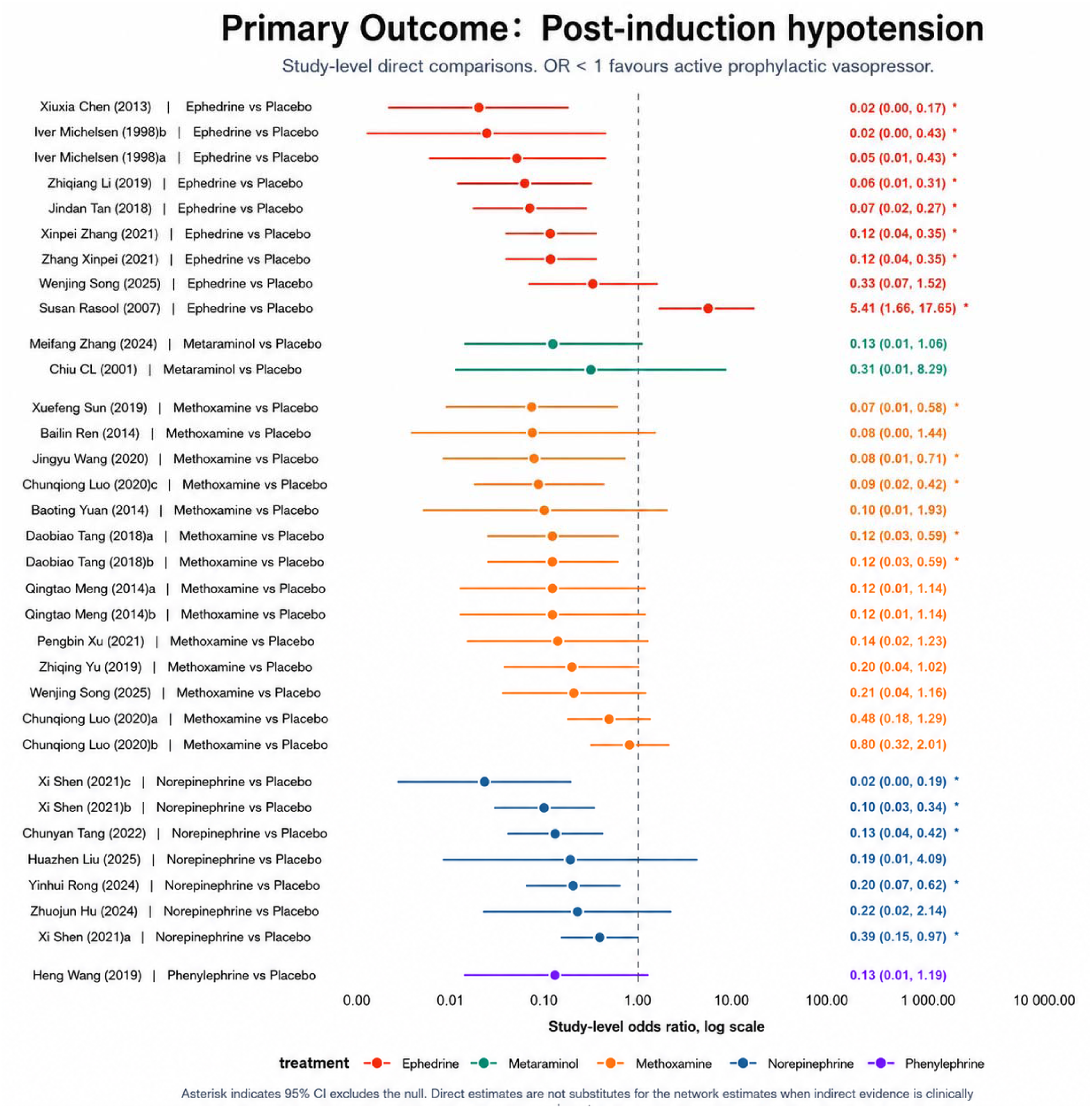
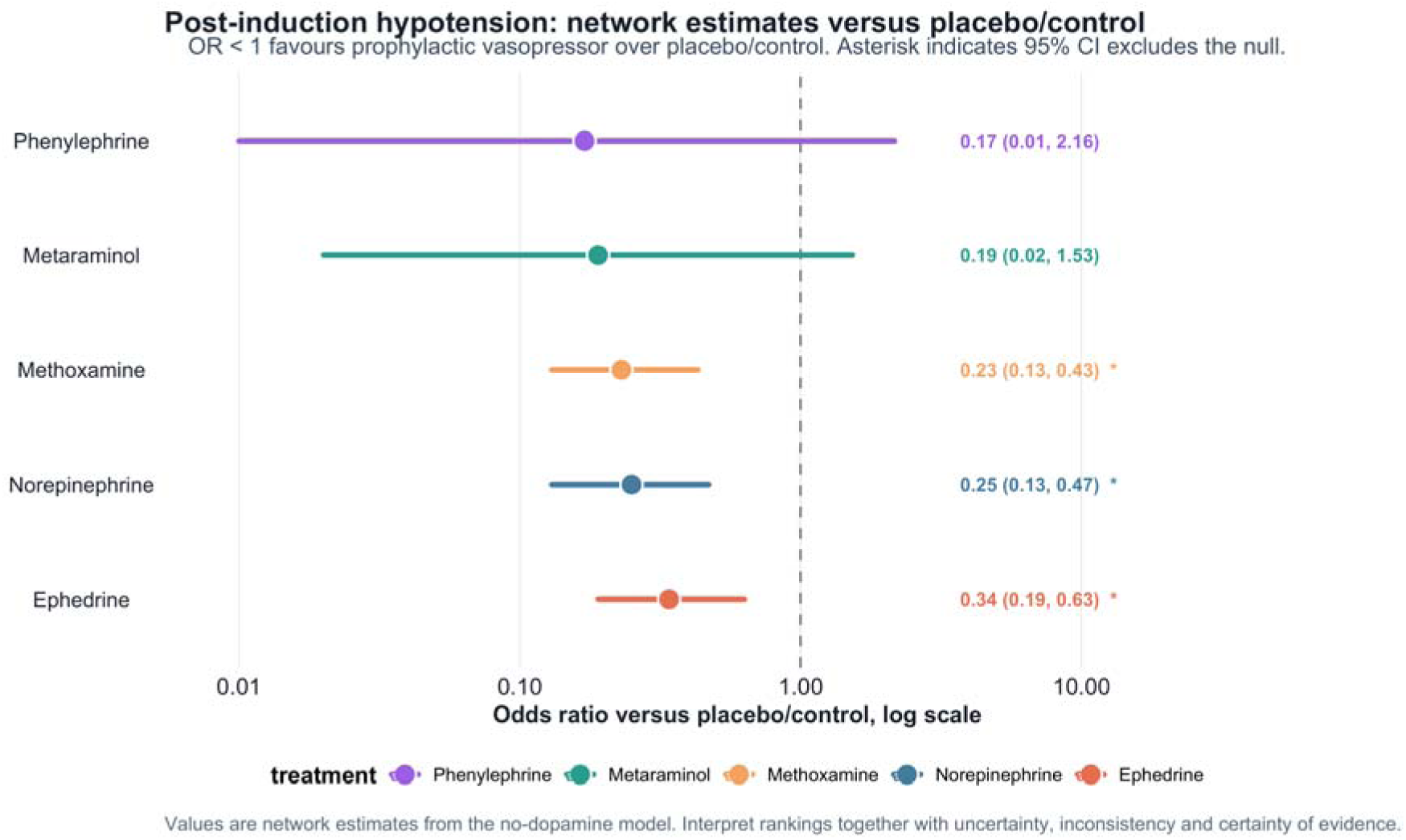
Primary-outcome forest plots for post-induction hypotension. (A) Study-level direct pairwise odds ratios grouped by prophylactic vasopressor. (B) Random-effects network estimates versus placebo/control from the revised no-dopamine network. Odds ratios less than 1 favour active prophylactic vasopressor. Points represent effect estimates and horizontal lines represent 95% confidence intervals; asterisks indicate 95% confidence intervals excluding the null. CI, confidence interval; OR, odds ratio.

#### Post-induction mean arterial pressure

The post-induction MAP network included placebo/control, ephedrine, metaraminol, methoxamine, norepinephrine and phenylephrine. Compared with placebo/control, phenylephrine produced the largest estimated MAP increase (MD 18.90 mm Hg, 95% CI 4.15 to 33.65). Methoxamine (MD 15.13 mm Hg, 95% CI 10.42 to 19.85), norepinephrine (MD 14.56 mm Hg, 95% CI 5.67 to 23.45) and ephedrine (MD 13.43 mm Hg, 95% CI 8.20 to 18.65) also increased MAP. Metaraminol favoured higher MAP but had a less precise estimate (MD 9.45 mm Hg, 95% CI -1.32 to 20.23). MAP rankings placed phenylephrine first (SUCRA 79.4; probability of being ranked best 57.5%). This pressure ranking should be interpreted alongside HR and flow-related considerations, rather than as evidence of superior organ perfusion.

#### Post-induction systolic arterial pressure

For post-induction SBP, phenylephrine was not estimable in the revised SBP network because extractable SBP data did not connect this node. Norepinephrine produced the largest estimated SBP increase compared with placebo/control (MD 18.30 mm Hg, 95% CI 8.60 to 28.00), followed by ephedrine (MD 16.97 mm Hg, 95% CI 8.62 to 25.31), metaraminol (MD 11.93 mm Hg, 95% CI -3.13 to 26.99) and methoxamine (MD 7.49 mm Hg, 95% CI 0.91 to 14.07). The SBP ranking placed norepinephrine first (SUCRA 82.1; probability of being ranked best 48.7%), closely followed by ephedrine (SUCRA 77.8; probability of being ranked best 34.6%).

#### Post-induction heart rate

Ephedrine was associated with the highest post-induction HR compared with placebo/control (MD 10.01 beats min-1, 95% CI 6.98 to 13.04). Norepinephrine was near neutral (MD 0.24 beats min-1, 95% CI -3.22 to 3.80), as was metaraminol (MD 0.65 beats min-1, 95% CI -5.90 to 7.20). Methoxamine reduced HR (MD -3.00 beats min-1, 95% CI -5.35 to -0.58), whereas phenylephrine showed a lower-HR tendency consistent with alpha-agonism (MD -7.30 beats min-1, 95% CI -17.30 to 2.66). The HR ranking placed ephedrine first (SUCRA 99.9; probability of being ranked best 99.4%). This finding separates pressure-raising efficacy from chronotropic profile in older adults. The combined secondary-outcome forest plots show a clear separation between pressure-support and chronotropic profiles: phenylephrine produced the largest MAP increase, norepinephrine produced the largest estimable SBP increase, and ephedrine produced the clearest HR-preserving effect (Figure 4). The SUCRA profile showed an outcome-specific ranking pattern: phenylephrine ranked highest for MAP support, norepinephrine ranked highest for SBP support, and ephedrine ranked highest for HR preservation (Figure 5).

**Figure 4.**
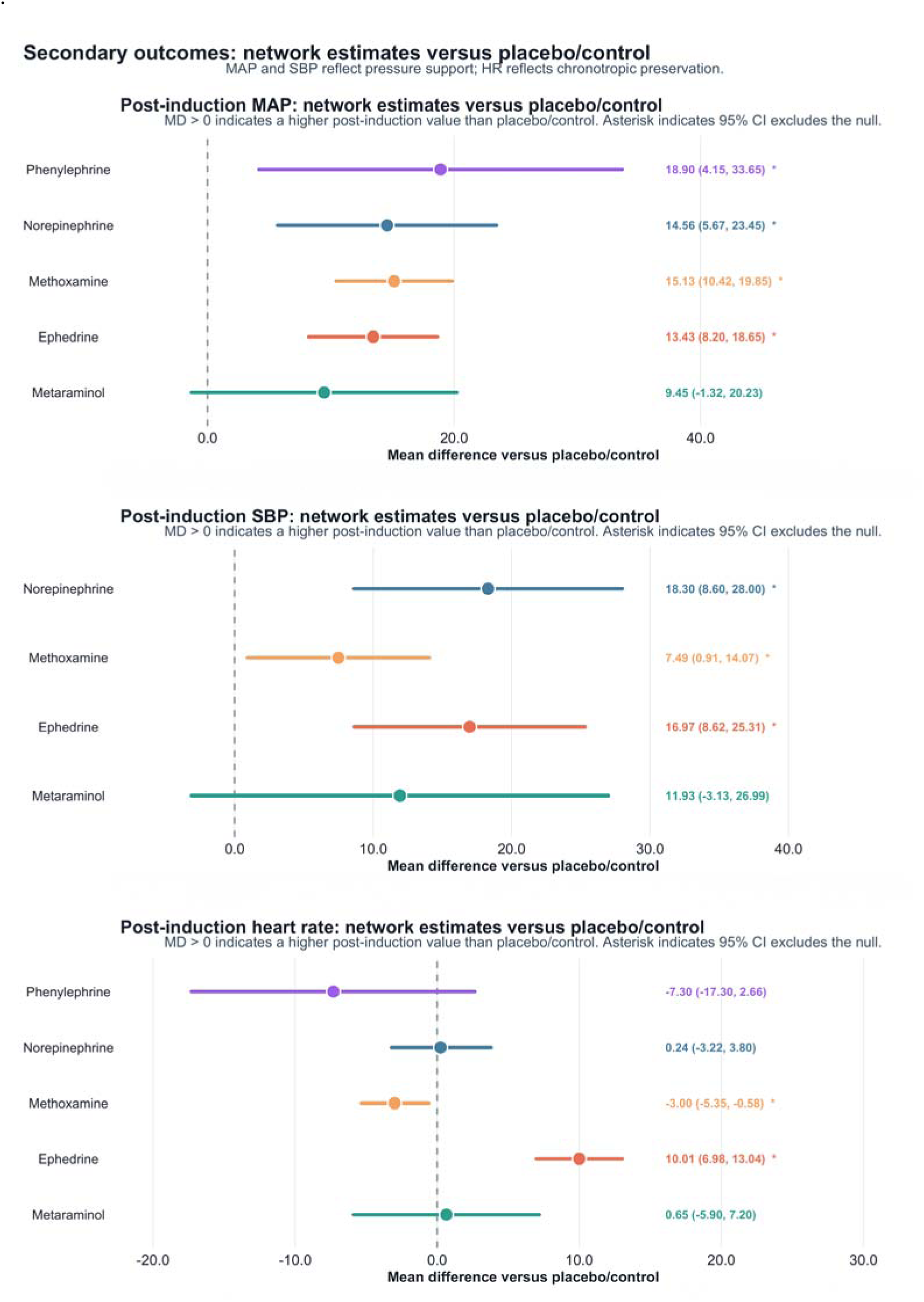
Combined network meta-analysis forest plots for secondary haemodynamic outcomes. Summary mean differences compare each active prophylactic vasopressor with placebo/control for post-induction mean arterial pressure, systolic arterial pressure and heart rate. Points represent network estimates, horizontal lines represent 95% confidence intervals, and right-side labels show mean differences with 95% confidence intervals. Positive mean differences indicate higher post-induction values with the active vasopressor; for MAP and SBP, positive values indicate greater pressure support, whereas for HR, positive values indicate greater heart-rate preservation. CI, confidence interval; HR, heart rate; MAP, mean arterial pressure; MD, mean difference; SBP, systolic arterial pressure.

**Figure 5.**
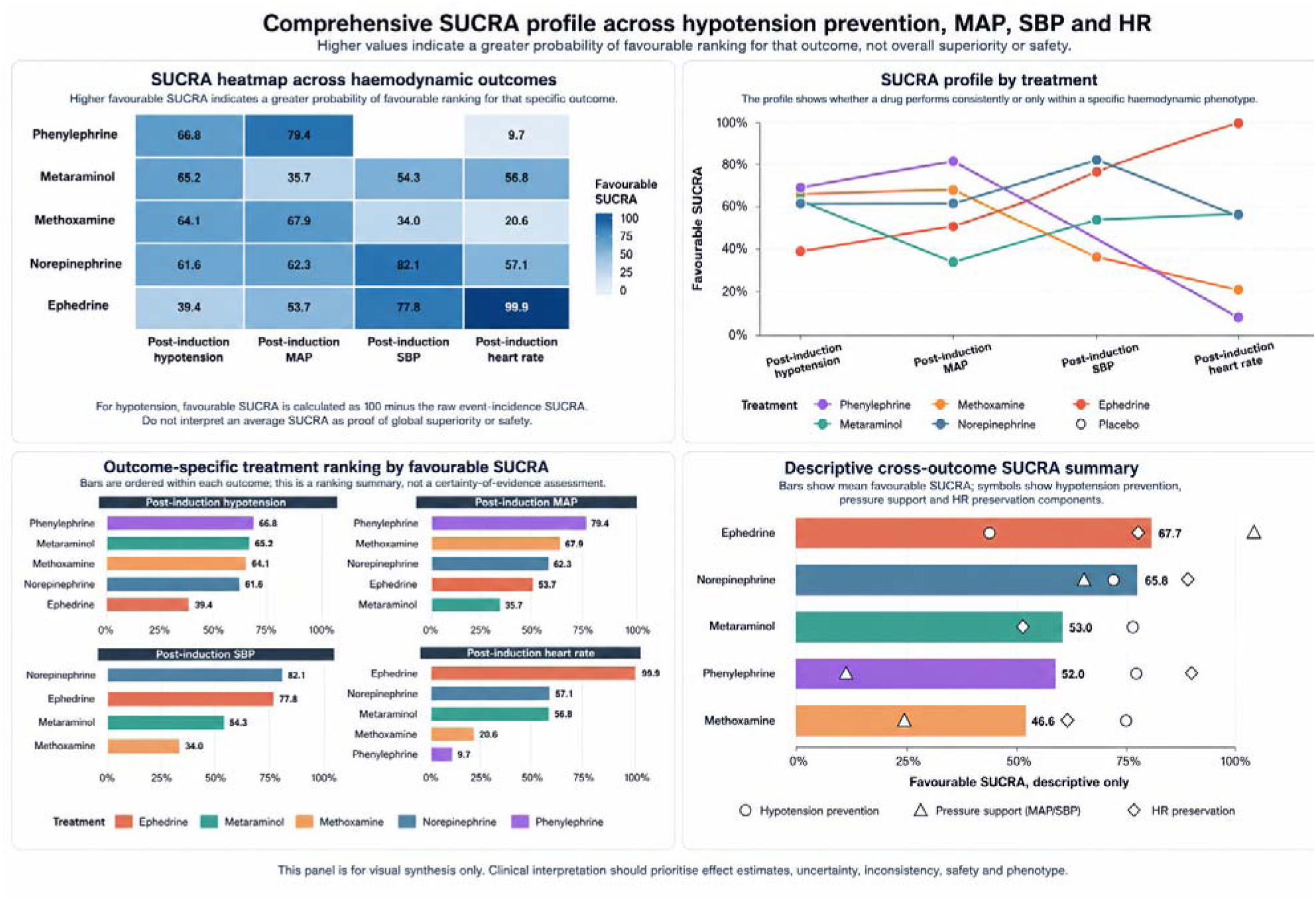
Comprehensive SUCRA profile across hypotension prevention, MAP support, SBP support and HR preservation. (A) Heatmap of favourable SUCRA percentages for each treatment-outcome combination. (B) SUCRA profile by treatment, showing whether a drug performs consistently or mainly within a specific haemodynamic phenotype. (C) Outcome-specific treatment ranking by favourable SUCRA. (D) Descriptive cross-outcome SUCRA summary. Higher values indicate a greater probability of favourable ranking for that outcome, not overall superiority, certainty of evidence or safety. For hypotension, favourable SUCRA was calculated as 100 minus the raw event-incidence SUCRA. MAP, mean arterial pressure; SBP, systolic arterial pressure; HR, heart rate; SUCRA, surface under the cumulative ranking curve.

#### Inconsistency and certainty

Global inconsistency was detected for post-induction SBP (chi-square 13.69; P=0.0002). No statistically significant global inconsistency was detected for incidence of post-induction hypotension (chi-square 1.36; P=0.7158), MAP (chi-square 0.13; P=0.7134) or HR (chi-square 2.05; P=0.1527). Node-splitting did not identify statistically significant local inconsistency in individual comparisons, although indirect estimates were often imprecise because several active nodes were informed by few studies. These inconsistency and imprecision signals strengthen the case for profile-based interpretation rather than ranking-based overstatement.

#### Applicability by induction hypnotic

Induction hypnotic was a prespecified clinical effect modifier because propofol, etomidate and mixed regimens may differ in baseline risk of post-induction hypotension. Propofol-based or propofol-containing induction contributed most evidence across active nodes. Etomidate-predominant studies were fewer and clustered within a limited set of comparisons rather than spanning all active agents. A separate etomidate-only hierarchy was therefore not presented because the available comparisons were more suitable for applicability assessment than for independent treatment ranking. The estimates are most directly applicable to propofol-based or propofol-containing induction. Etomidate-predominant induction represents a different baseline-risk context in which prophylaxis should be individualised.

#### Adverse events and safety reporting

Safety reporting varied across trials and was less complete than reporting of blood-pressure and HR outcomes. Bradycardia was the most consistently extractable haemodynamic adverse event, whereas tachycardia, reactive hypertension and other adverse events were reported inconsistently. The available profile suggested predictable trade-offs: phenylephrine and methoxamine increased arterial pressure but tended to lower HR; ephedrine preserved HR but may be unsuitable when tachycardia or increased myocardial oxygen demand is a concern; and low-dose norepinephrine appeared comparatively HR-neutral in the current network. These short-term haemodynamic outcomes should not be interpreted as evidence that any prophylactic agent reduces myocardial injury, acute kidney injury, stroke, delirium or mortality.

**Table 1.**
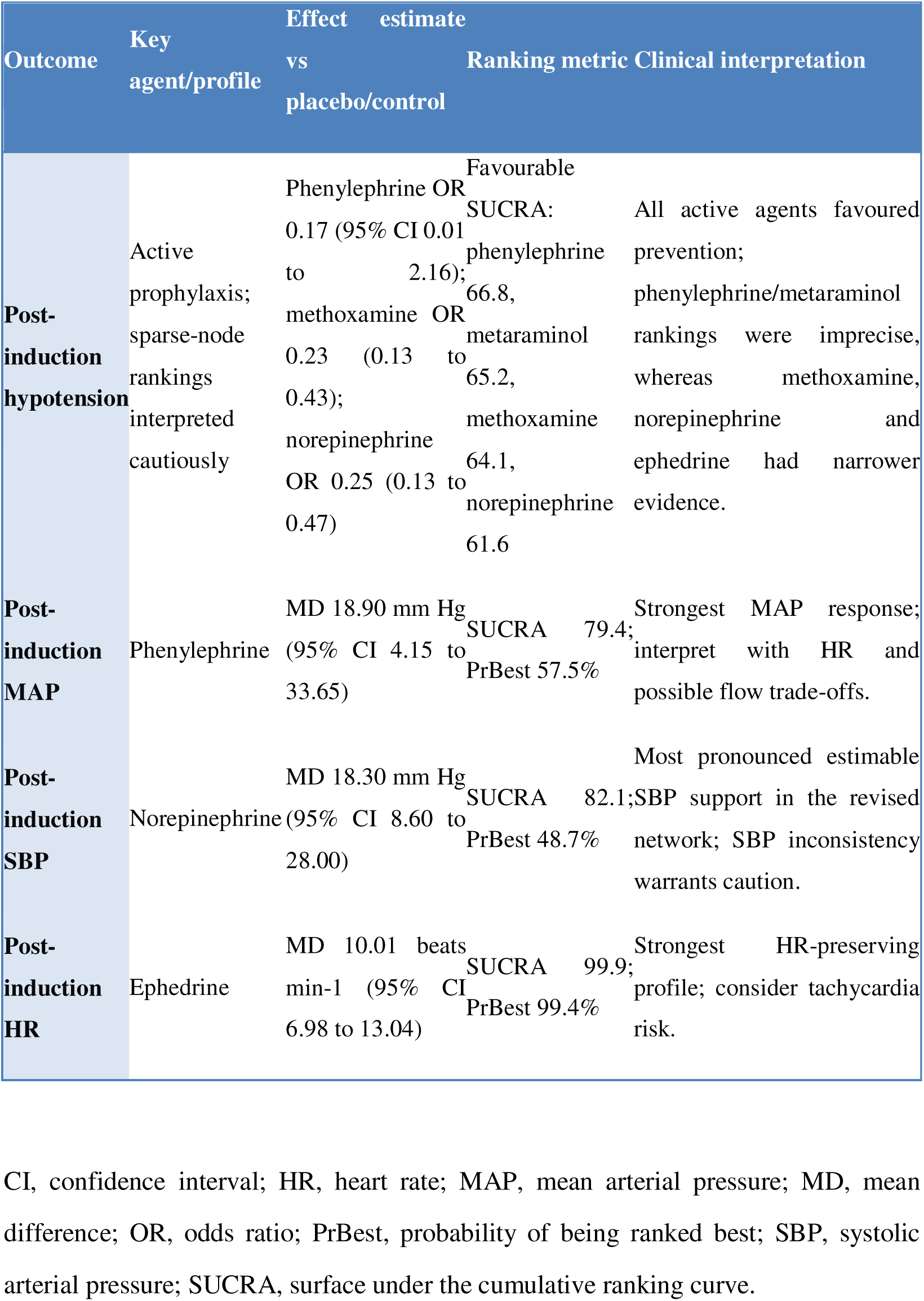
Key network estimates and haemodynamic interpretation.

## Discussion

### Principal findings

This network meta-analysis frames a common induction-period decision: which prophylactic vasopressor, if any, should be used before hypotension develops in older adults undergoing general anaesthesia. Across 31 randomised trials and 2,821 participants in the revised network, active prophylaxis generally favoured lower post-induction hypotension compared with inactive control. Secondary haemodynamic outcomes separated the agents into clinically recognisable profiles. Methoxamine, norepinephrine and ephedrine had the most precise hypotension-prevention estimates, whereas phenylephrine and metaraminol were directionally favourable but imprecise. Norepinephrine combined a clear pressure-support effect with a near-neutral estimated effect on HR, and therefore remained a comparatively balanced preventive option in the current network. Phenylephrine provided the greatest MAP response among estimable nodes, norepinephrine provided the greatest SBP response, and ephedrine preserved HR most clearly. This profile-based interpretation supports practical drug selection while avoiding reliance on treatment rankings alone.

### How the findings address the clinical controversy

The findings clarify rather than remove the clinical controversy. They support prophylaxis most strongly when the anticipated mechanism is loss of vascular tone rather than isolated preload deficit. In a euvolaemic older patient at high risk of propofol-opioid vasodilatation, waiting for rescue treatment may expose the patient to avoidable depth and duration of hypotension. In that setting, a low-dose, titratable vasopressor strategy is mechanistically coherent and consistent with the direction of randomised evidence. Conversely, when hypotension risk is driven mainly by hypovolaemia, bleeding, fasting-related volume deficit or poor preload, fluid optimisation and correction of the underlying cause remain essential. This analysis therefore does not support routine vasopressor use for every older patient. It identifies the clinical conditions under which prophylaxis may function as targeted prevention.

#### Norepinephrine as a balanced preventive strategy

The norepinephrine signal is biologically plausible. Propofol-based induction reduces sympathetic tone, increases venous capacitance and lowers systemic vascular resistance.[6-12] Norepinephrine directly restores vascular tone and has modest beta-1 activity, which may help preserve cardiac output compared with pure alpha-agonism. Recent randomised evidence is consistent with this interpretation. In high-risk patients undergoing major abdominal surgery, early norepinephrine reduced post-induction hypotension compared with repeated ephedrine boluses.[58] In cardiac surgery patients, prophylactic norepinephrine infusion has also been tested to reduce severe hypotension during induction.[59] Norepinephrine should nevertheless be described as a comparatively balanced agent in the current revised network, not as a universal first-line drug or as the single best-ranked agent for every outcome. Its benefit is most plausible when the expected problem is vasodilatation, when frequent blood-pressure measurement or arterial monitoring is available, and when infusion preparation and titration can be performed safely.

#### Alpha-agonists: pressure support with flow-related trade-offs

Phenylephrine ranked highest for MAP support, consistent with alpha-1-predominant pharmacology; however, extractable SBP evidence did not support a connected phenylephrine node in the revised network. A stronger pressor effect should not be equated with a more favourable overall haemodynamic profile. Alpha-mediated vasoconstriction may increase arterial pressure at the cost of reflex bradycardia, increased afterload and reduced stroke volume or cardiac output. Phenylephrine may therefore be appropriate for isolated vasodilatation with preserved cardiac-output reserve, but less attractive in frail older patients with low preload, bradycardia, diastolic dysfunction or ventricular impairment. Methoxamine reduced post-induction hypotension and increased MAP, although its longer duration and alpha-predominant profile may limit titratability. Its evidence base was largely derived from small, incompletely reported Chinese-language trials. Metaraminol showed favourable but imprecise estimates, supporting its use as a context-dependent alpha-agonist option rather than an established preferred strategy.

#### Ephedrine: chronotropic profile

Ephedrine reduced post-induction hypotension and showed the clearest HR-preserving profile. It is therefore most relevant when chronotropic support is desirable, particularly in patients with bradycardia or low baseline HR. Its mixed direct and indirect sympathomimetic action explains its long-standing use during propofol-based induction in older adults. The same chronotropic and catecholamine-releasing properties may be undesirable in patients vulnerable to tachyarrhythmia, coronary ischaemia, uncontrolled hypertension or myocardial oxygen supply-demand imbalance. The revised network therefore strengthens the focus on agents with clearer prophylactic profiles and more clinically interpretable titration during induction.

### Induction hypnotic as an effect modifier

Induction hypnotic is a plausible effect modifier because propofol and etomidate create different baseline risks of hypotension. The benefit of prophylactic vasopressors is therefore likely to depend partly on the hypnotic regimen used. Most evidence in this network came from propofol-based or propofol-containing induction, making the findings most directly applicable to this higher-risk context. Etomidate-predominant studies were fewer and unevenly distributed across treatment nodes, so they inform applicability rather than a separate drug hierarchy. Future trials should stratify randomisation by hypnotic regimen and test whether vasopressor effects differ across induction strategies.

### Strengths and limitations

The key strength of this review is its clinically precise scope. It focused on prophylactic administration during induction in older adults, a decision window that is predictable, short and directly modifiable, and separated this question from rescue treatment of established intraoperative hypotension. The inclusion of English- and Chinese-language trials expanded the evidence base for agents that are widely used in practice but under-represented in English-only reviews. By comparing multiple active drugs in a connected network and by analysing hypotension, MAP, SBP and HR together, the study moves beyond a single blood-pressure ranking and provides a practical haemodynamic profile for each drug. Several limitations should also be considered. Definitions of post-induction hypotension, vasopressor dosing, route of administration and anaesthetic regimens varied across studies. Some active nodes, particularly phenylephrine and metaraminol, were informed by fewer trials than norepinephrine, ephedrine and methoxamine. Safety reporting was inconsistent, and most trials focused on immediate haemodynamic outcomes rather than myocardial injury, acute kidney injury or mortality. The detected global inconsistency for SBP further supports cautious phenotype-based interpretation rather than ranking-based overstatement.

### Future directions

Future trials should move from placebo-controlled efficacy testing towards active-comparator, phenotype-guided strategies. Priority designs include comparisons of low-dose norepinephrine infusion with phenylephrine- or ephedrine-based protocols during propofol-based induction, with stratification by frailty, chronic hypertension, baseline HR, cardiac function and hypnotic regimen. Trials should capture immediate haemodynamic stability and clinically relevant outcomes such as myocardial injury, acute kidney injury, delirium, stroke and mortality. A shared core outcome set for induction hypotension would improve comparability and strengthen future evidence synthesis.

### Conclusions

This systematic review and network meta-analysis supports prophylactic vasopressor selection as a practical strategy for reducing post-induction hypotension in older adults undergoing general anaesthesia. In the revised network, the findings continue to favour a phenotype-guided approach rather than a single universal first-line agent. In the current network, methoxamine, norepinephrine and ephedrine had the most precise hypotension-prevention estimates, while phenylephrine and metaraminol were directionally favourable but imprecise. Norepinephrine showed a comparatively balanced profile, combining pressure support with heart-rate neutrality, particularly in propofol-based induction where vasodilatation is expected. Phenylephrine and related alpha-agonists provided strong pressure support and may be most appropriate when vascular tone is the dominant target and HR and cardiac-output reserve are preserved. Ephedrine best matched scenarios in which chronotropic support is also desired. Methoxamine and metaraminol remain context-dependent alpha-agonist options. Overall, these results support a shift from reactive rescue of established hypotension towards phenotype-guided prevention, while highlighting the need for active-comparator trials powered for clinically important outcomes.

**Table 2.**
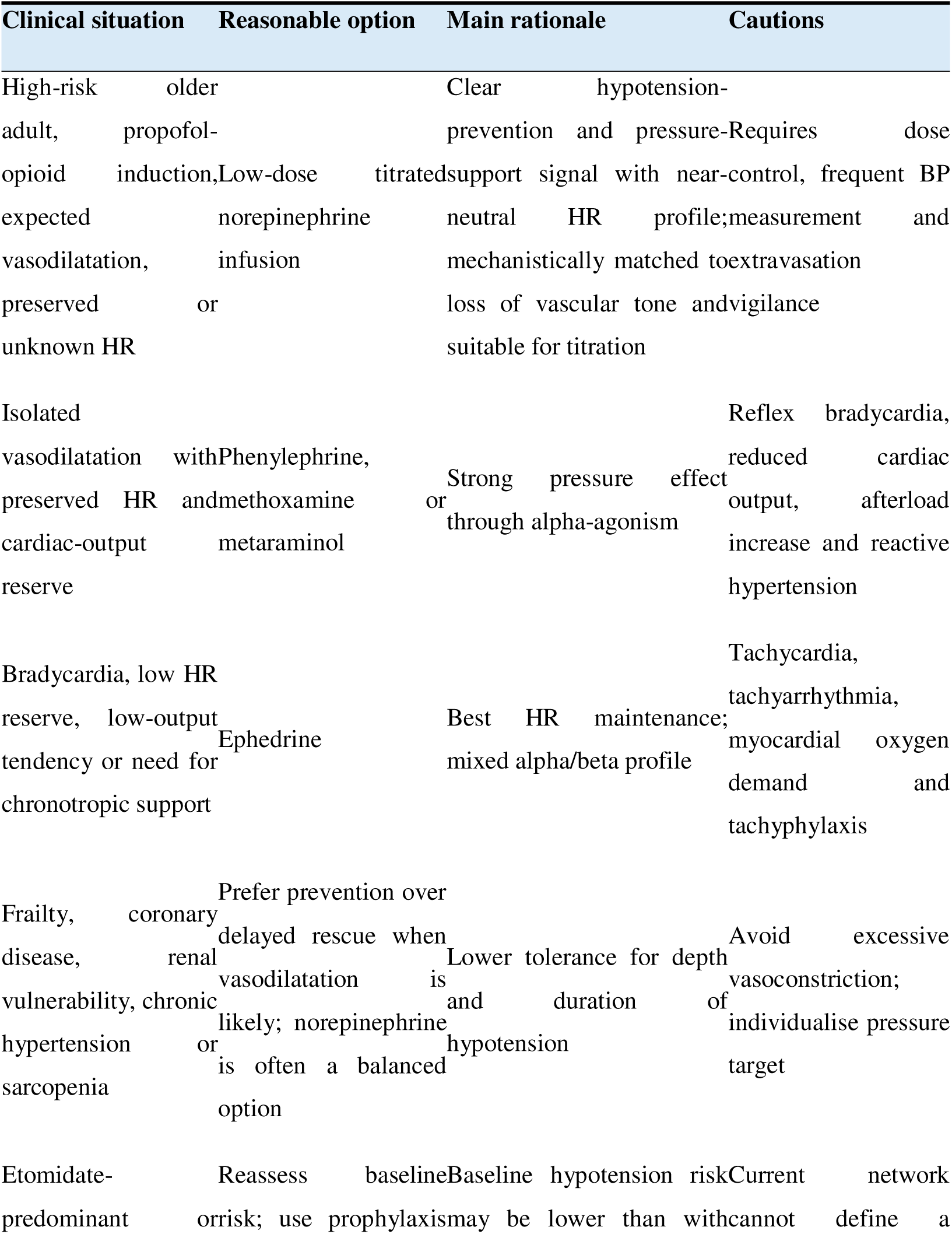
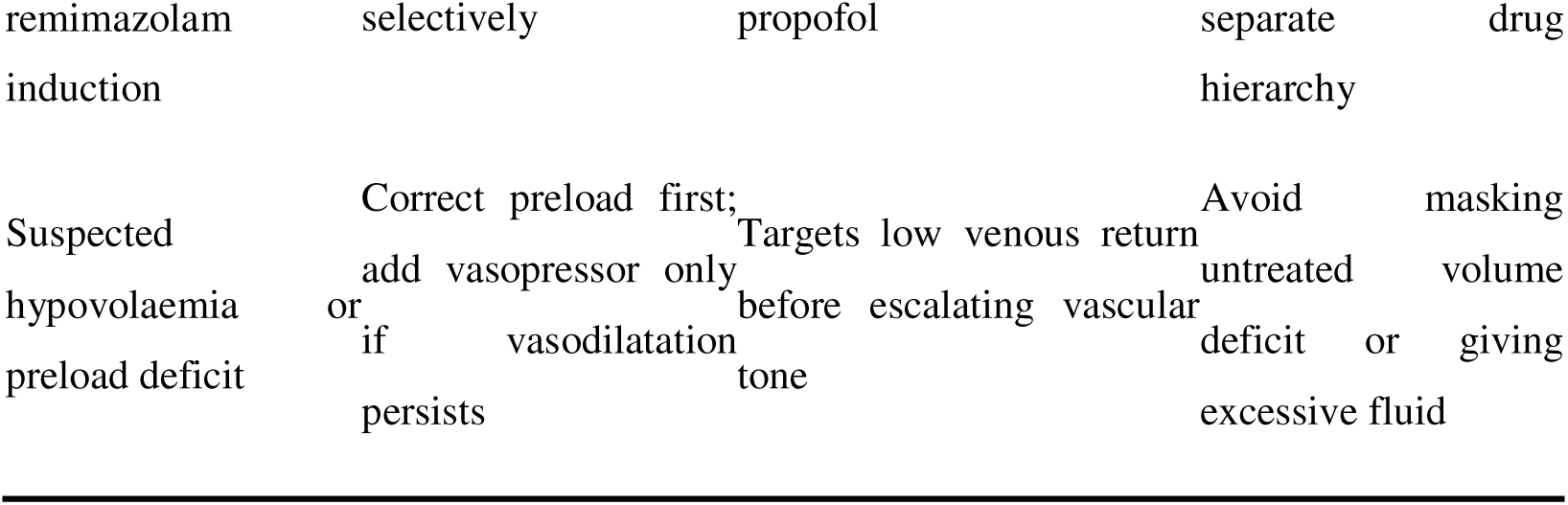
Phenotype-guided framework for prophylactic vasopressor selection.

## Declarations

### Authors’ contributions

The review was designed and conceptualized by H. H.Z. and Z.Y. Z. and Z.Y. Z extracted aggregate data and analysis, Z.Y. Z, D.X.W, C.L.D.,X.Y.D., R.R W analysized data, wrote the draft of the manuscript and drafted the fgure. Z.Y. Z, D.X.W, C.L.D.,X.Y.D., R.R W and H. H.Z reviewed the manuscript and participated in the revisions.

### Data availability

Extracted aggregate data and analysis code will be made available on reasonable request or in an online repository.

## Supplementary Appendix

## Appendix 1: Search strategy

**Table S1.** Search strategy of PubMed.

| Set | Search Query |
| --- | --- |
| #1 | (Aged[MeSH Terms] OR Elderly[Text Word] OR "Older adults"[Text Word] OR Geriatric[Text Word] OR Aging[Text Word]) |
| #2 | ("Anesthesia, General"[MeSH Terms] OR "Anesthesia Induction"[MeSH Terms] OR "Induction of anesthesia"[Text Word] OR "General anesthesia induction"[Text Word]) |
| #3 | ("Vasoconstrictor Agents"[MeSH Terms] OR "Vasoactive agents"[Text Word] OR Norepinephrine[Text Word] OR Phenylephrine[Text Word] OR Ephedrine[Text Word] OR Metaraminol[Text Word] OR Methoxamine[Text Word]) |
| #4 | (Hypotension[MeSH Terms] OR Hypotension[Text Word] OR "Blood Pressure, Low"[Text Word] OR Prevent*[Text Word] OR Prophylactic[Text Word]) |
| #5 | #1 AND #2 AND #3 AND #4 |
| #6 | #5 AND (randomized controlled trial[Filter]) |

**Table S2.**
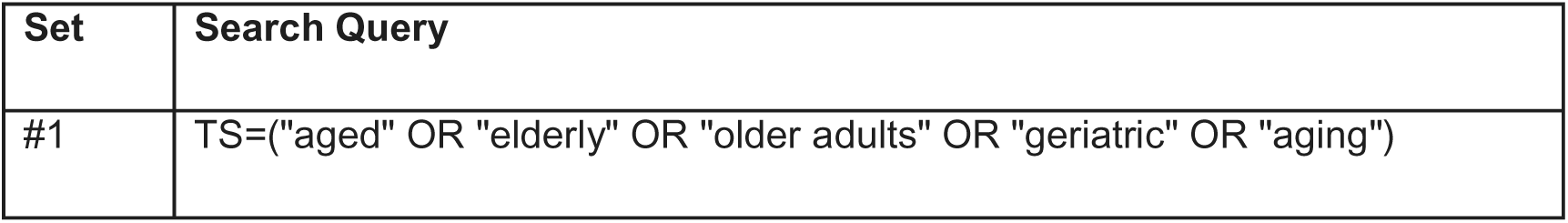
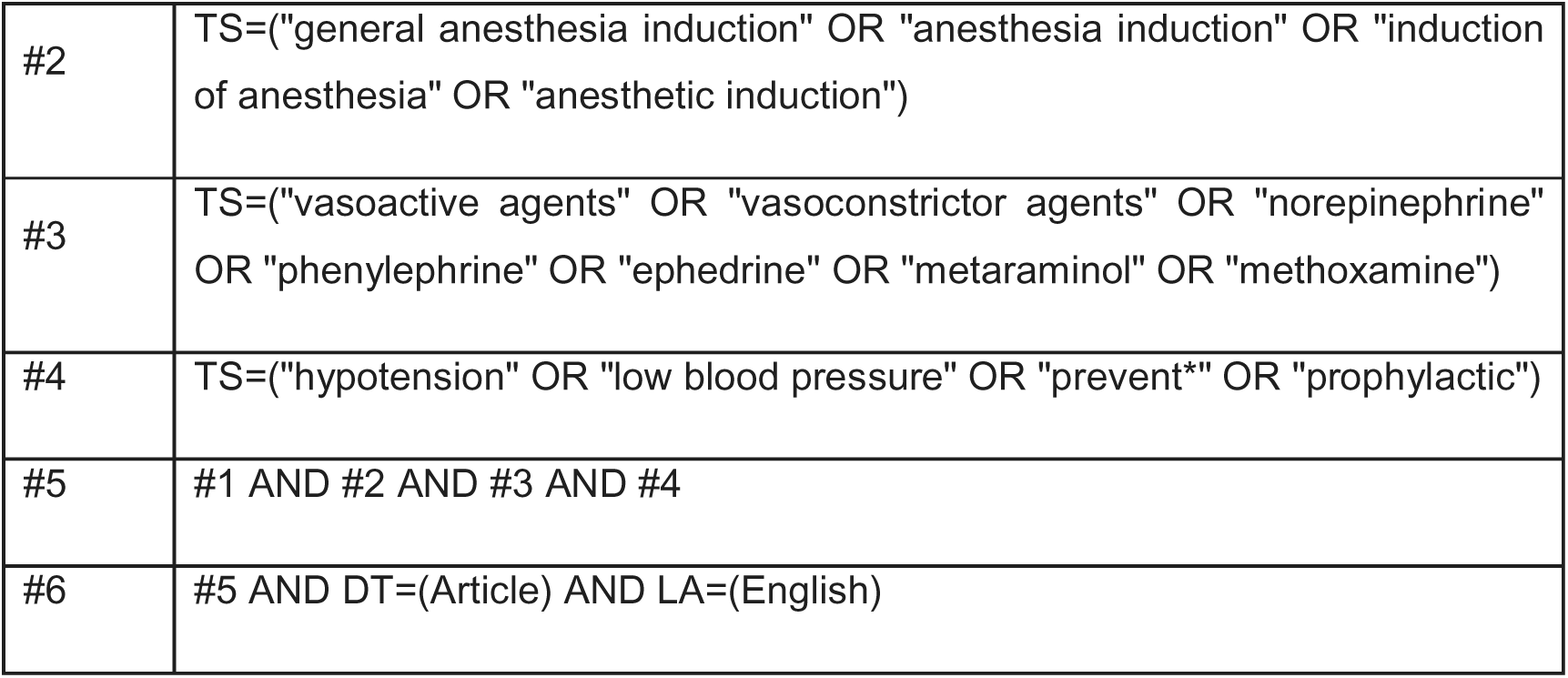
Search strategy of Web of Science: Science Citation Index Expanded.

**Table S3.** Search strategy of Embase.

| Set | Search Query |
| --- | --- |
| #1 | 'aged'/exp OR 'elderly':ti,ab,kw OR aged:ti,ab,kw OR 'older adult*':ti,ab,kw OR geriatric*:ti,ab,kw OR aging:ti,ab,kw OR ageing:ti,ab,kw |
| #2 | 'anesthesia induction'/exp OR 'general anesthesia induction':ti,ab,kw OR 'anaesthesia induction':ti,ab,kw OR 'anesthesia induction':ti,ab,kw OR 'induction of anesthesia':ti,ab,kw OR 'induction of anaesthesia':ti,ab,kw OR 'anesthetic induction':ti,ab,kw OR 'anaesthetic induction':ti,ab,kw |
| #3 | 'vasoactive agent'/exp OR 'vasoconstrictor agent'/exp OR 'vasopressor agent'/exp OR 'norepinephrine'/exp OR 'phenylephrine'/exp OR 'ephedrine'/exp OR 'metaraminol'/exp OR 'methoxamine'/exp OR 'vasoactive agent*':ti,ab,kw OR 'vasoconstrictor agent*':ti,ab,kw OR vasopressor*:ti,ab,kw OR norepinephrine:ti,ab,kw OR noradrenaline:ti,ab,kw OR phenylephrine:ti,ab,kw OR ephedrine:ti,ab,kw OR metaraminol:ti,ab,kw OR methoxamine:ti,ab,kw |
| #4 | 'hypotension'/exp OR hypotension:ti,ab,kw OR 'low blood pressure':ti,ab,kw OR prevent*:ti,ab,kw OR prophylactic*:ti,ab,kw OR prophylaxis:ti,ab,kw |
| #5 | #1 AND #2 AND #3 AND #4 |
| #6 | #5 AND [article]/lim AND [english]/lim |

**Table S4.** Search strategy of Cochrane Central Register of Controlled Trials (CENTRAL)

| Set | Search Query |
| --- | --- |
| #1 | [mh "Aged"] OR (aged OR elderly OR "older adults" OR geriatric OR aging):ti,ab,kw |
| #2 | [mh "Anesthesia, General"] OR [mh "Anesthesia Induction"] OR ("general anesthesia induction" OR "anesthesia induction" OR "induction of anesthesia"):ti,ab,kw |
| #3 | [mh "Vasoconstrictor Agents"] OR (vasoactive OR "vasoconstrictor agents" OR norepinephrine OR phenylephrine OR ephedrine OR metaraminol OR methoxamine):ti,ab,kw |
| #4 | [mh "Hypotension"] OR (hypotension OR "low blood pressure" OR prevent* OR prophylactic):ti,ab,kw |
| #5 | #1 AND #2 AND #3 AND #4 |
| #6 | #5 in Trials |

**Table S5.** Search strategy of CNKI and Wanfang.

| Set | Search Query (Translated for reference) |
| --- | --- |
| #1 | "Elderly" OR "Aged" |
| #2 | "General anesthesia induction" |
| #3 | "Vasoactive agents" OR "Vasoactive drugs" |
| #4 | "Norepinephrine" OR "Phenylephrine" OR "Ephedrine" OR "Metaraminol"<br>OR "Methoxamine" |
| #5 | "Hypotension" OR "Prevention" |
| #6 | #1 AND #2 AND (#3 OR #4) AND #5 |

**Table S6.** Search strategy of VIP.

| Set | Search Query (Translated for reference) |
| --- | --- |
| #1 | "Elderly" OR "Aged" OR "Older adults" OR "Older patients" OR "Ageing" |
| #2 | "General anesthesia" OR "General anaesthesia" OR "General anesthesia induction" OR "Induction of anesthesia" OR "Induction period" |
| #3 | "Vasoactive agents" OR "Vasoactive drugs" OR "Vasopressors" OR "Pressor agents" OR "Vasoconstrictors" |
| #4 | "Norepinephrine" OR "Phenylephrine" OR "Ephedrine" OR "Metaraminol"<br>OR "Methoxamine" |
| #5 | "Hypotension" OR "Blood pressure decrease" OR "Prevention" OR "Prophylaxis" OR "Pretreatment" OR "Pre-injection" |
| #6 | #1 AND #2 AND (#3 OR #4) AND #5 |

## Appendix 2: Characteristics of included studies

**Table S2.1:**
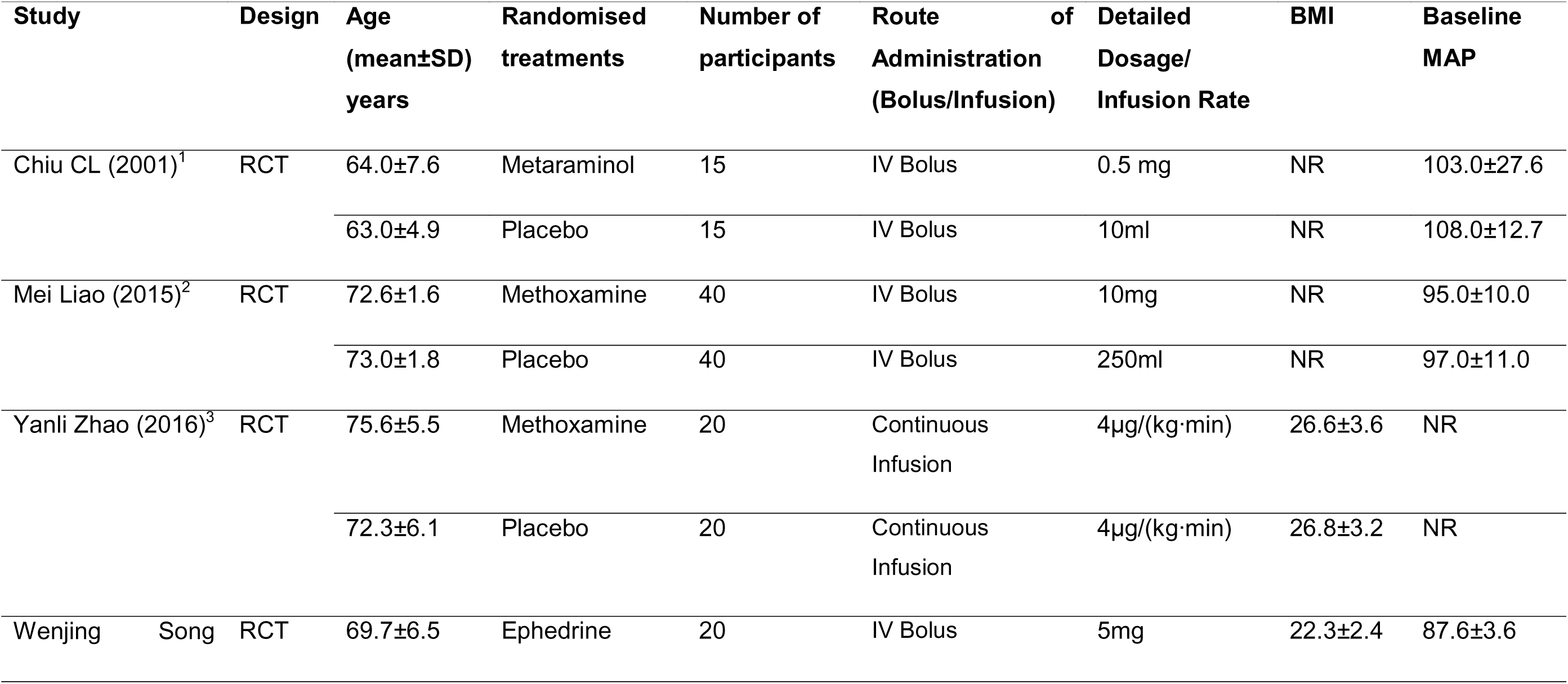
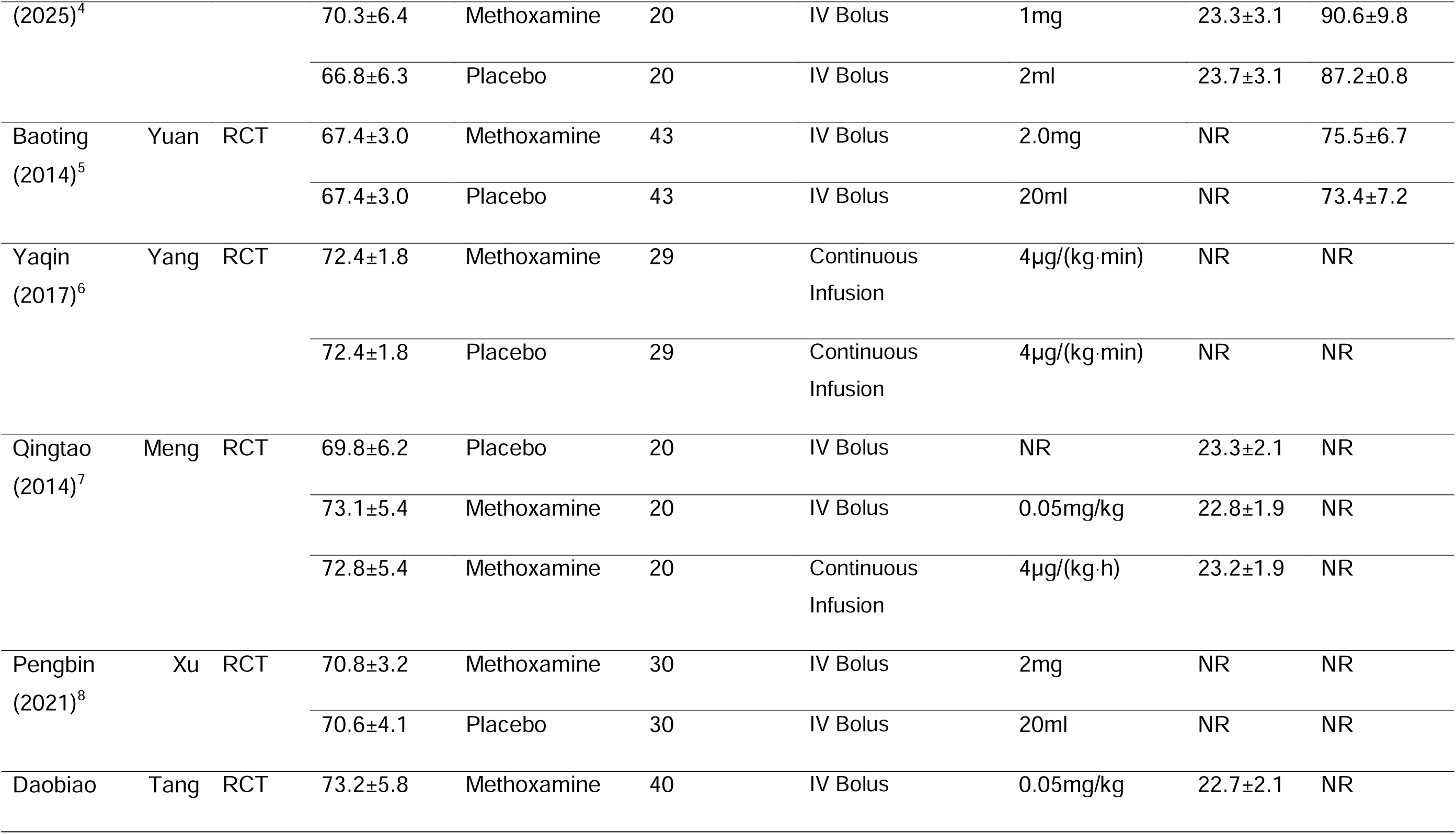
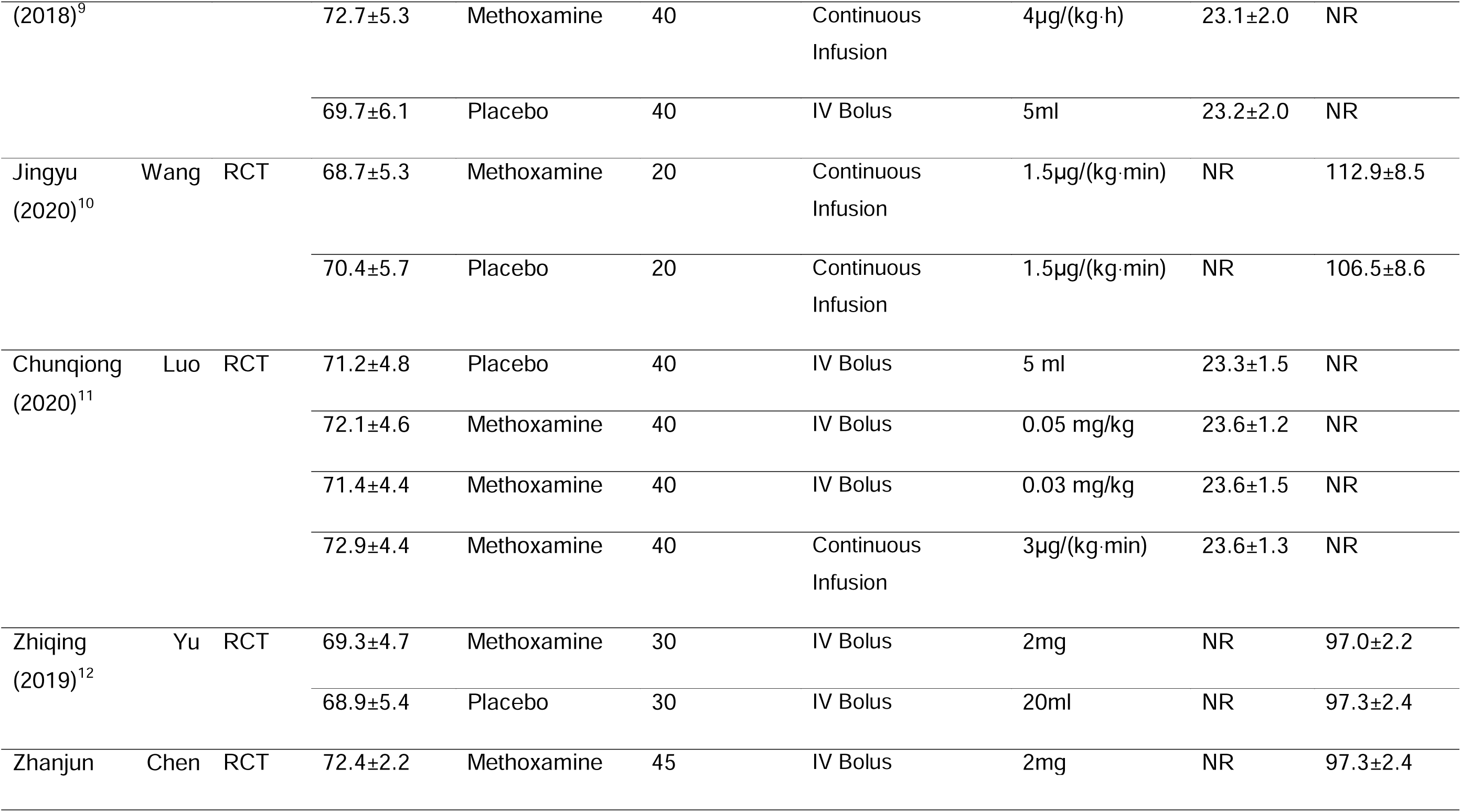
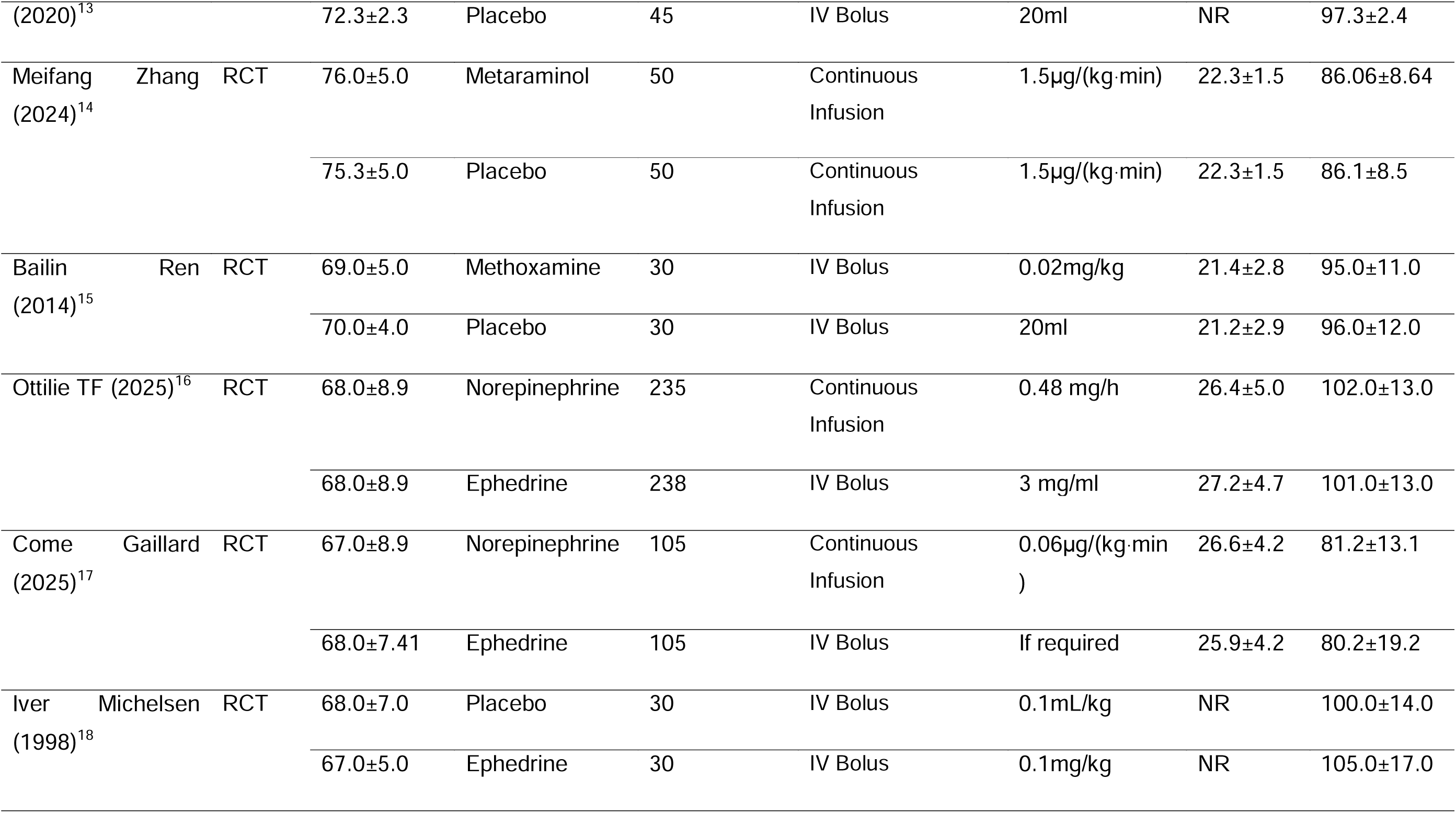
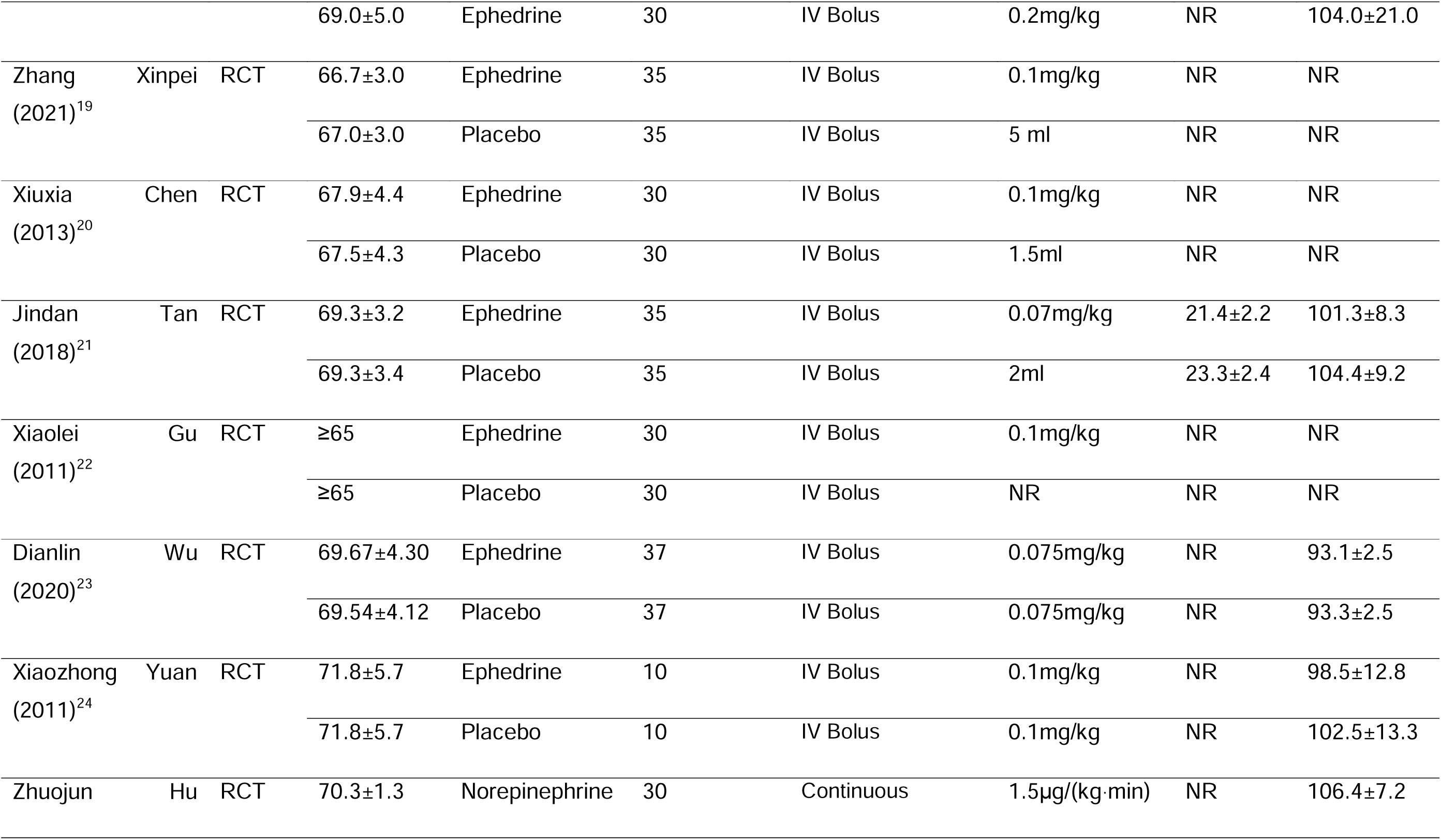
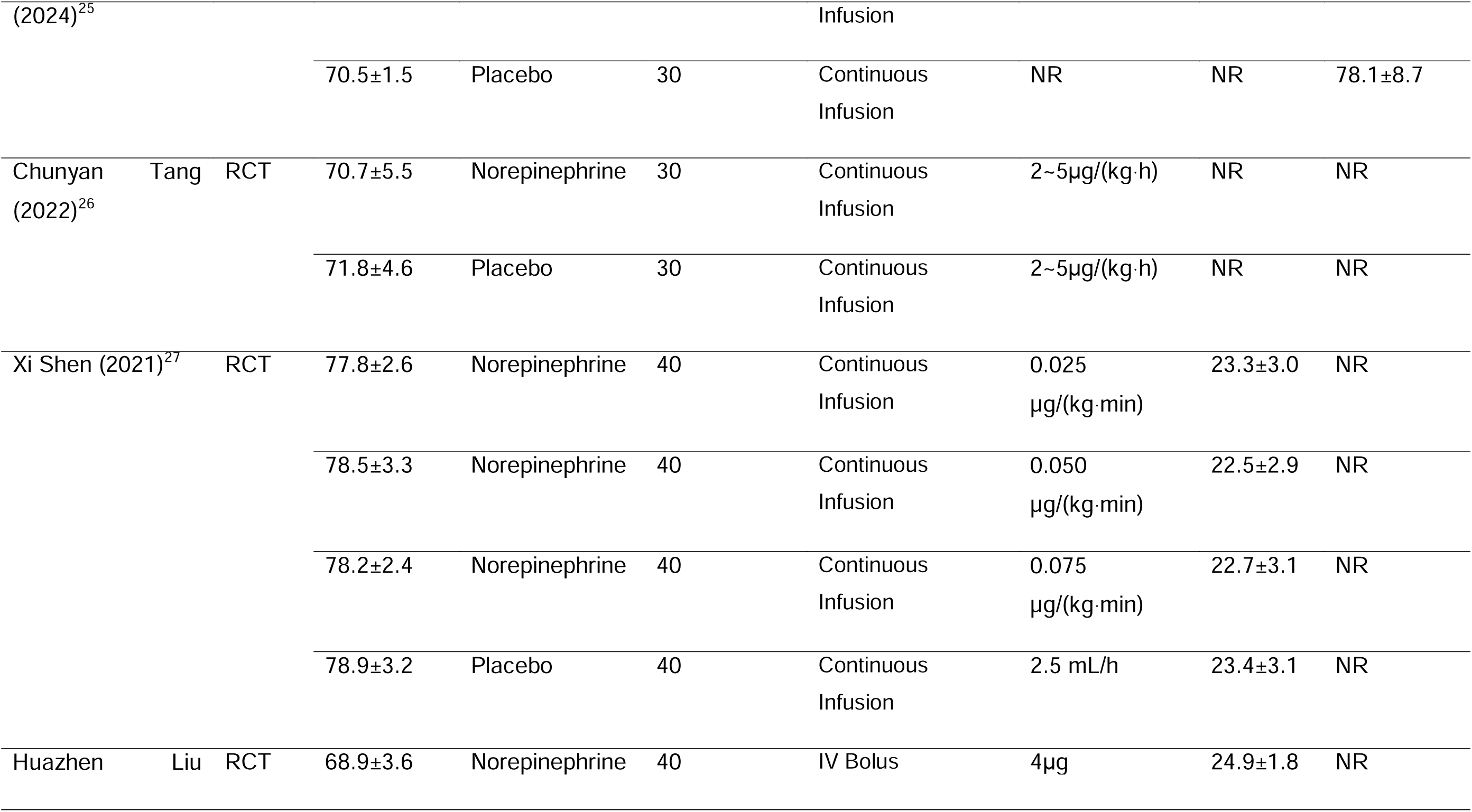
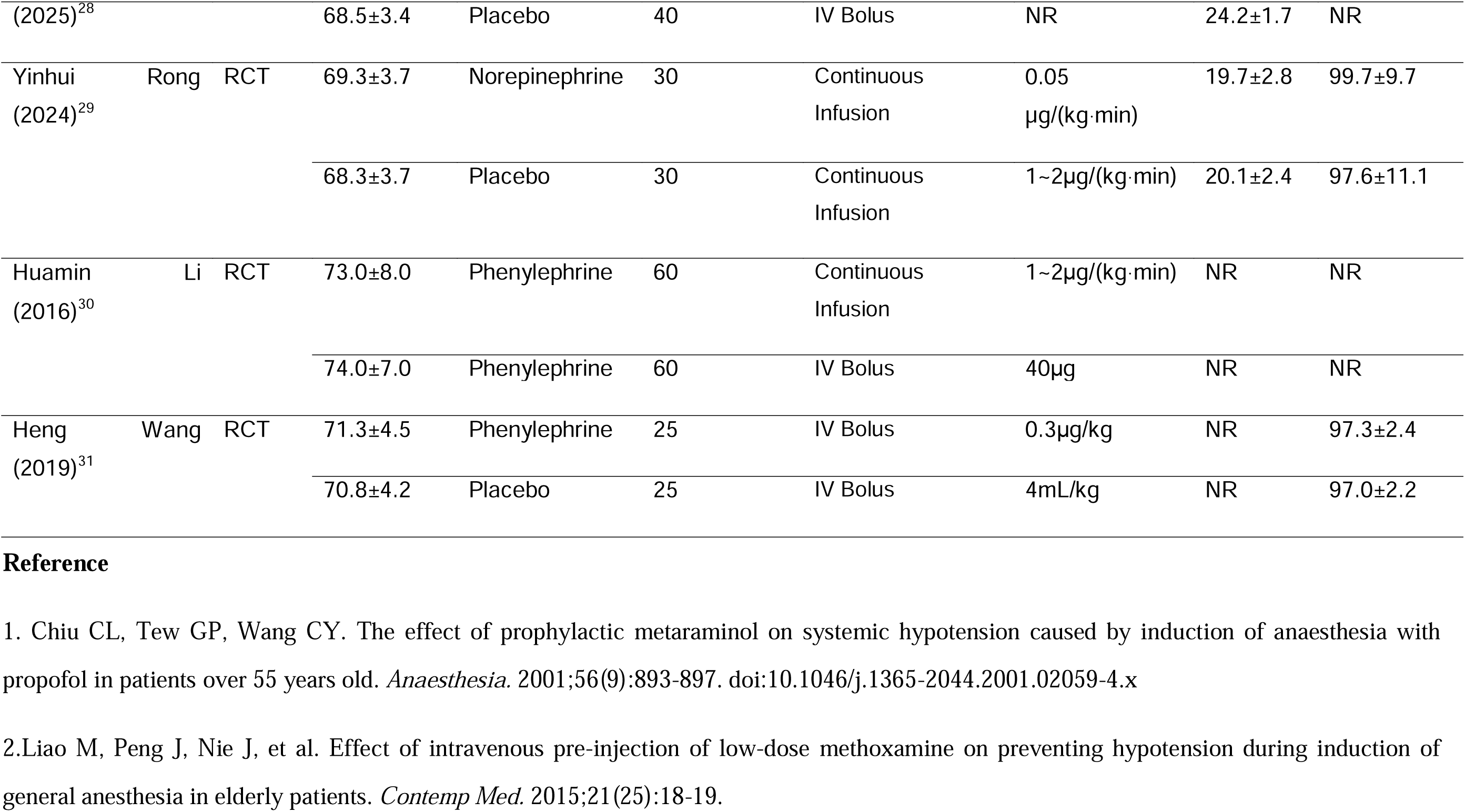
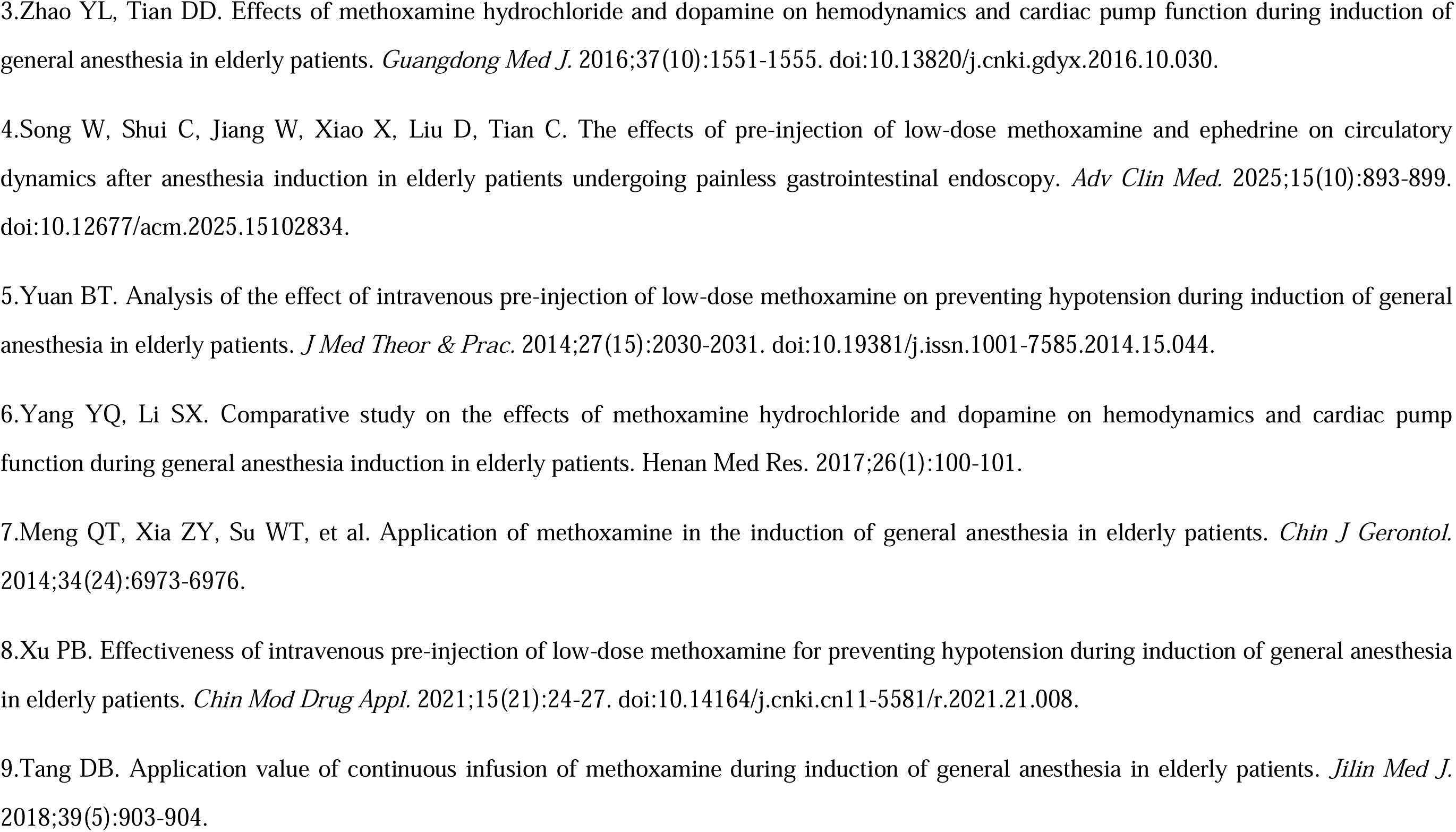
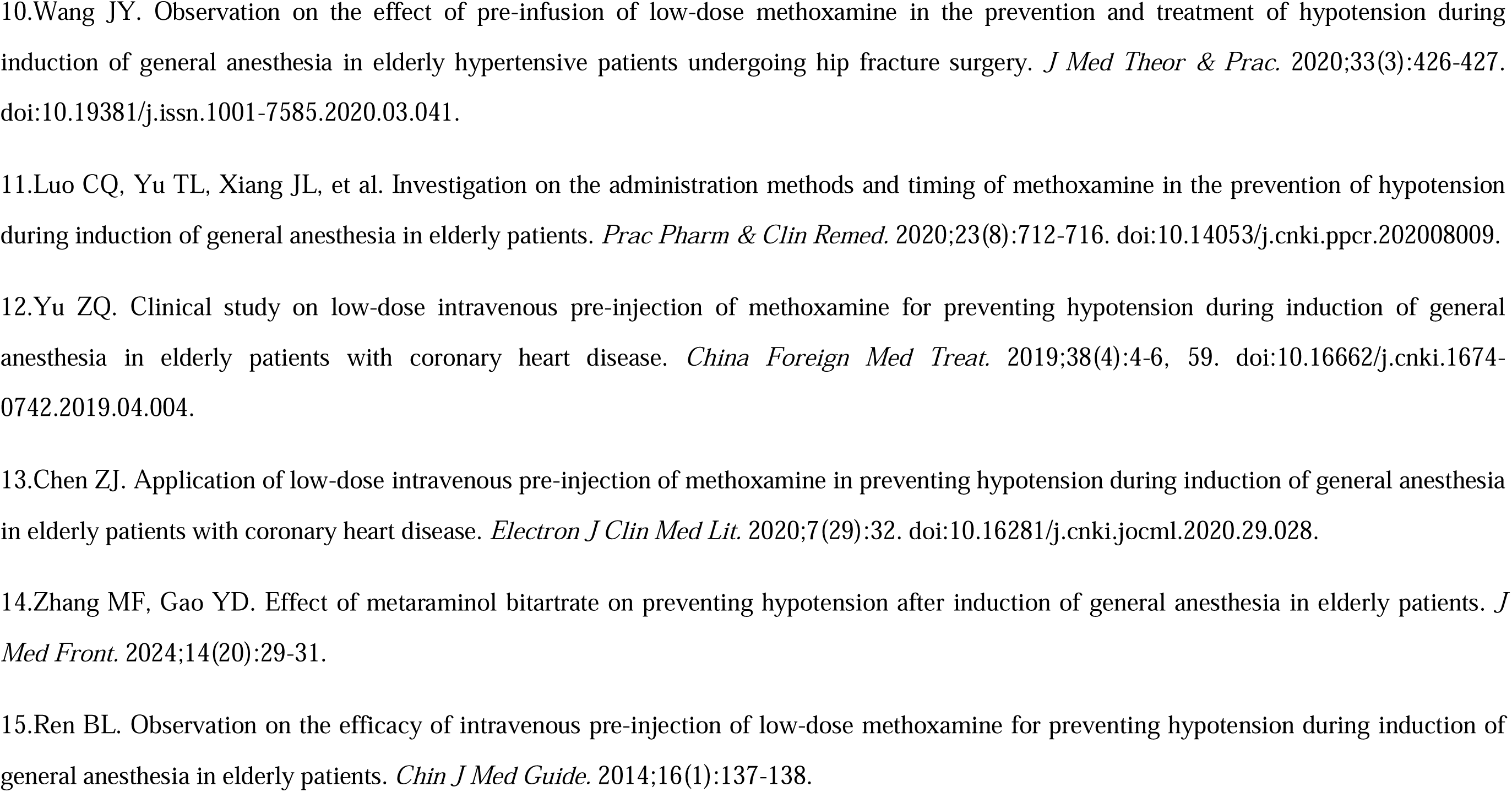
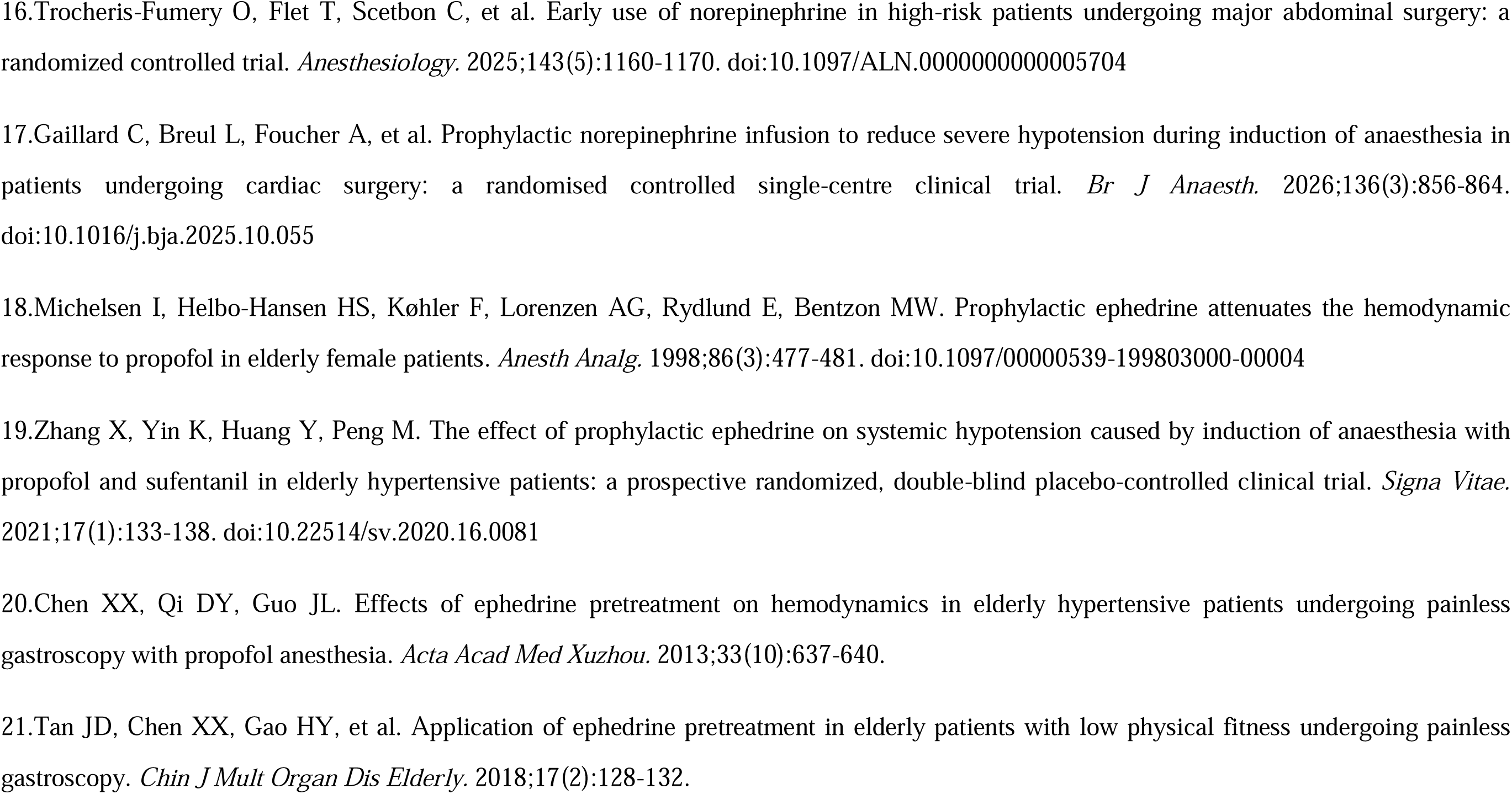
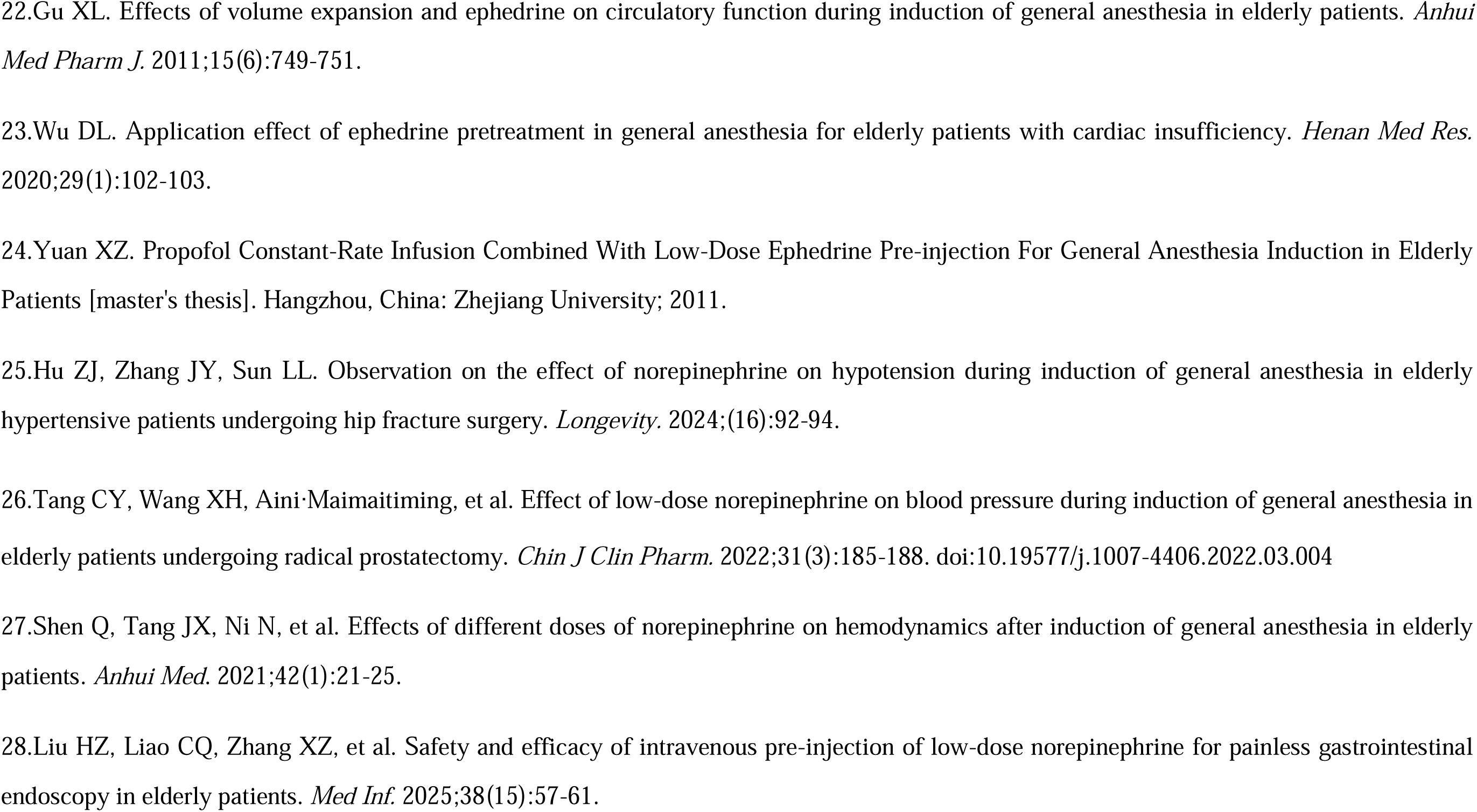
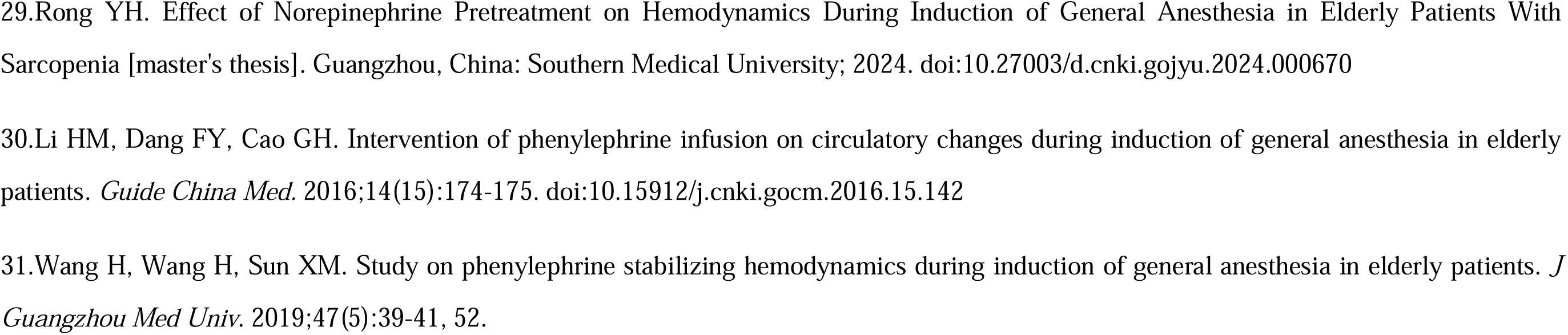
Baseline of characteristics of included studies.

**Table S2.2:**
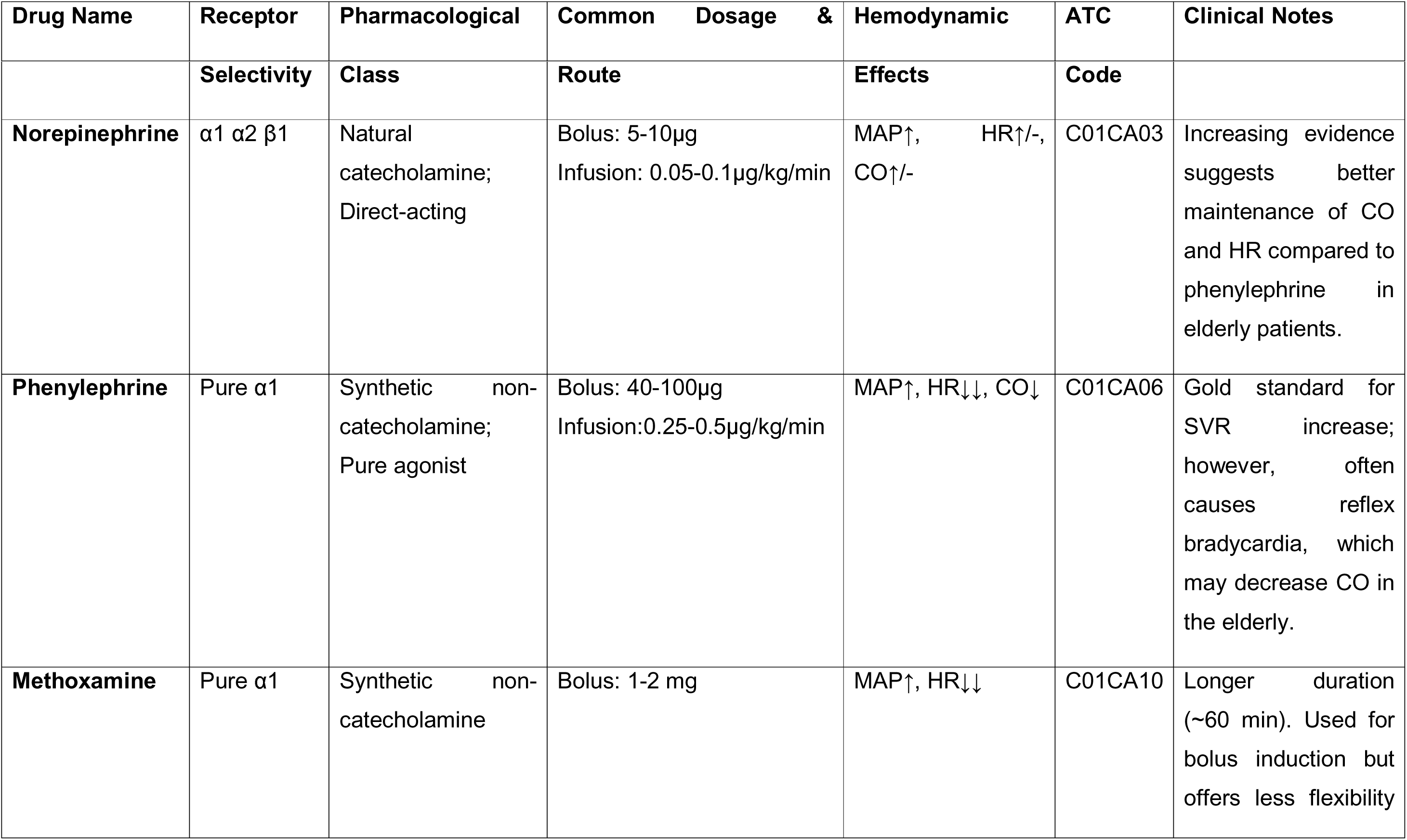
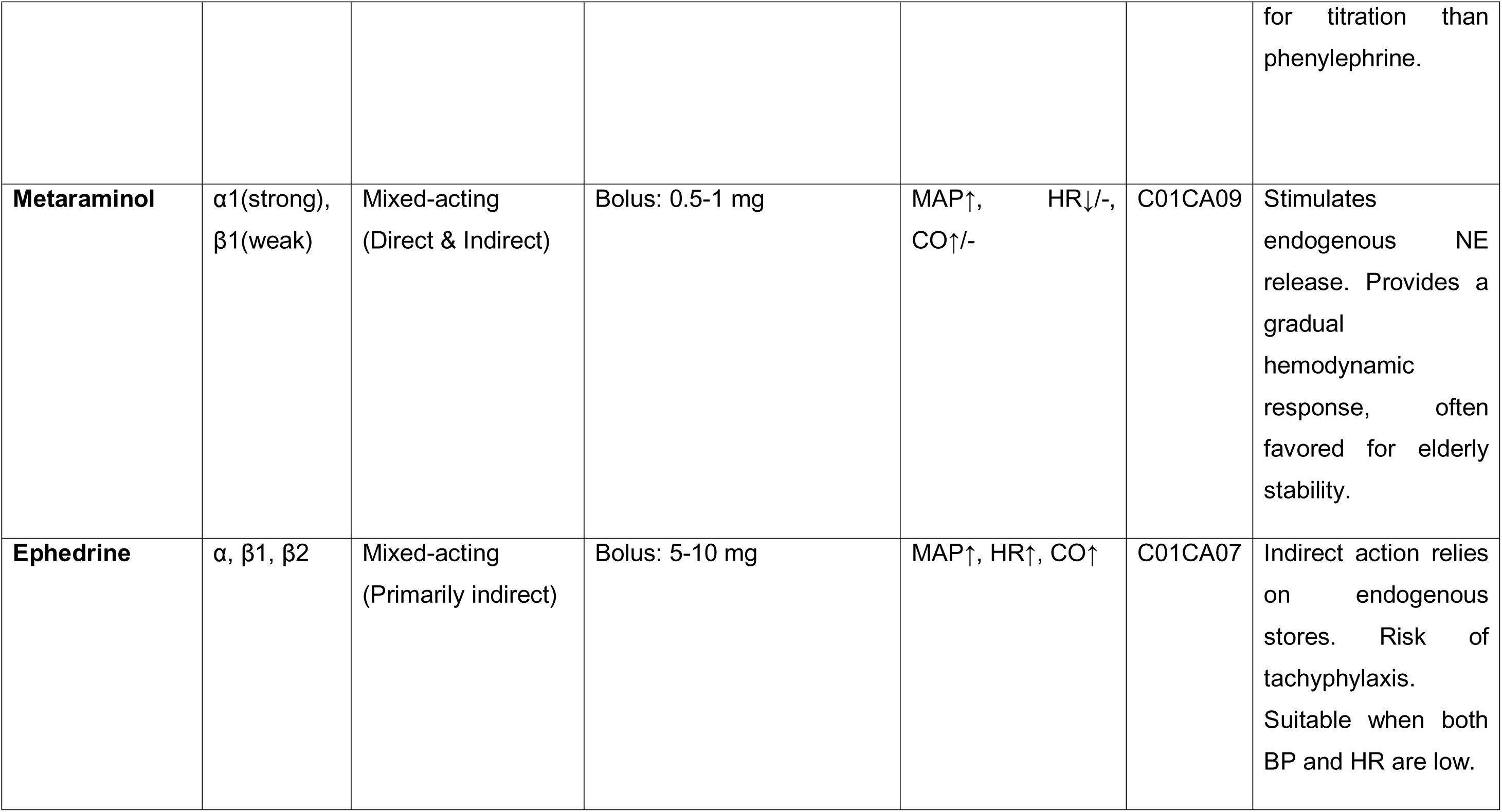
Characteristics of Vasoactive Drugs for Preventing Hypotension During Anesthesia Induction in the Elderly.

## Appendix 3: Data extraction items

| Data Category | List of Variables |
| --- | --- |
| Study Information | Study ID (Author, Year),<br>Number of participants (n/N per group) |
| Patients' Baseline | Age (mean±SD, years), BMI (kg/m<sup>2</sup>) |
| Interventions | Randomized treatments,<br>Route of Administration (Bolus/Infusion), Detailed Dosage,<br>Timing of Administration |
| Efficacy Outcomes | Definition of hypotension,<br>Incidence of hypotension (n/N), Baseline MAP (T0),<br>Endpoint MAP (T1),<br>MAP Endpoint timing |
| Adverse Events | Incidence of Bradycardia (n/N) |

## Appendix 4: Risk of bias of randomized clinical trials

**Figure S4.**
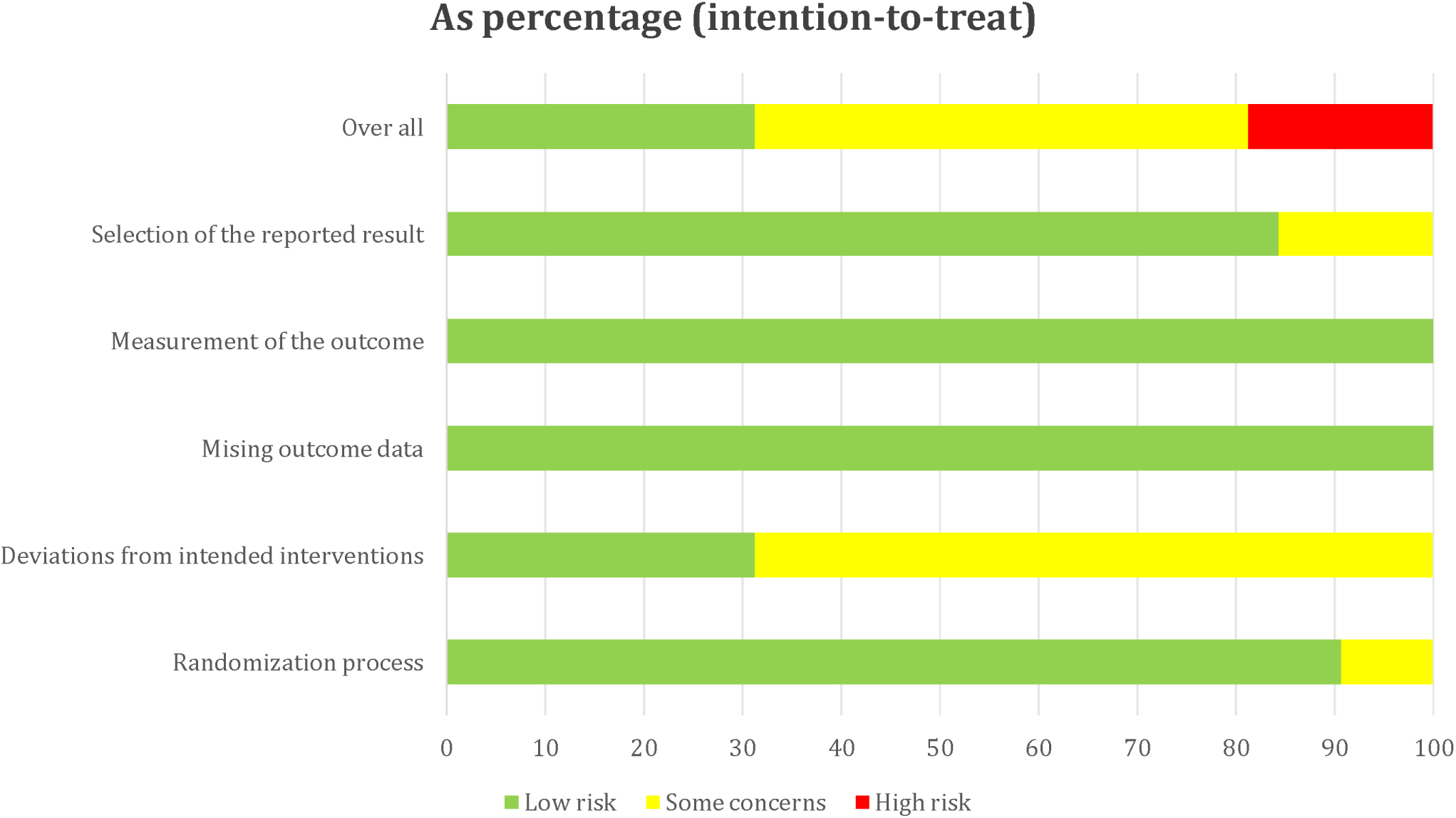
Overall risk of bias presented as percentage of each risk of bias item across all included studies. Green□=□Low risk, Red□=□High risk, Yellow□=□Some concerns.

**Table S4:**
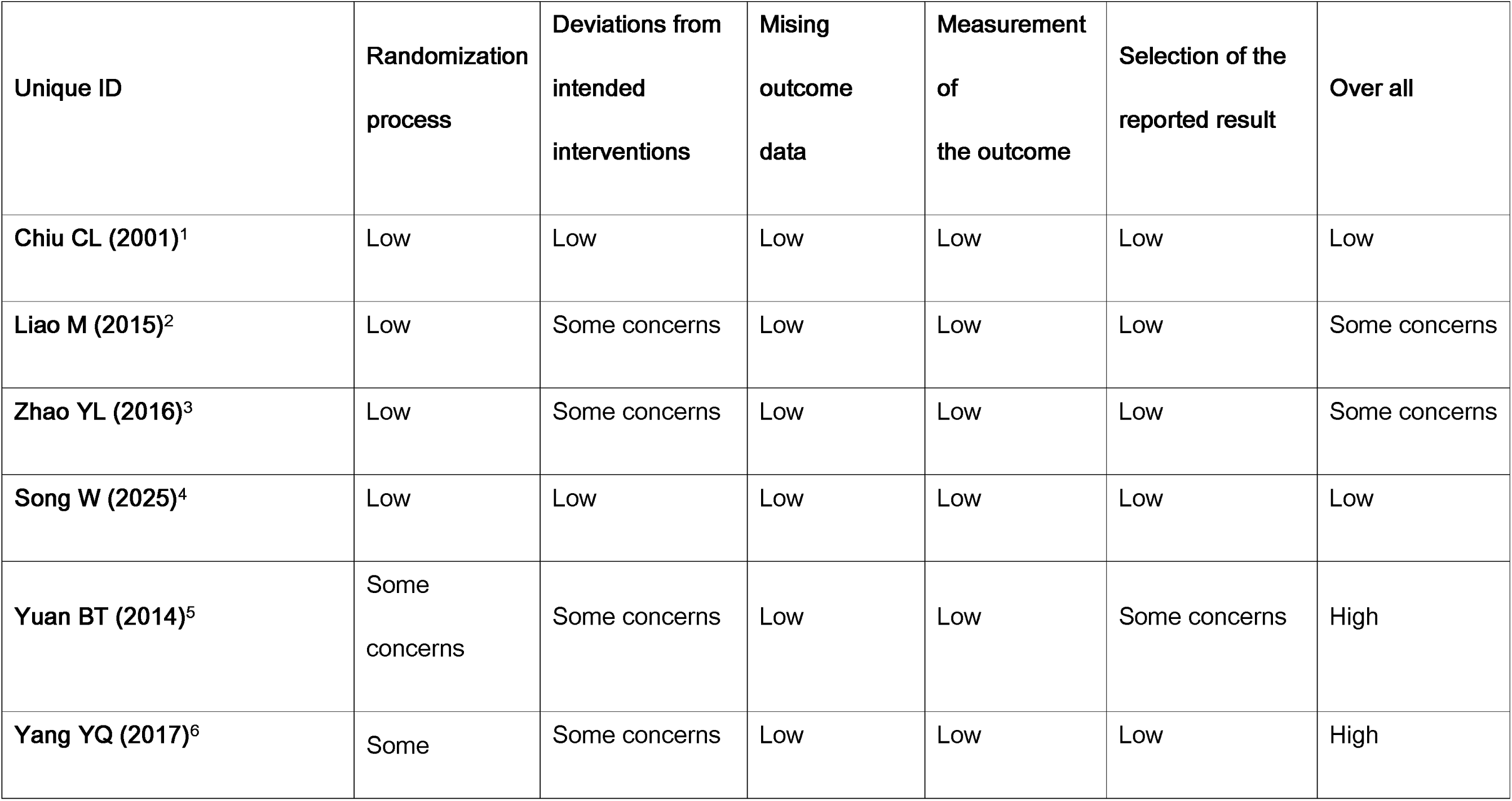
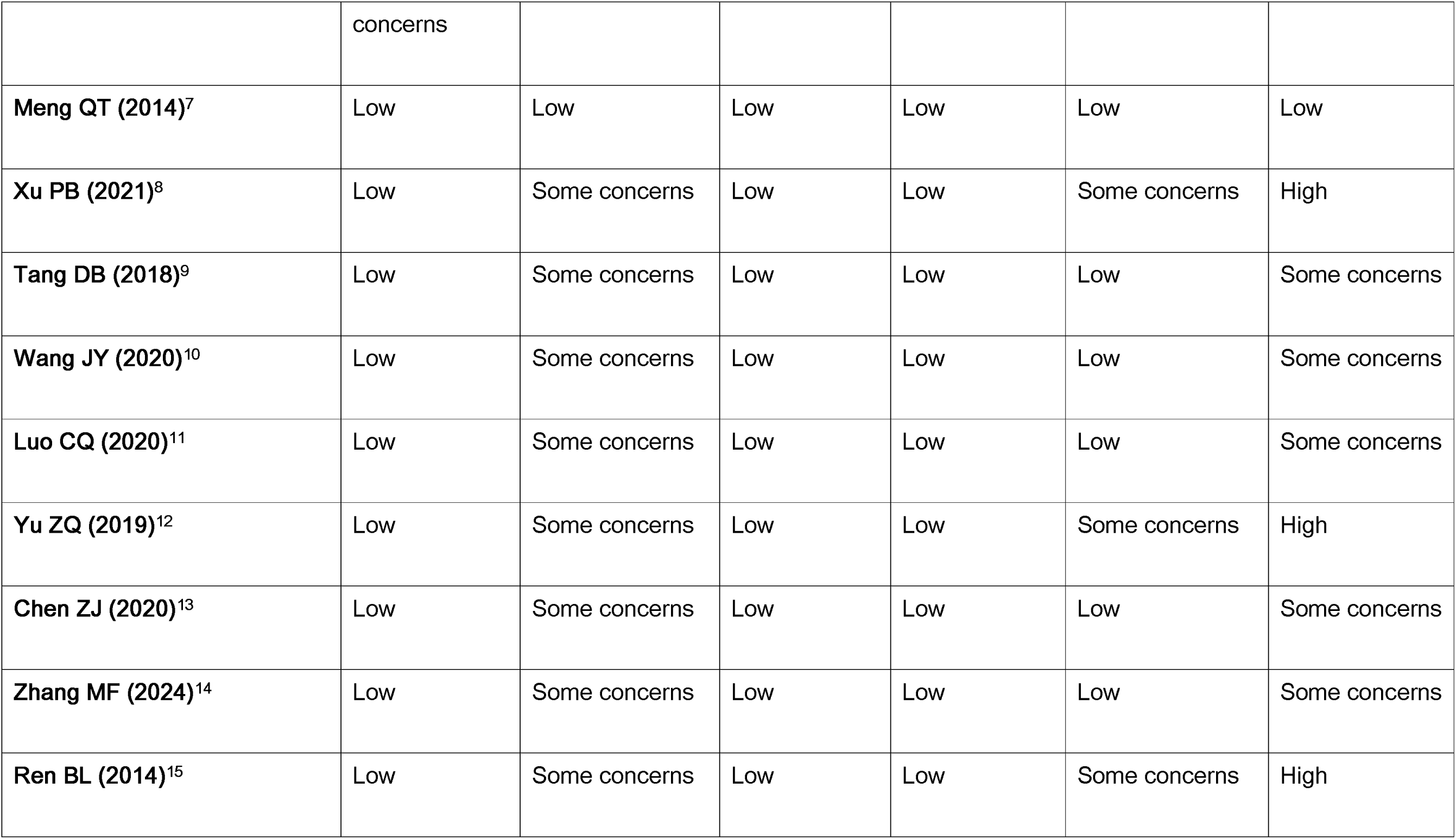
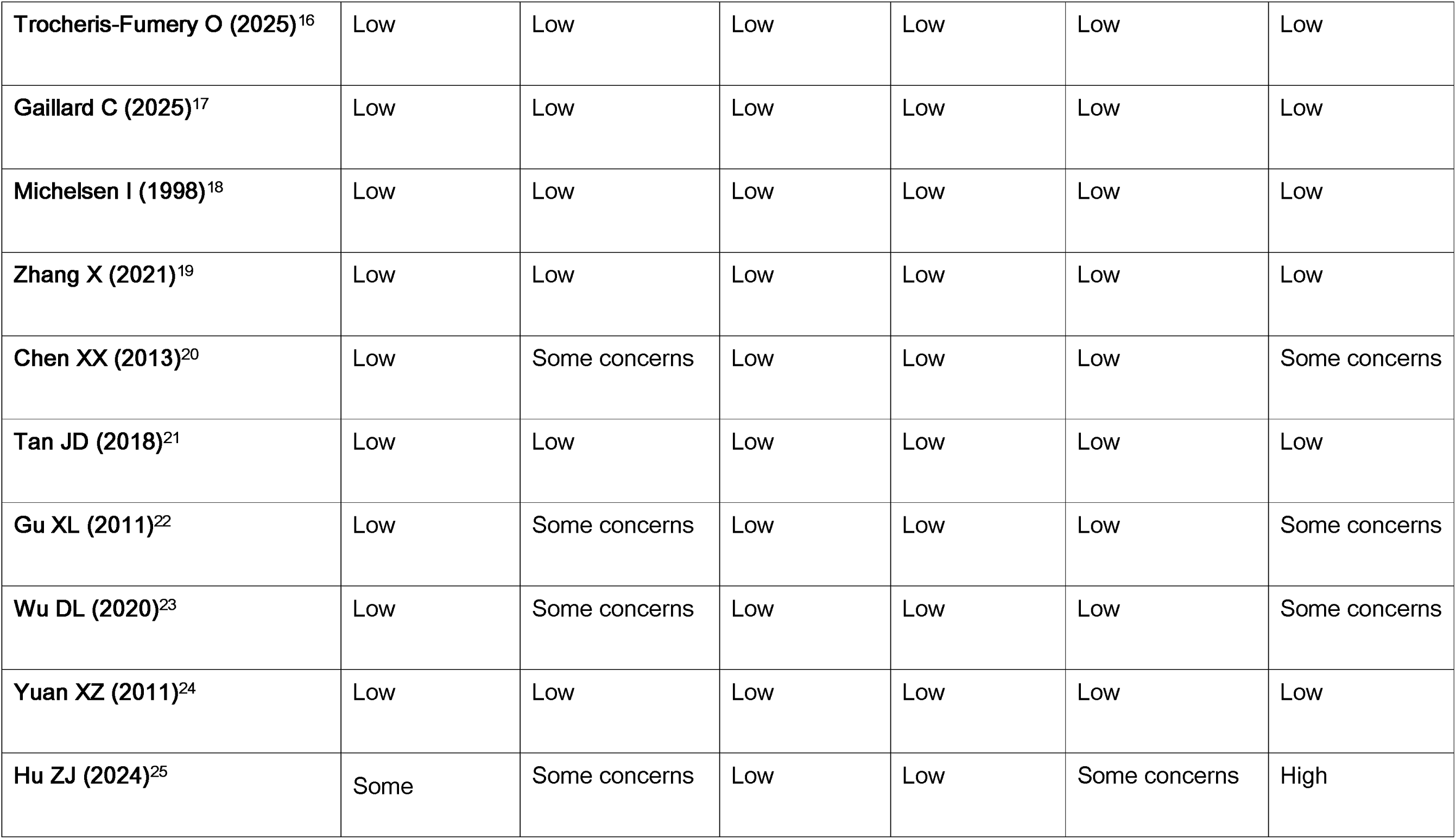
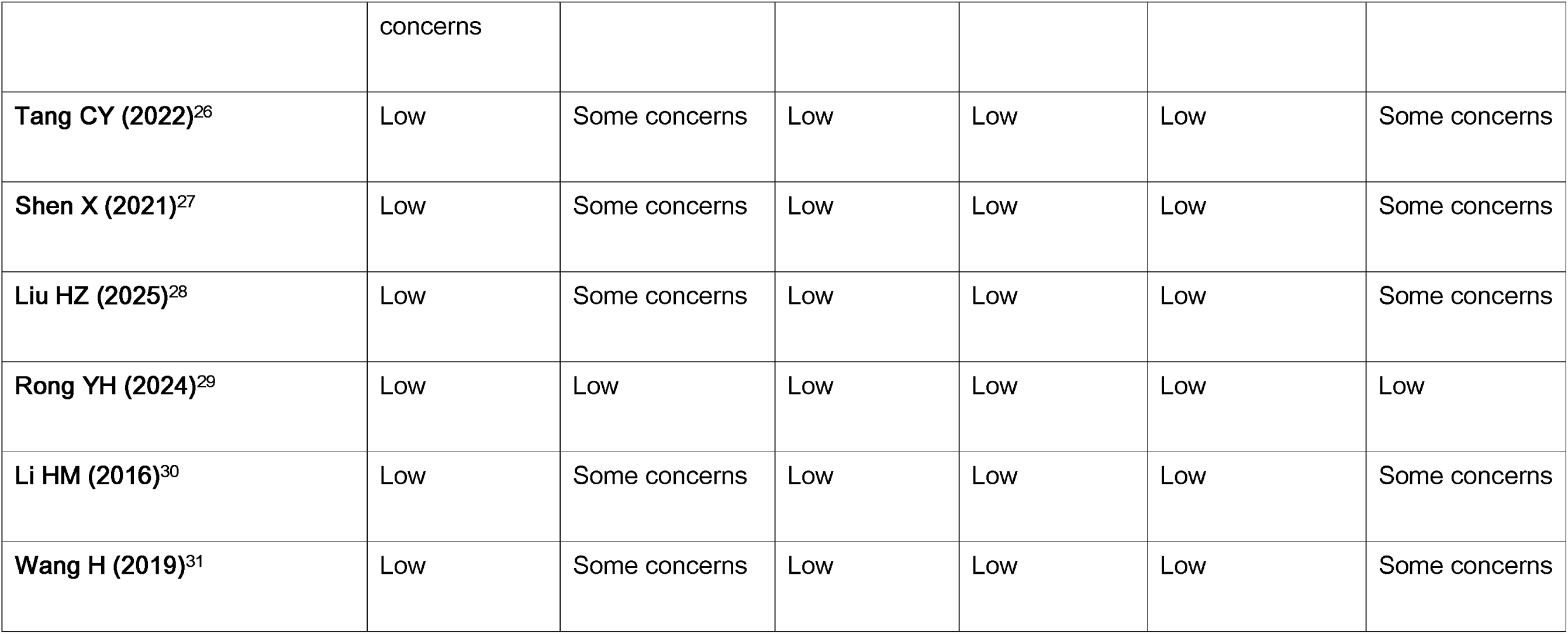
Study level risk of bias assessment using Cochrane risk of bias tool 2.0 for assessing risk of bias of randomized clinical trials.

## Appendix 5: Evaluation of inconsistency and heterogeneity

**Table S5.1:** Global consistency.

| Clinical outcome | Chi square | P value |
| --- | --- | --- |
| Incidence of Hypotension | 1.36 | 0.7158 |
| Post-induction MAP | 0.13 | 0.7134 |
| Post-induction SBP | 13.69 | 0.0002 |
| Post-induction HR | 2.05 | 0.1527 |

**Table S5.2:** Side-splitting of Incidence of Hypotension. Inconsistency test between direct and indirect treatment comparisons in mixed treatment comparison.

| Comparison | Direct |  | Indirect |  | Difference |  |  | Tau |
| --- | --- | --- | --- | --- | --- | --- | --- | --- |
|  | Coef. | Std. Err. | Coef. | Std. Err. | Coef. | Std. Err. | P>\|z\| |  |
| Ephedrine v.s. Methoxamine | -0.4294366 | 1.172893 | -0.3772262 | 0.4593031 | -0.0522104 | 1.259749 | 0.967 | 0.8067499 |
| Ephedrine v.s. Norepinephrine | -0.8108491 | 0.5655673 | 0.0500031 | 0.5075577 | -0.8608522 | 0.7599264 | 0.257 | 0.7841246 |
| Ephedrine v.s. Placebo | 1.231978 | 0.3459652 | 0.470527 | 0.6515213 | 0.7614512 | 0.7291009 | 0.296 | 0.7846384 |
| Metaraminol v.s. Placebo* | 1.646833 | 1.057207 | 0.4944947 | 446.7789 | 1.152339 | 446.7819 | 0.998 | 0.7901339 |
| Methoxamine v.s. Placebo* | 1.457041 | 0.3144698 | 1.815645 | 2.00072 | -0.3586038 | 2.007111 | 0.858 | 0.8106472 |
| Norepinephrine v.s. Placebo | 1.188599 | 0.3818584 | 2.049449 | 0.6616006 | -0.8608508 | 0.7599286 | 0.257 | 0.784125 |
| Phenylephrine v.s. Placebo* | 1.791759 | 1.308044 | 0.3495623 | 632.582 | 1.442197 | 632.585 | 0.998 | 0.790134 |

**Table S5.3:** Side-splitting of Post-induction MAP. Inconsistency test between direct and indirect treatment comparisons in mixed treatment comparison.

| Comparison | Direct | Indirect | Difference | Tau |
| --- | --- | --- | --- | --- |

|  | Coef. | Std. Err. | Coef. | Std. Err. | Coef. | Std. Err. | P>\|z\| |  |
| --- | --- | --- | --- | --- | --- | --- | --- | --- |
| Ephedrine v.s. Norepinephrine | 3.3 | 7.696074 | -0.3626753 | 6.330542 | 3.662675 | 9.965205 | 0.713 | 7.603338 |
| Ephedrine v.s. Placebo | -13.74168 | 2.851638 | -10.13696 | 9.483256 | -3.604714 | 9.903376 | 0.716 | 7.600443 |
| Metaraminol v.s. Placebo* | -9.45311 | 5.496056 | -17.40209 | 447.3448 | 7.948985 | 447.3851 | 0.986 | 7.391095 |
| Methoxamine v.s. Placebo* | -15.13456 | 2.404342 | -11.71715 | 200.1245 | -3.417416 | 200.1406 | 0.986 | 7.391152 |
| Norepinephrine v.s. Placebo | -13.37985 | 5.651777 | -17.042 | 8.206414 | 3.662155 | 9.965277 | 0.713 | 7.603334 |
| Phenylephrine v.s. Placebo* | -18.9 | 7.522599 | -7.953292 | 632.612 | -10.94671 | 632.6598 | 0.986 | 7.391042 |

**Table S5.4:** Side-splitting of Post-induction SBP. Inconsistency test between direct and indirect treatment comparisons in mixed treatment comparison.

| Comparison | Direct |  | Indirect |  | Difference |  |  | Tau |
| --- | --- | --- | --- | --- | --- | --- | --- | --- |
|  | Coef. | Std. Err. | Coef. | Std. Err. | Coef. | Std. Err. | P>\|z\| |  |
| Ephedrine v.s. Placebo* | -16.96745 | 4.256012 | -10.98468 | 74.58102 | -5.982775 | 74.70234 | 0.936 | 10.51063 |
| Metaraminol v.s. Placebo* | -11.9305 | 7.679994 | -21.96901 | 447.4843 | 10.03851 | 447.5587 | 0.982 | 10.50949 |
| Methoxamine v.s. Placebo* | -7.488474 | 3.354372 | -26.4451 | 190.9613 | 18.95663 | 190.997 | 0.921 | 10.51023 |
| Norepinephrine v.s. Placebo* | -18.2979 | 4.946119 | -15.59559 | 283.0972 | -2.702311 | 283.1488 | 0.992 | 10.50955 |

**Table S5.5:** Side-splitting of Post-induction HR. Inconsistency test between direct and indirect treatment comparisons in mixed treatment comparison.

| Comparison | Direct |  | Indirect |  | Difference |  |  | Tau |
| --- | --- | --- | --- | --- | --- | --- | --- | --- |
|  | Coef. | Std. Err. | Coef. | Std. Err. | Coef. | Std. Err. | P>\|z\| |  |
| Ephedrine v.s. Placebo* | -10.00726 | 1.545057 | 1.94992 | 63.25288 | -11.95718 | 63.27173 | 0.85 | 4.301057 |
| Metaraminol v.s. Placebo* | -0.6504399 | 3.339455 | -19.35058 | 447.2448 | 18.70014 | 447.2616 | 0.967 | 4.300936 |
| Methoxamine v.s. Placebo* | 2.963254 | 1.216572 | -22.97273 | 163.3178 | 25.93598 | 163.3238 | 0.874 | 4.300998 |
| Norepinephrine v.s. Placebo* | -0.2428403 | 1.815016 | -19.76062 | 239.1271 | 19.51778 | 239.1367 | 0.935 | 4.300965 |
| Phenylephrine v.s. Placebo* | 7.3 | 5.102753 | -27.30108 | 632.5403 | 34.60108 | 632.5728 | 0.956 | 4.30094 |

## Appendix 6: Network maps and forest plots of outcomes

**Figure S6.1.**
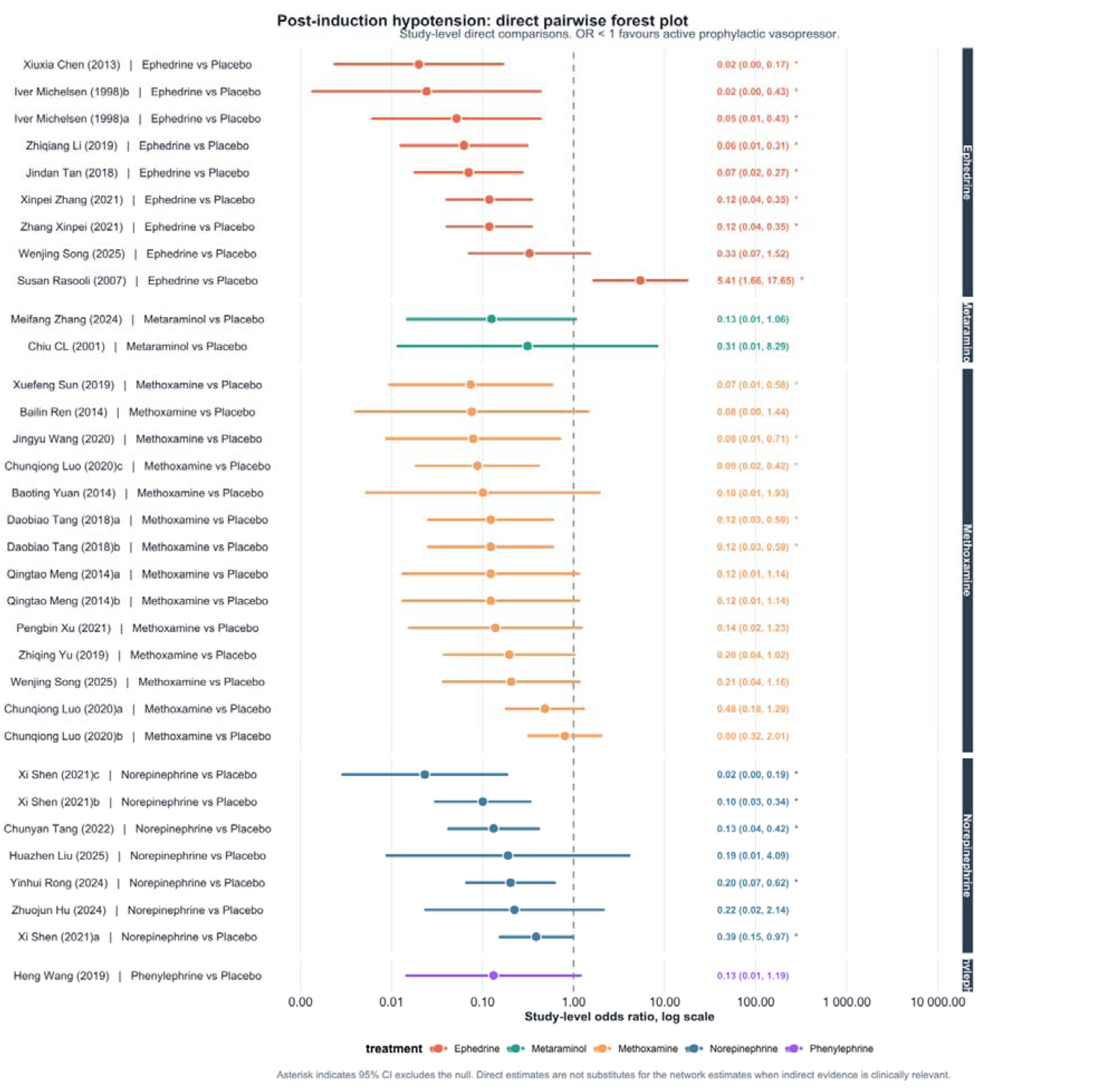
Direct pairwise forest plot for post-induction hypotension. Study-level odds ratios compare active prophylactic vasopressors with placebo/control; odds ratios less than 1 favour active prophylaxis.

**Figure S6.2.**
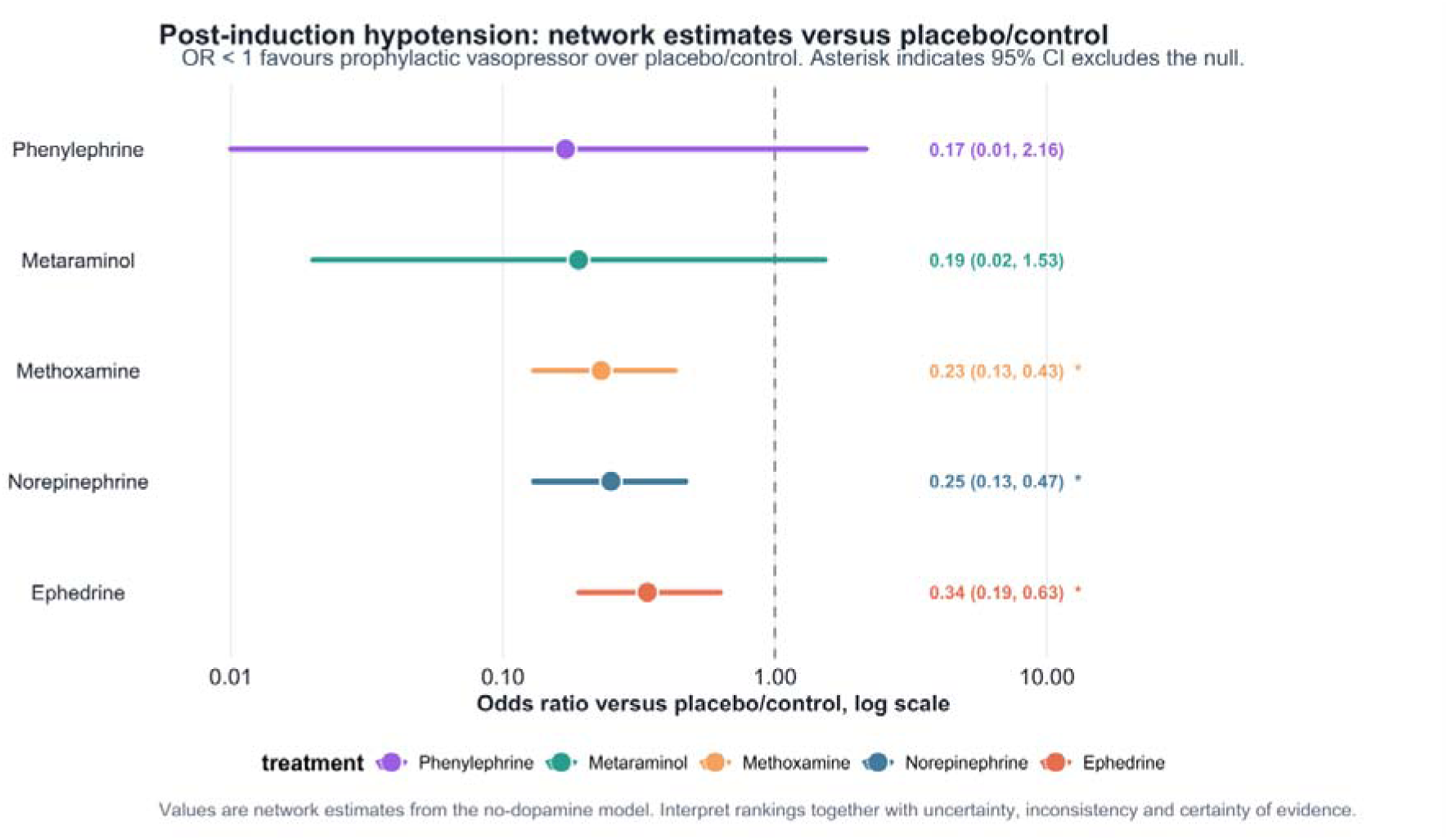
Network estimates versus placebo/control for post-induction hypotension. Odds ratios less than 1 favour active prophylaxis.

**Figure S6.3.**
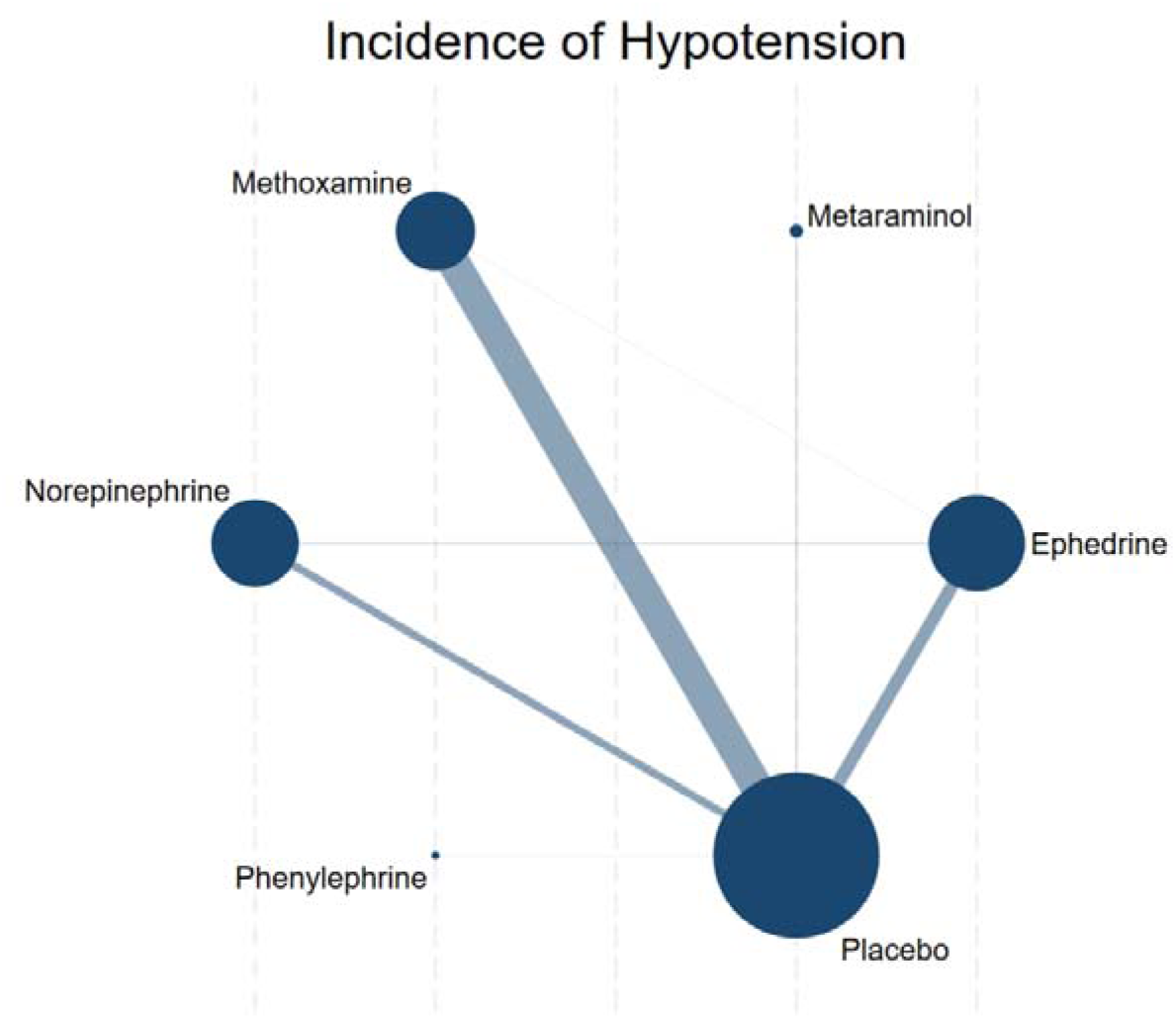
Network map of the effect on Incidence of Hypotension.The size of the nodes was proportional to the number of participants included in the trial, and the thickness of lines between the interventions relates to the number of studies for that comparison.

**Figure S6.4.**
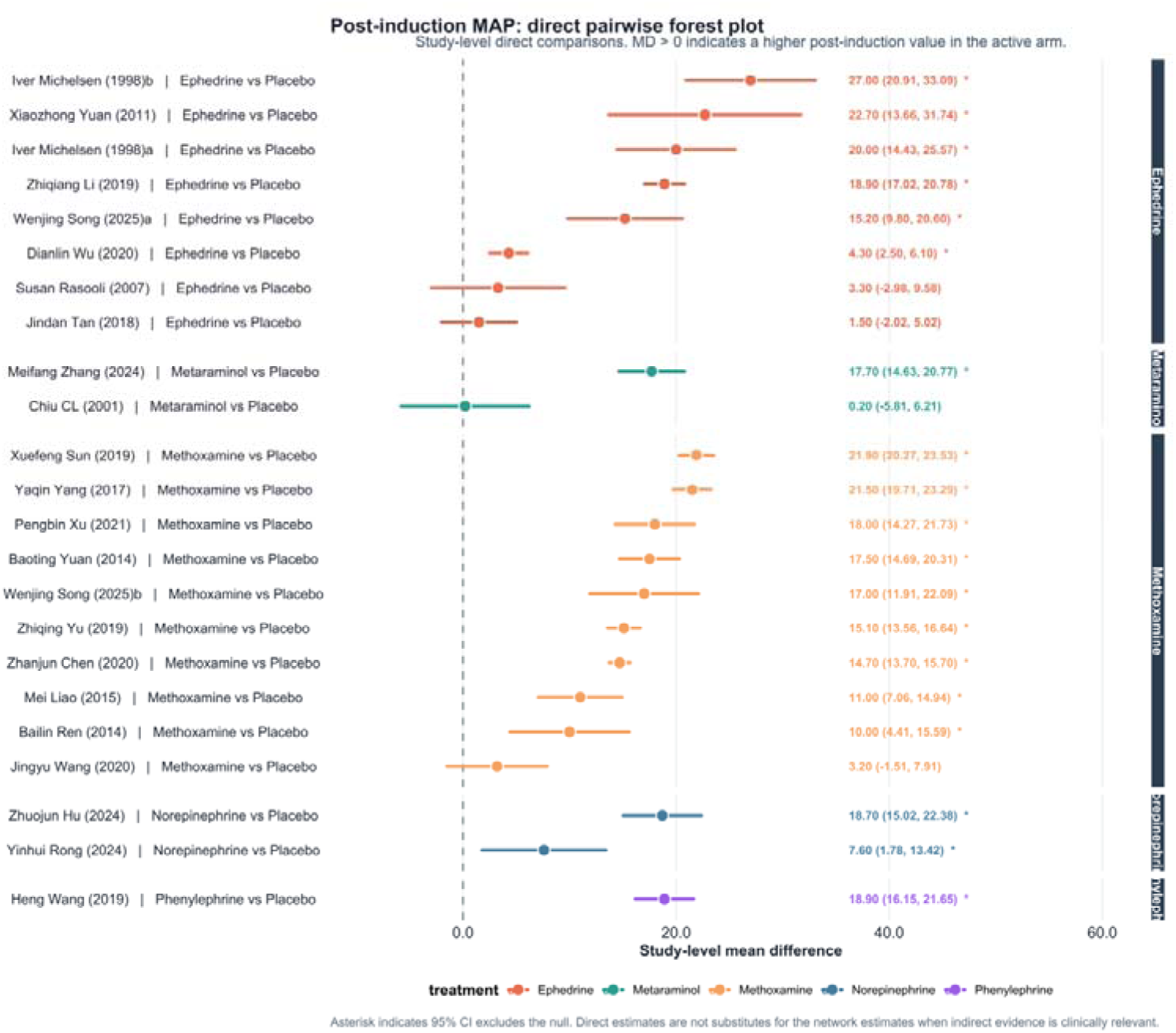
Direct pairwise forest plot for post-induction mean arterial pressure. Positive mean differences indicate higher post-induction MAP in the active arm.

**Figure S6.5.**
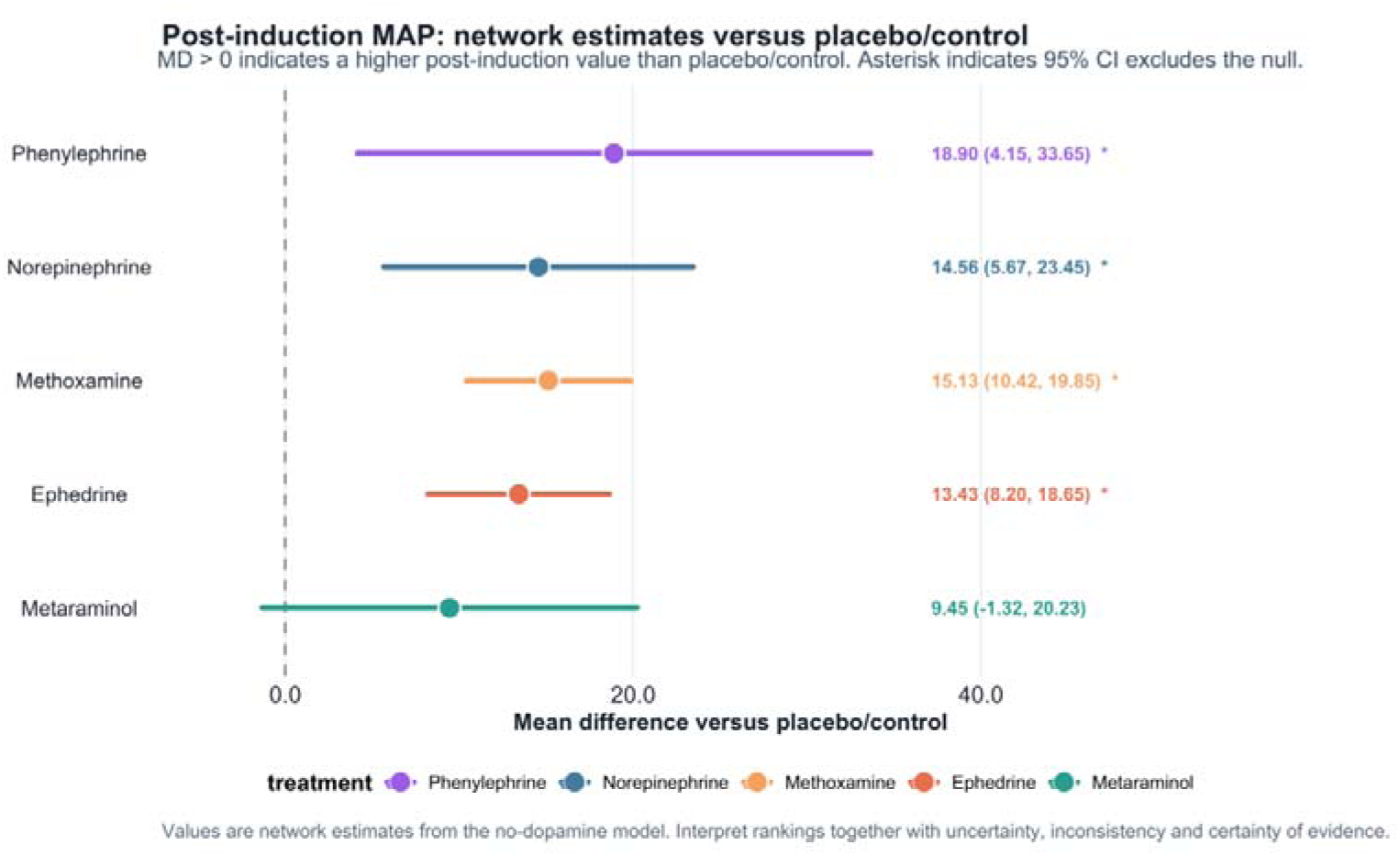
Network estimates versus placebo/control for post-induction mean arterial pressure. Positive mean differences indicate higher MAP with active prophylaxis.

**Figure S6.6.**
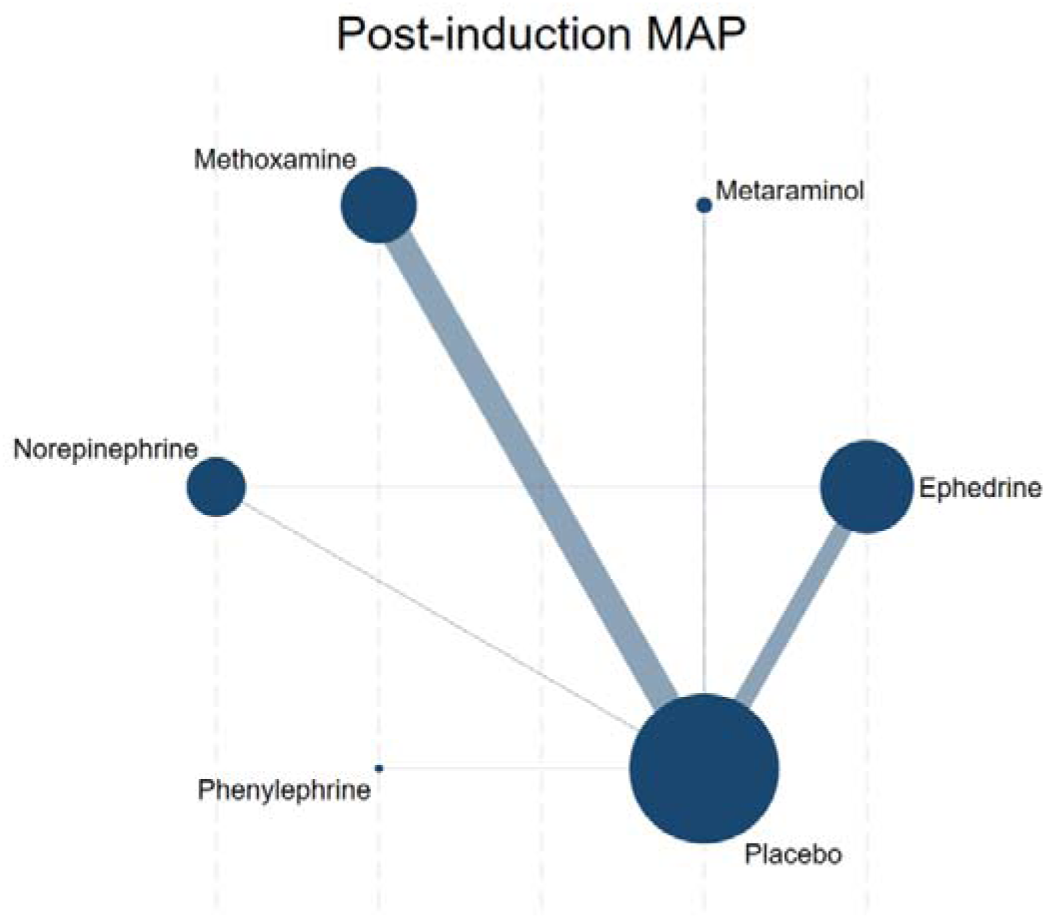
Network map of the effect on Incidence of Post-induction MAP.The size of the nodes was proportional to the number of participants included in the trial, and the thickness of lines between the interventions relates to the number of studies for that comparison.

**Figure S6.5.**
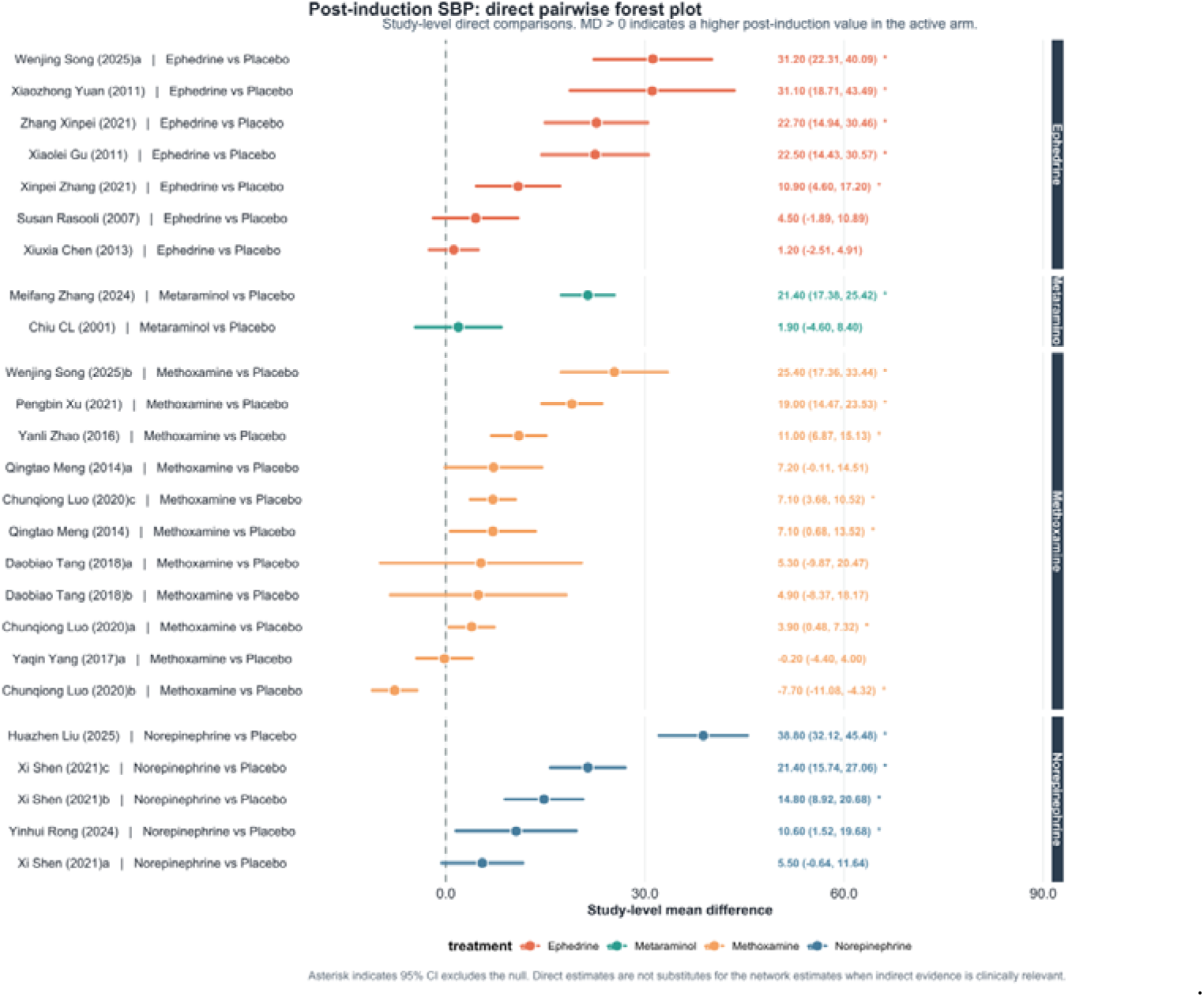
Direct pairwise forest plot for post-induction systolic arterial pressure. Positive mean differences indicate higher post-induction SBP in the active arm.

**Figure S6.6.**
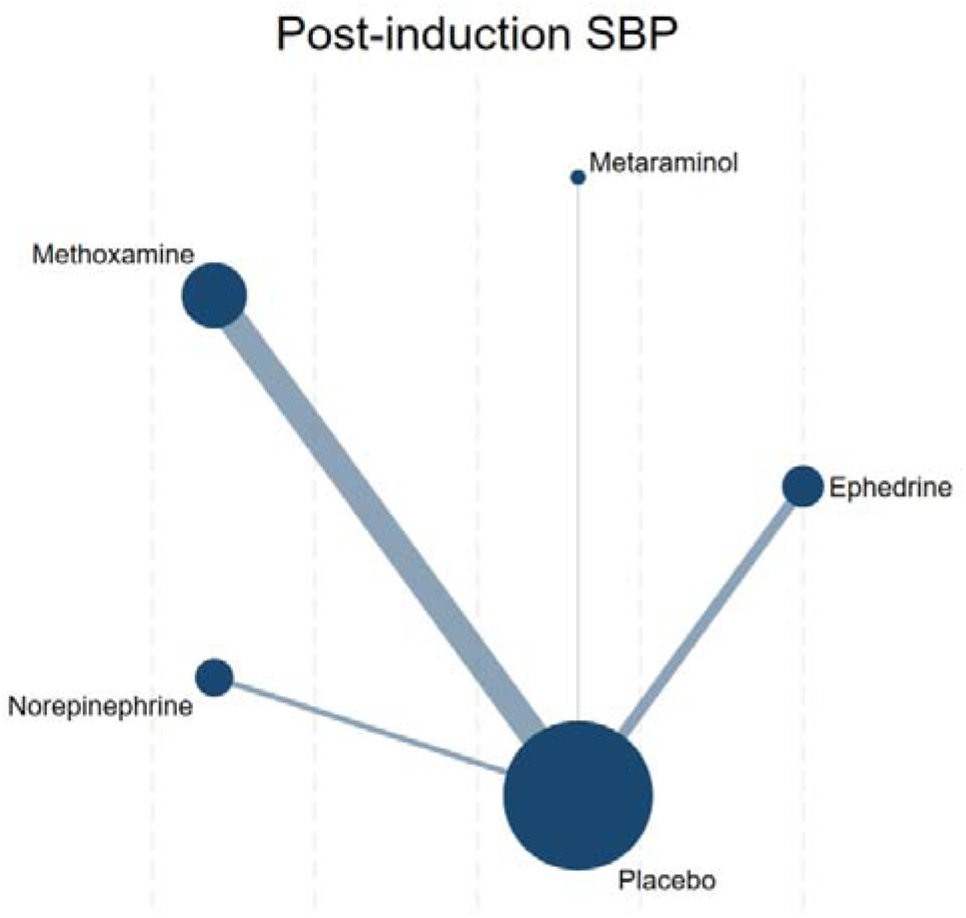
Network map of the effect on Incidence of Post-induction SBP.The size of the nodes was proportional to the number of participants included in the trial, and the thickness of lines between the interventions relates to the number of studies for that comparison.

**Figure S6.7.**
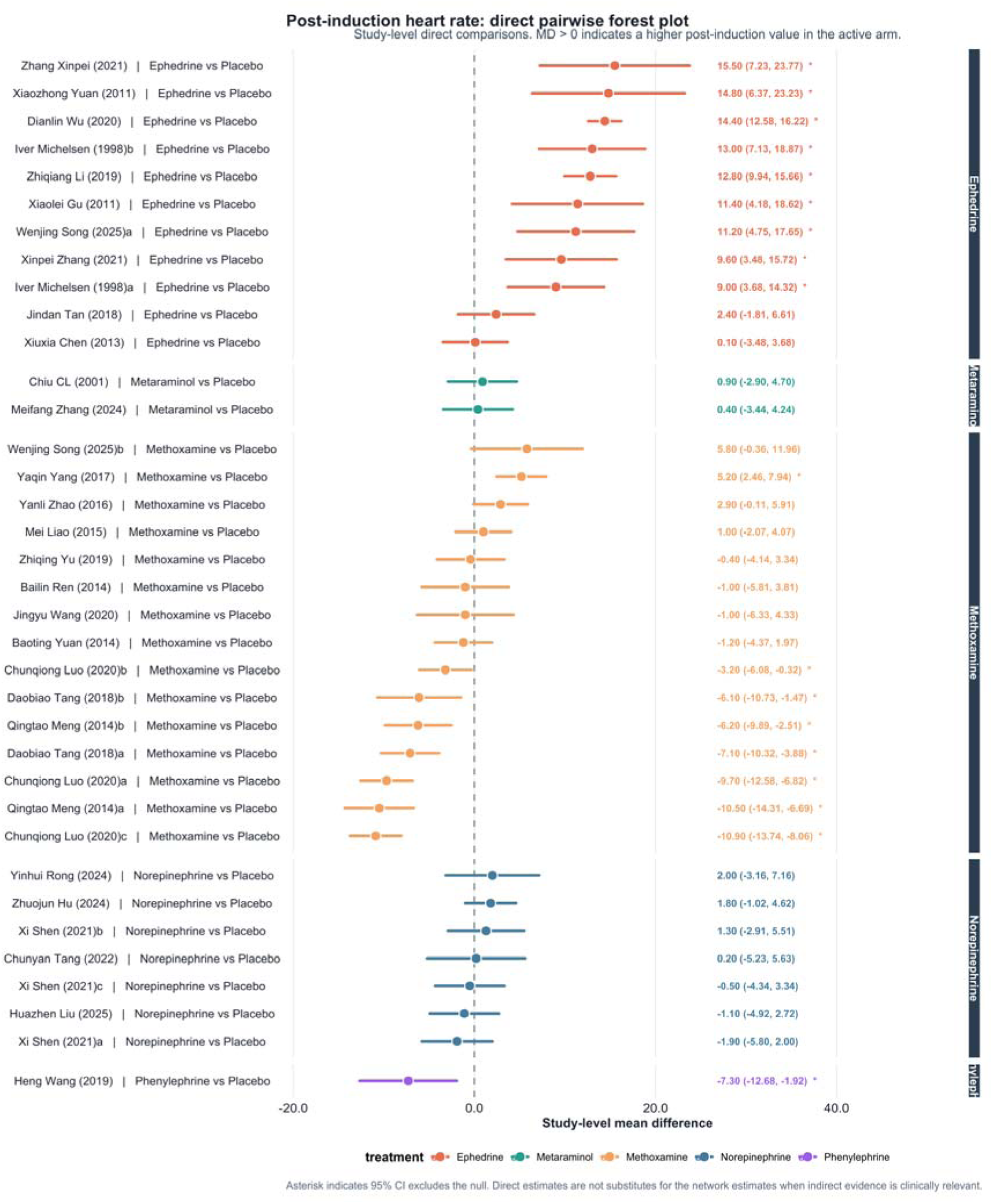
Direct pairwise forest plot for post-induction heart rate. Positive mean differences indicate higher post-induction HR in the active arm.

**Figure S6.8.**
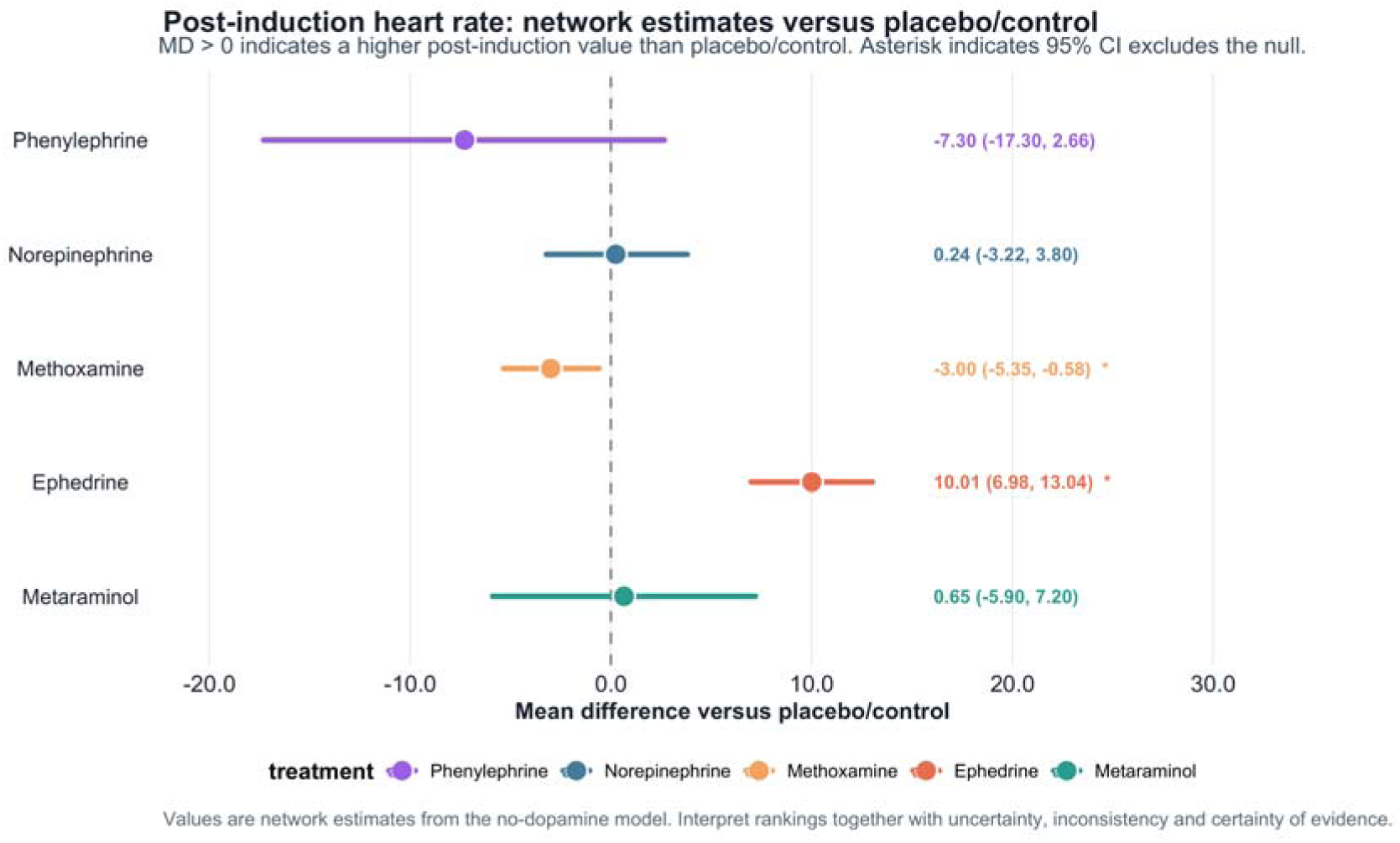
Network estimates versus placebo/control for post-induction heart rate. Positive mean differences indicate higher HR with active prophylaxis.

**Figure S6.6.**
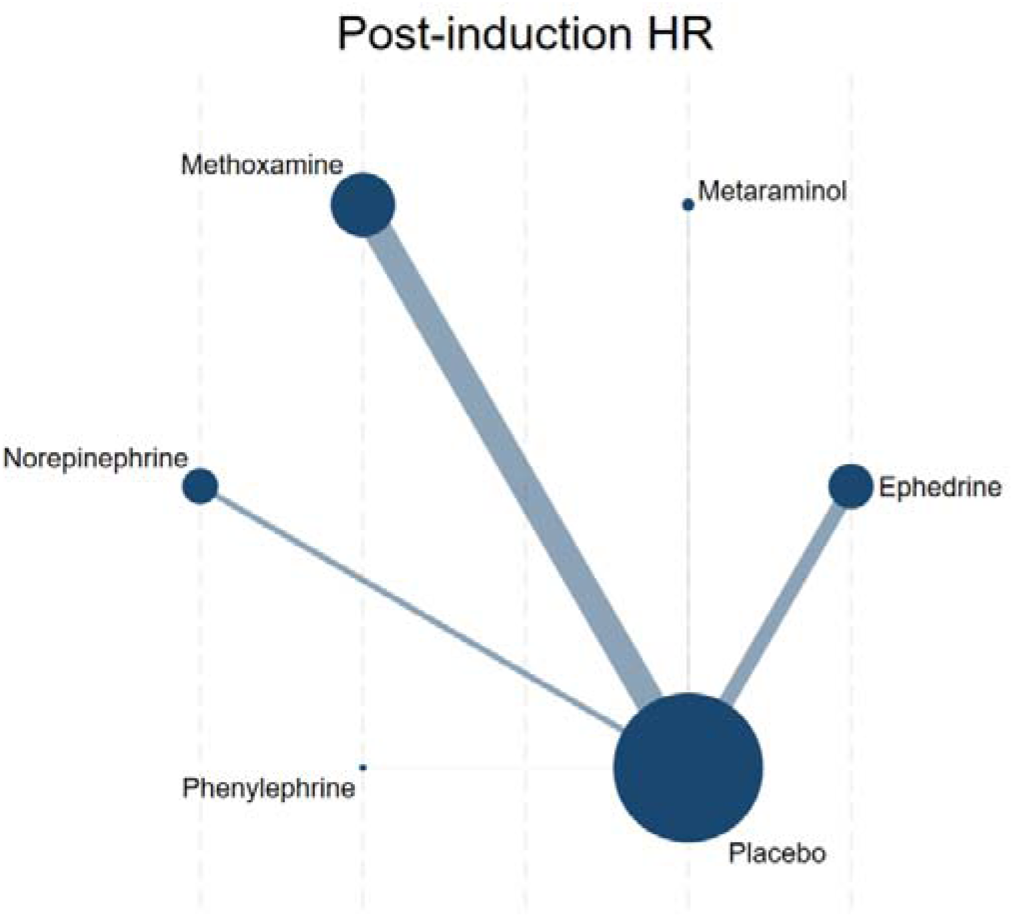
Network map of the effect on Incidence of Post-induction HR.The size of the nodes was proportional to the number of participants included in the trial, and the thickness of lines between the interventions relates to the number of studies for that comparison.

## Appendix 7: SUCRA and cumulative probability plots

**Figure S7.1.**
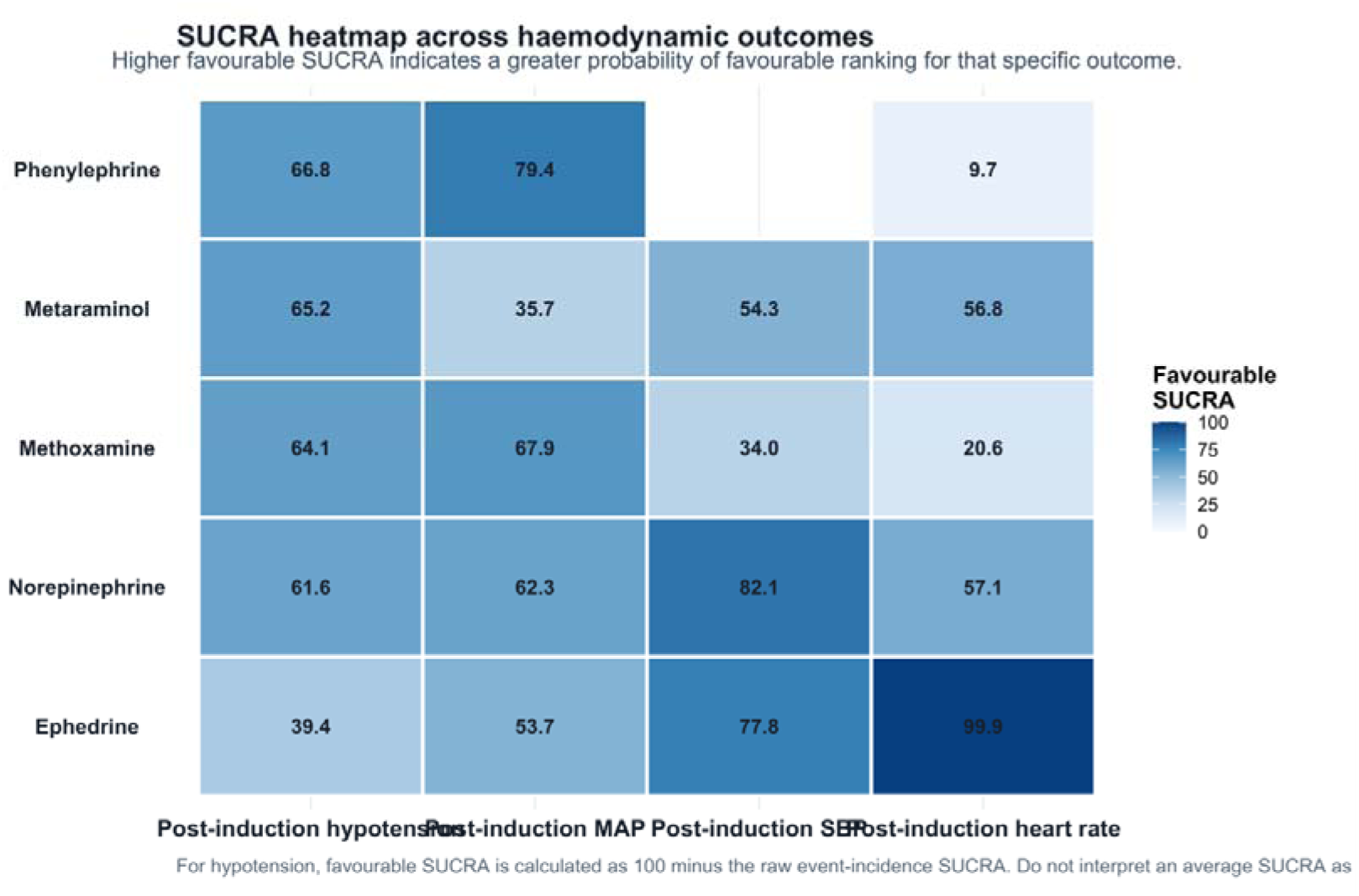
SUCRA heatmap across haemodynamic outcomes.

**Figure S7.2.**
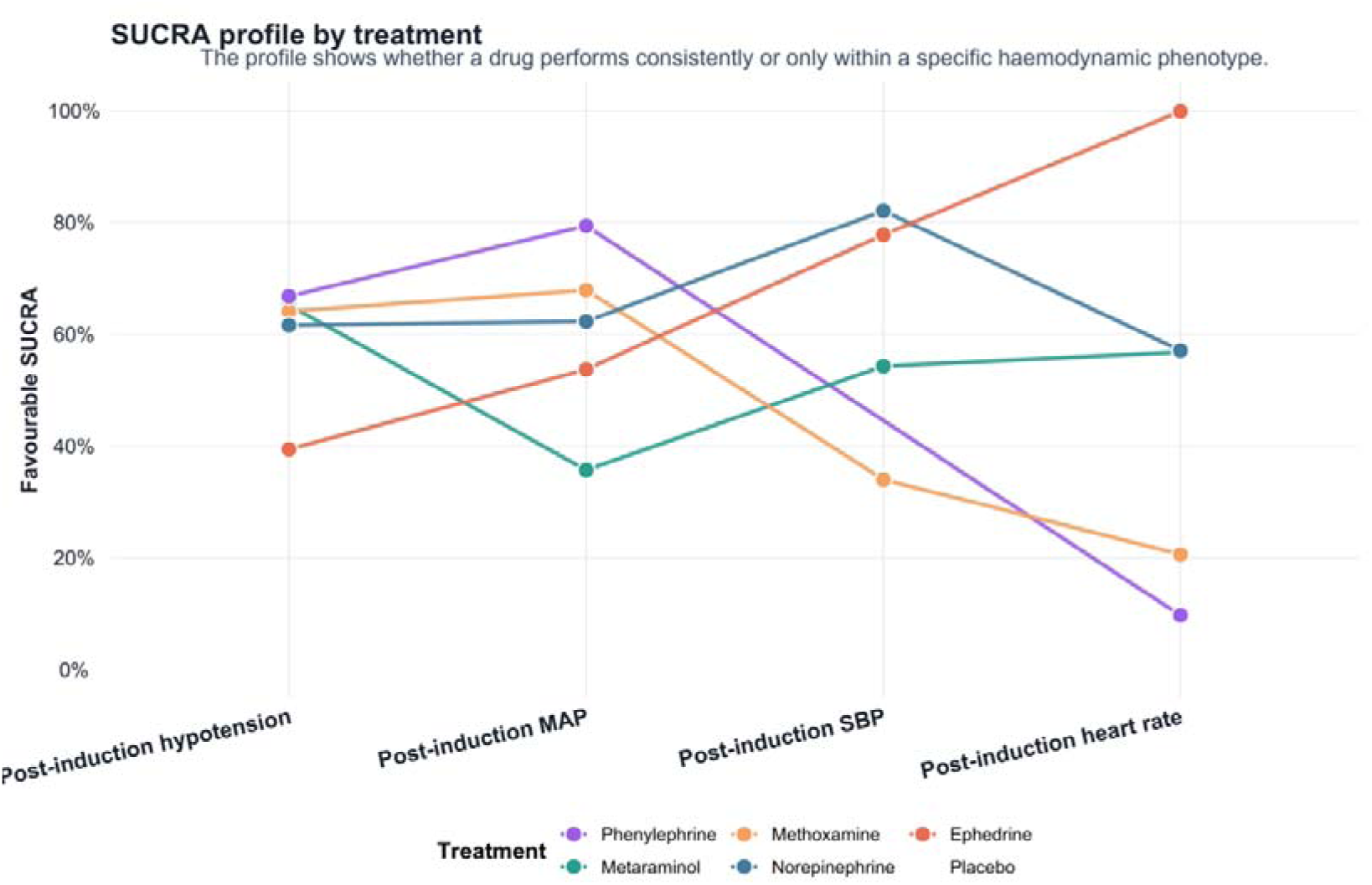
SUCRA profile by treatment across post-induction hypotension, MAP, SBP and HR. The profile emphasises outcome-specific haemodynamic patterns rather than a single overall hierarchy.

**Figure S7.3.**
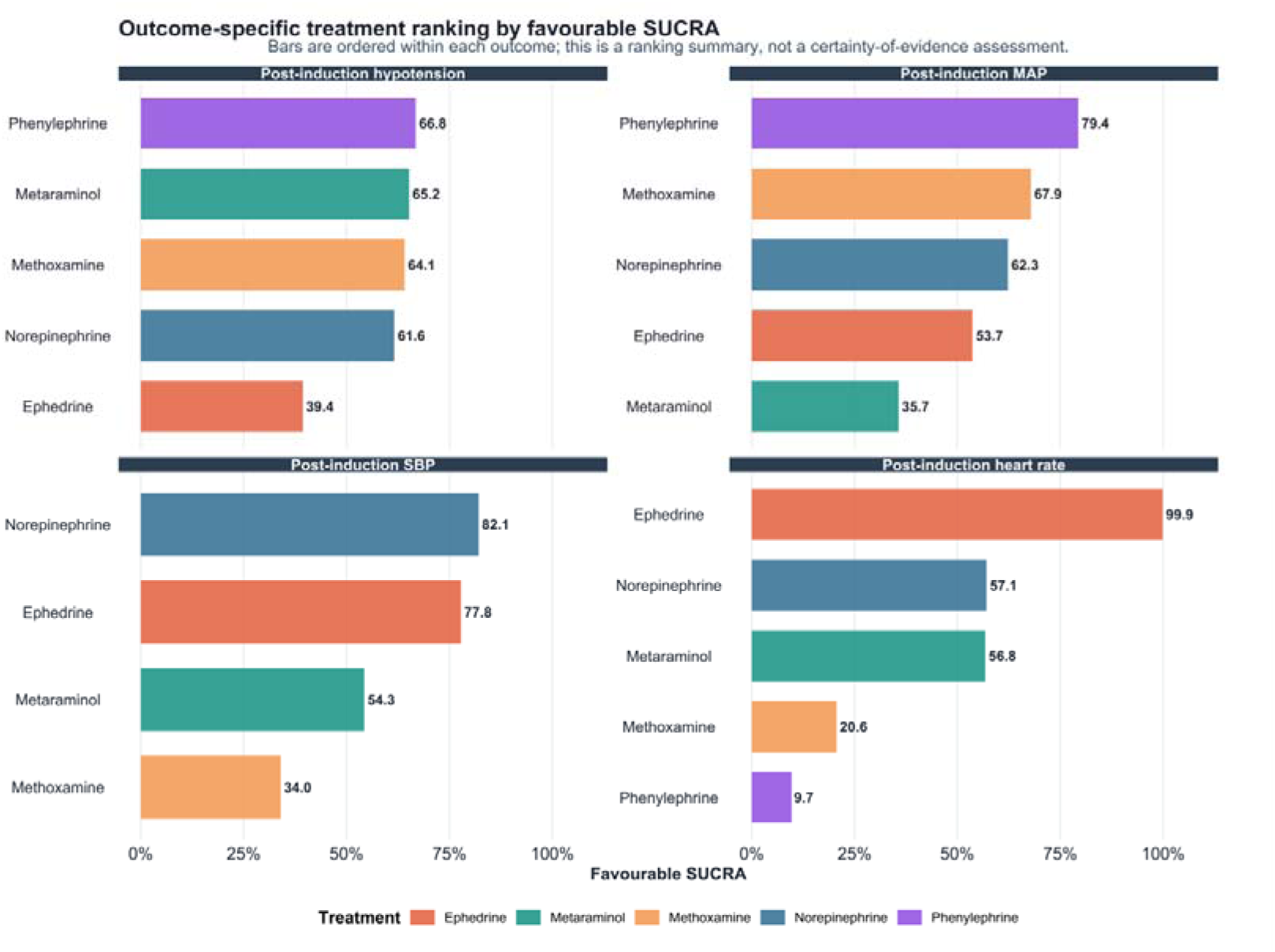
Outcome-specific treatment ranking by favourable SUCRA. Bars are ordered within each outcome and should be interpreted alongside effect estimates, uncertainty, inconsistency and certainty of evidence.

**Figure S7.4.**
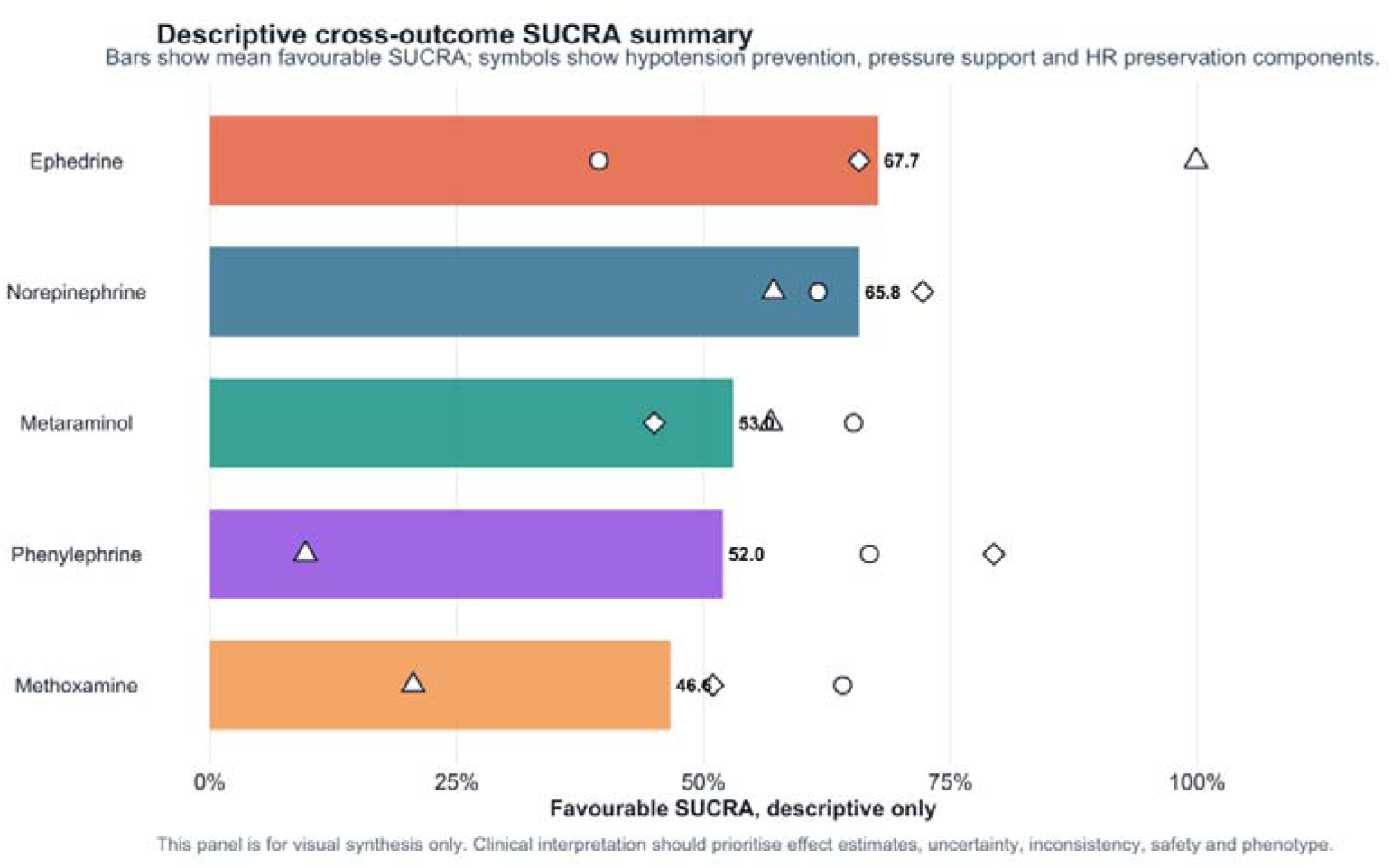
Descriptive cross-outcome SUCRA summary. This visual synthesis is descriptive only and should not be interpreted as proof of global superiority or safety.

**Table S7.1:**
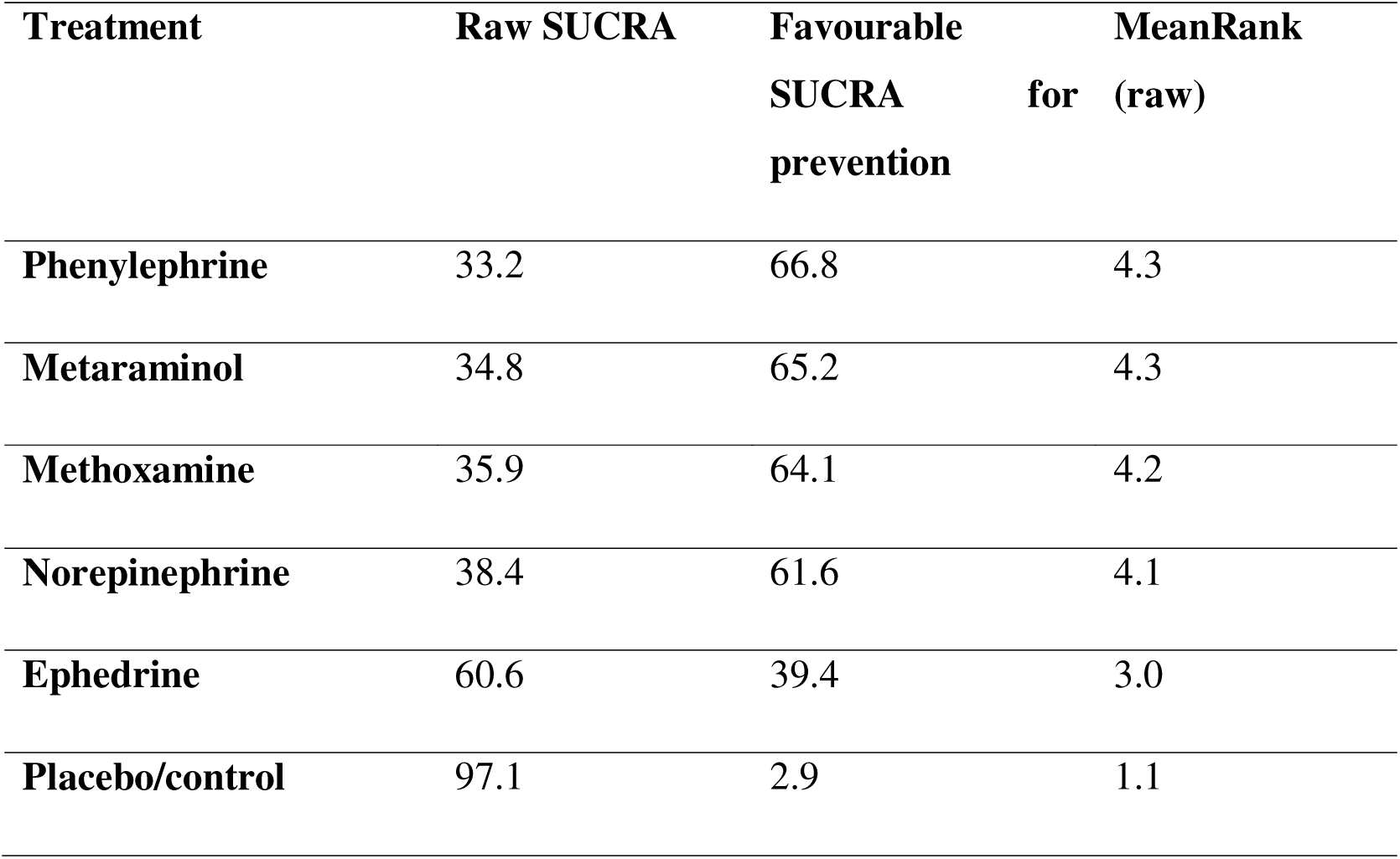
SUCRA and ranking metrics of five active vasoactive agents and placebo/control for the incidence of hypotension.

**Table S7.2:** SUCRA and ranking metrics of five active vasoactive agents and placebo/control for post-induction MAP.

| **Treatment** | **SUCRA** | **PrBest** | **MeanRank** |
| --- | --- | --- | --- |
| **Phenylephrine** | 79.4 | 57.5 | 2.0 |
| **Methoxamine** | 67.9 | 15.2 | 2.6 |
| **Norepinephrine** | 62.3 | 17.7 | 2.9 |
| **Ephedrine** | 53.7 | 4.9 | 3.3 |
| **Metaraminol** | 35.7 | 4.7 | 4.2 |
| **Placebo/control** | 1.1 | 0.0 | 5.9 |

**Table S7.3:** SUCRA and ranking metrics of estimable active vasoactive agents and placebo/control for post-induction SBP.

| **Treatment** | **SUCRA** | **PrBest** | **MeanRank** |
| --- | --- | --- | --- |
| **Norepinephrine** | 82.1 | 48.7 | 1.7 |
| **Ephedrine** | 77.8 | 34.6 | 1.9 |
| **Metaraminol** | 54.3 | 16.5 | 2.8 |
| **Methoxamine** | 34.0 | 0.2 | 3.6 |
| **Placebo/control** | 1.8 | 0.0 | 4.9 |

**Table S7.4:** SUCRA and ranking metrics of five active vasoactive agents and placebo/control for post-induction HR.

| **Treatment** | **SUCRA** | **PrBest** | **MeanRank** |
| --- | --- | --- | --- |
| **Ephedrine** | 99.9 | 99.4 | 1.0 |
| **Norepinephrine** | 57.1 | 0.0 | 3.1 |
| **Metaraminol** | 56.8 | 0.5 | 3.2 |
| **Placebo/control** | 55.9 | 0.0 | 3.2 |
| **Methoxamine** | 20.6 | 0.0 | 5.0 |
| **Phenylephrine** | 9.7 | 0.0 | 5.5 |

## Appendix 8: league table of Summary Estimates for Prophylactic Vasopressors On Preventing Post-induction Hypotension in the Elderly Derived from Network Meta-analysis of 31 Trials

**Table S8.1:**
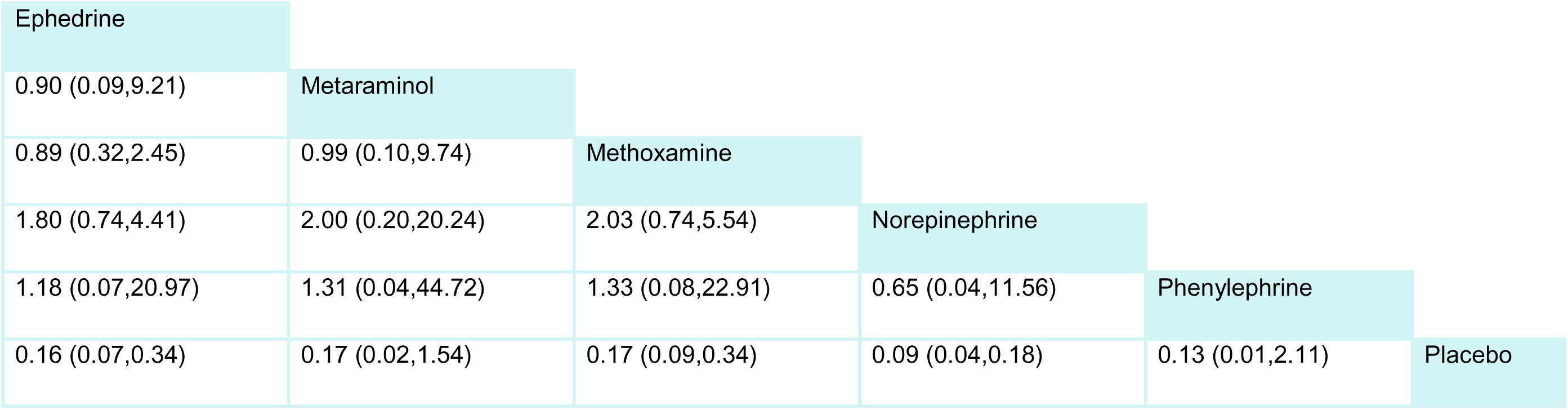
Incidence of Hypotension. The columns represent the comparison of the row drug class to the column drug class. The rows represent the comparison of the row drug class to the column drug class. The effect estimates are expressed as odds ratio and 95% confidence interval. For example, the odds ratio in Incidence of Hypotension for Ephedrine compared to Metaraminol is 0.90 (95% confidence interval 0.09 to 9.21). OR <1 favors the drug in the column, and OR>1 favors the drug in the row.

**Table S8.2:** Post-induction MAP. The columns represent the comparison of the row drug class to the column drug class. The rows represent the comparison of the row drug class to the column drug class. The effect estimates are expressed as mean difference and 95% confidence interval. For example, the mean difference in Post-induction MAP for Ephedrine compared to Metaraminol is 4.37 (95% confidence interval -7.25 to 16.00). Mean difference <0 favors the drug in the column, and mean difference >0 favors the drug in the row.

|  |  |  |  |  |  |
| --- | --- | --- | --- | --- | --- |
| Ephedrine |  |  |  |  |  |
| 4.37 (-7.25,16.00) | Metaraminol |  |  |  |  |
| -1.27 (-8.39,5.85) | -5.64 (-16.77,5.49) | Methoxamine |  |  |  |
| -0.90 (-8.53,6.73) | -5.27 (-18.05,7.50) | 0.37 (-8.51,9.25) | Norepinephrine |  |  |
| -5.02 (-20.00,9.96) | -9.39 (-26.65,7.87) | -3.75 (-18.36,10.86) | -4.12 (-20.01,11.78) | Phenylephrine |  |
| 13.88 (8.34,19.43) | 9.51 (-0.70,19.72) | 15.15 (10.70,19.60) | 14.78 (7.10,22.46) | 18.90 (4.98,32.82) | Placebo |

**Table S8.3:**
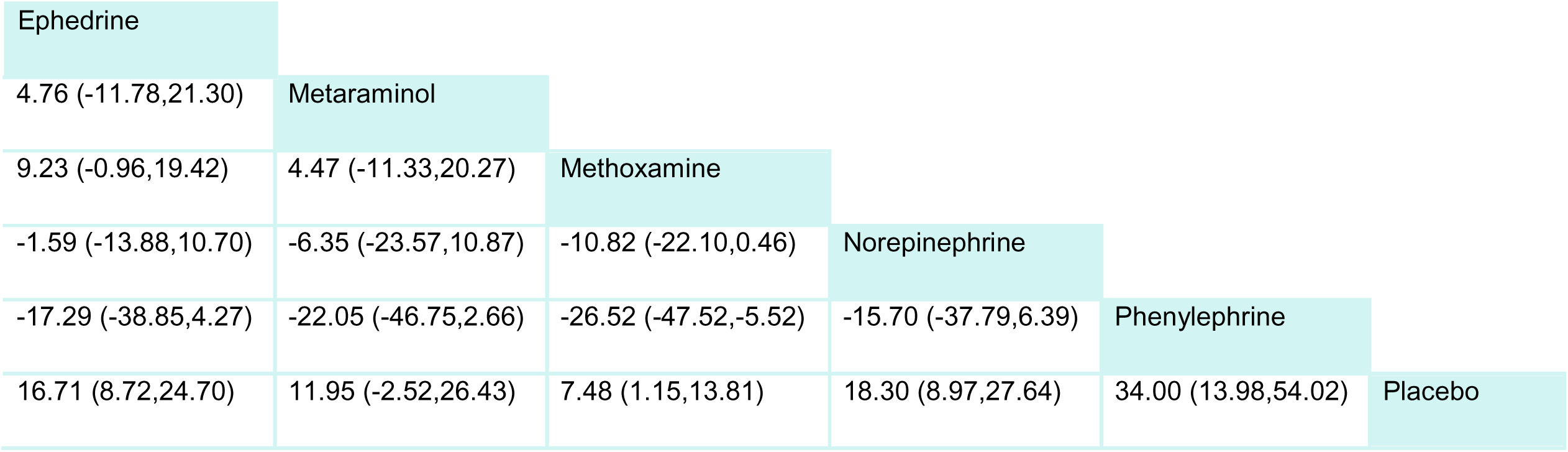
Post-induction SBP. The columns represent the comparison of the row drug class to the column drug class. The rows represent the comparison of the row drug class to the column drug class. The effect estimates are expressed as mean difference and 95% confidence interval. For example, the mean difference in Post-induction SBP for Ephedrine compared to Metaraminol is 4.76 (95% confidence interval -11.78 to 21.30). Mean difference <0 favors the drug in the column, and mean difference >0 favors the drug in the row.

**Table S8.4:**
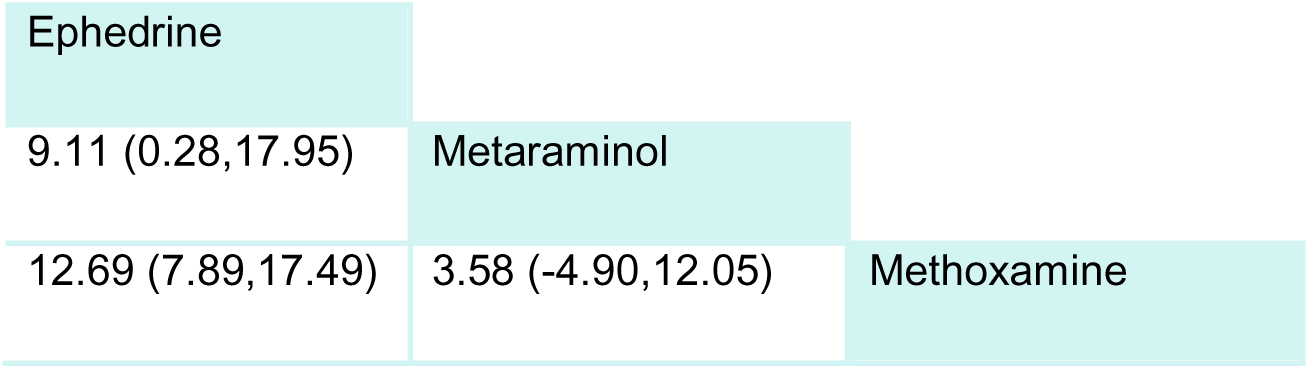
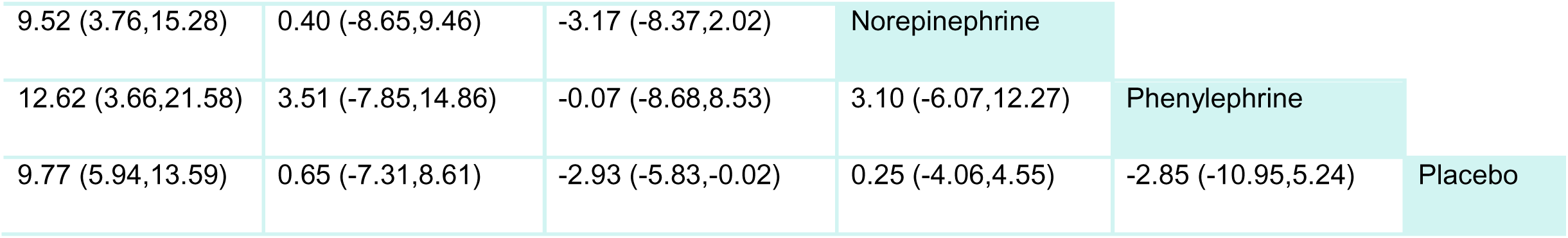
Post-induction HR. The columns represent the comparison of the row drug class to the column drug class. The rows represent the comparison of the row drug class to the column drug class. The effect estimates are expressed as mean difference and 95% confidence interval. For example, the mean difference in Post-induction HR for Ephedrine compared to Metaraminol is 9.11 (95% confidence interval 0.28 to 17.95). Mean difference <0 favors the drug in the column, and mean difference >0 favors the drug in the row.

|  |  |  |
| --- | --- | --- |
| Ephedrine |  |  |
| 9.11 (0.28,17.95) | Metaraminol |  |
| 12.69 (7.89,17.49) | 3.58 (-4.90,12.05) | Methoxamine |

|  |  |  |  |  |  |
| --- | --- | --- | --- | --- | --- |
| 9.52 (3.76,15.28) | 0.40 (-8.65,9.46) | -3.17 (-8.37,2.02) | Norepinephrine |  |  |
| 12.62 (3.66,21.58) | 3.51 (-7.85,14.86) | -0.07 (-8.68,8.53) | 3.10 (-6.07,12.27) | Phenylephrine |  |
| 9.77 (5.94,13.59) | 0.65 (-7.31,8.61) | -2.93 (-5.83,-0.02) | 0.25 (-4.06,4.55) | -2.85 (-10.95,5.24) | Placebo |

## Appendix 9: CINeMA Assessment

We use the CINeMA framework to evidence certainty, assessing it for each network estimate based on the following criteria:

**A: Within study bias:** We classified the overall risk of bias for each study as low risk of bias, the risk of bias as moderate when none of the four assessed risk of bias items were rated as high risk, and the risk of bias as high when one or both items were rated as high risk. See Appendix 4 for the bias assessment.

**Figure S9.1:**
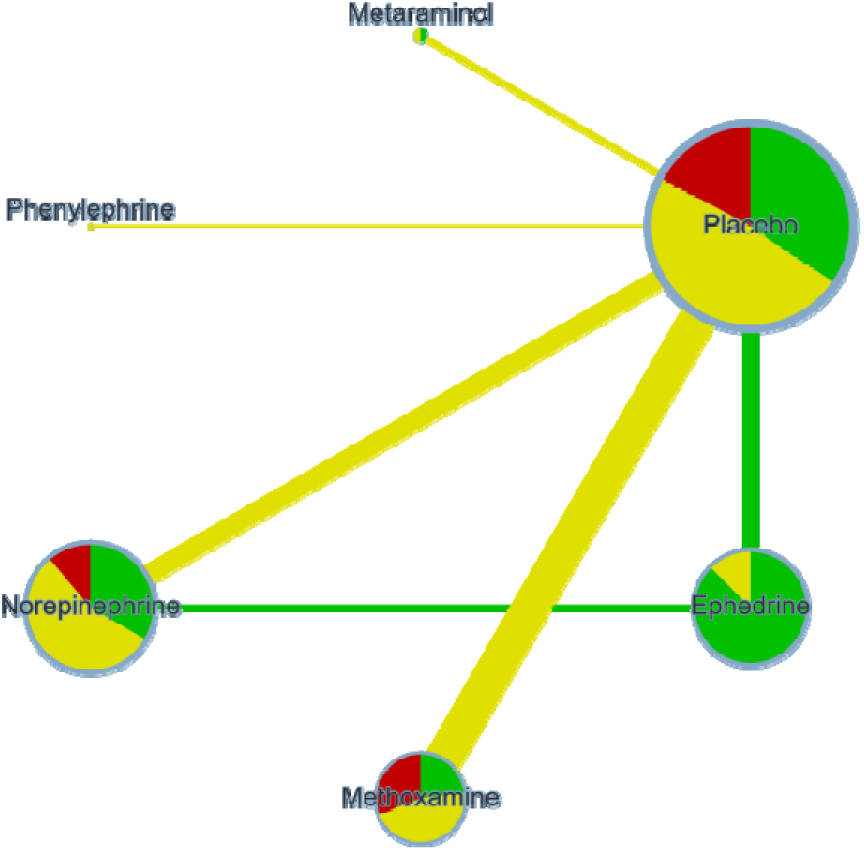
Risk of bias contribution by intervention group in Incidence of Hypotension

**Figure S9.2:**
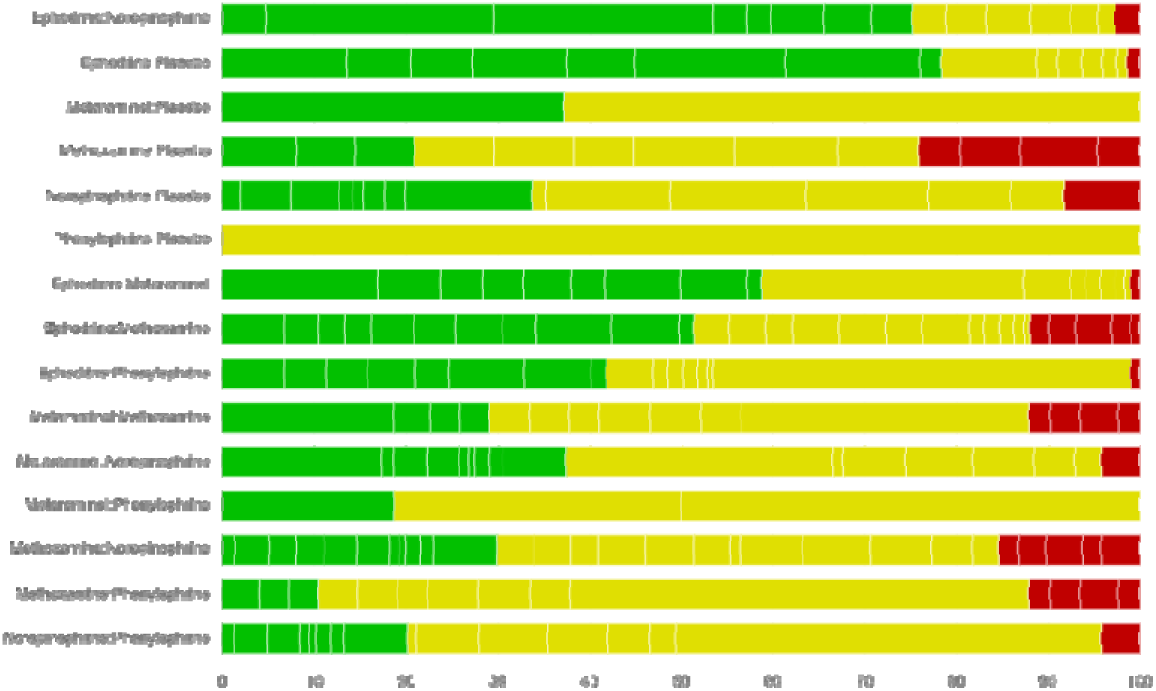
Overall risk of bias by treatment comparison in Incidence of Hypotension

**Figure S9.3:**
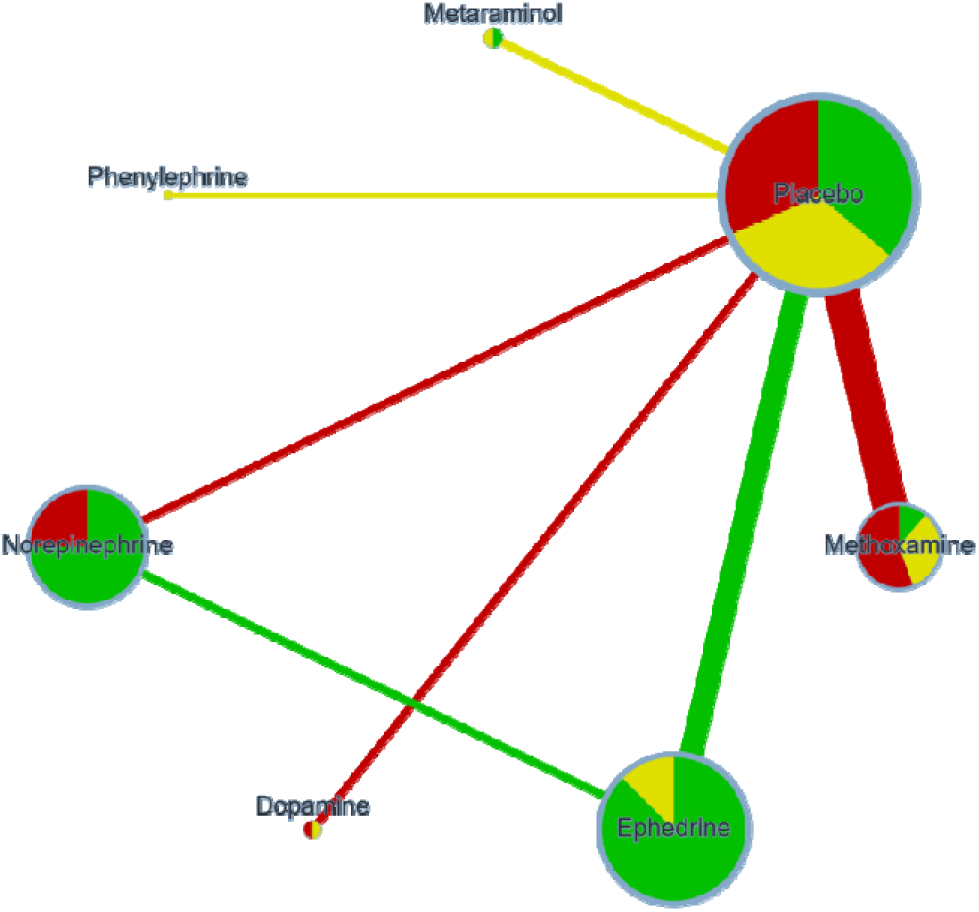
Risk of bias contribution by intervention group in Post-induction MAP

**Figure S9.4:**
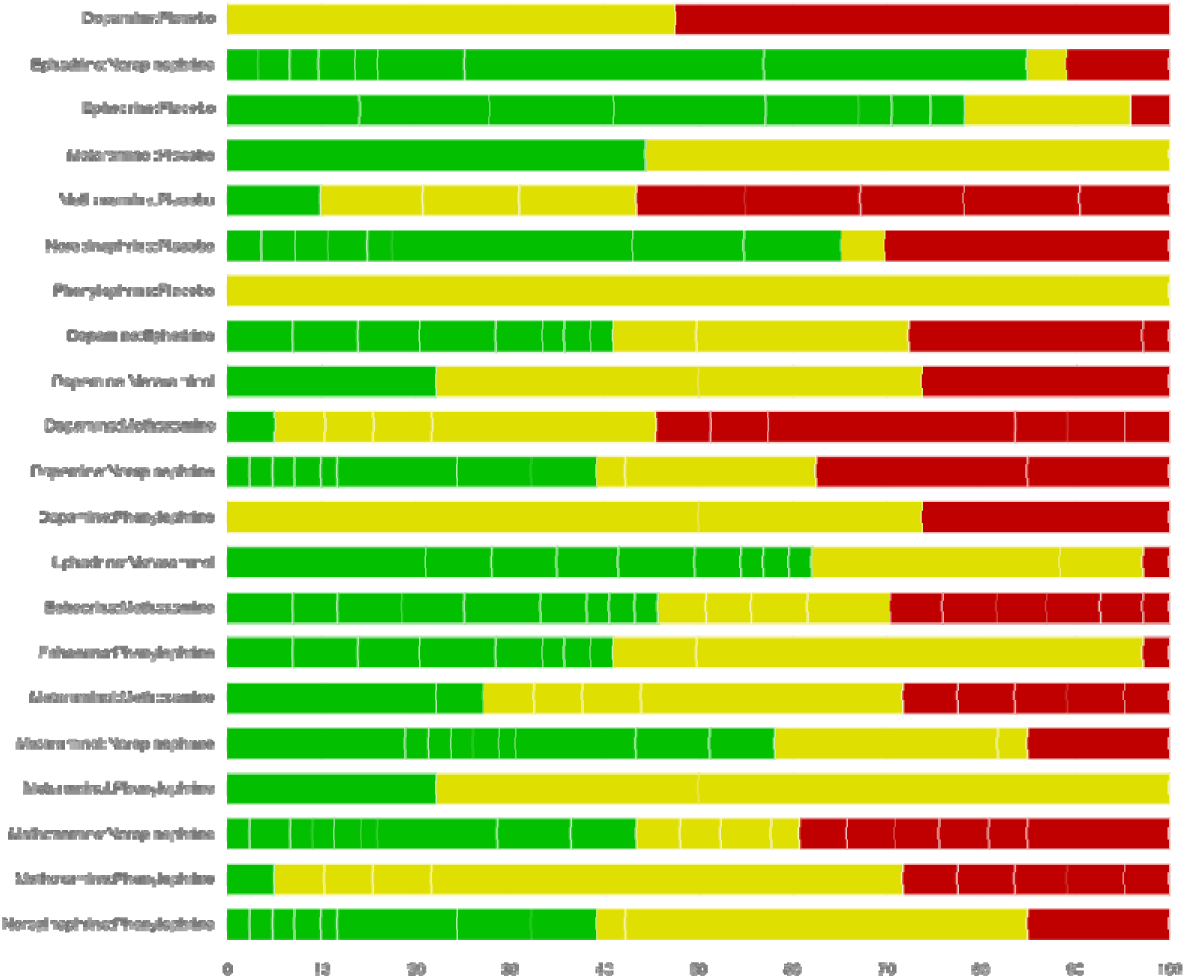
Overall risk of bias by treatment comparison in Post-induction MAP

**Figure S9.5:**
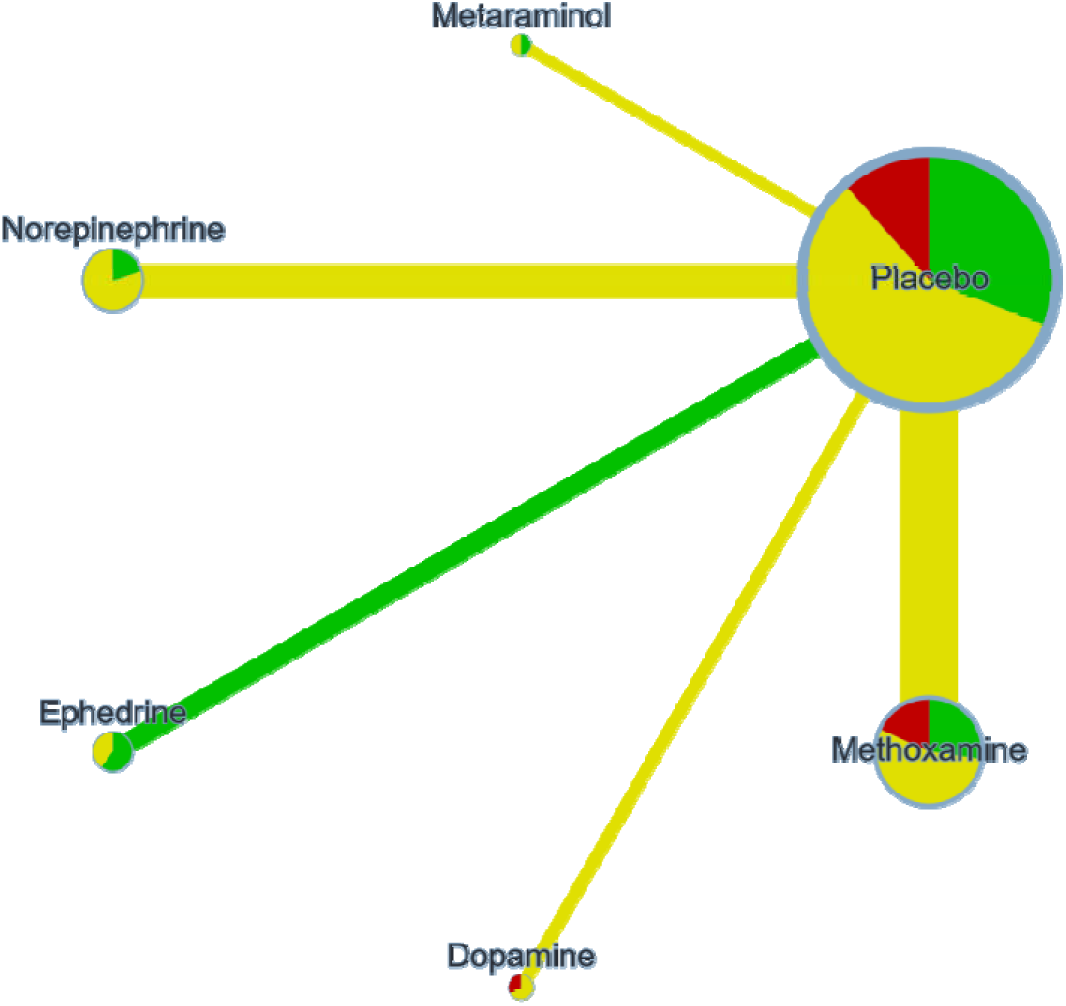
Risk of bias contribution by intervention group in Post-induction SBP

**Figure S9.6:**
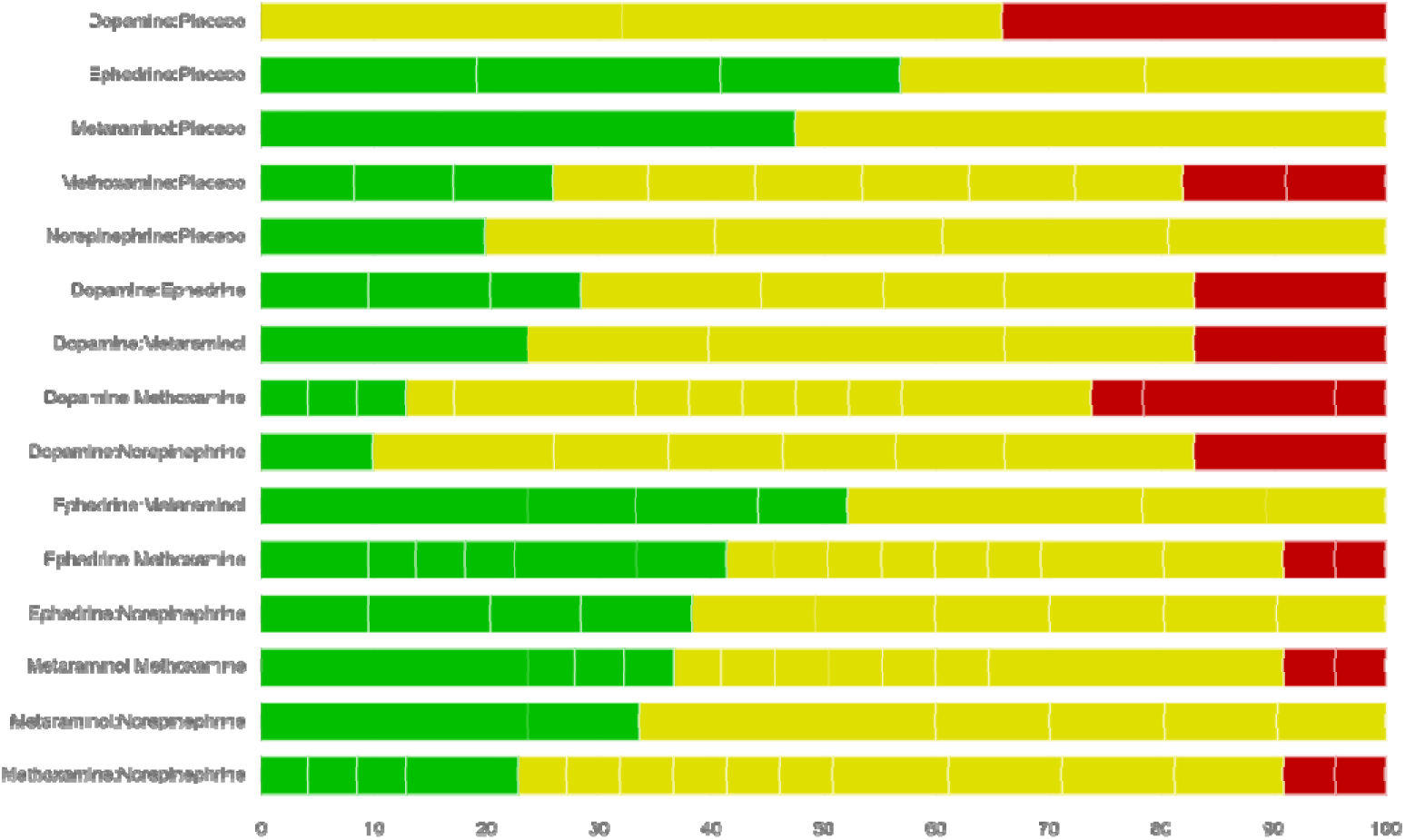
Overall risk of bias by treatment comparison in Post-induction SBP

**Figure S9.7:**
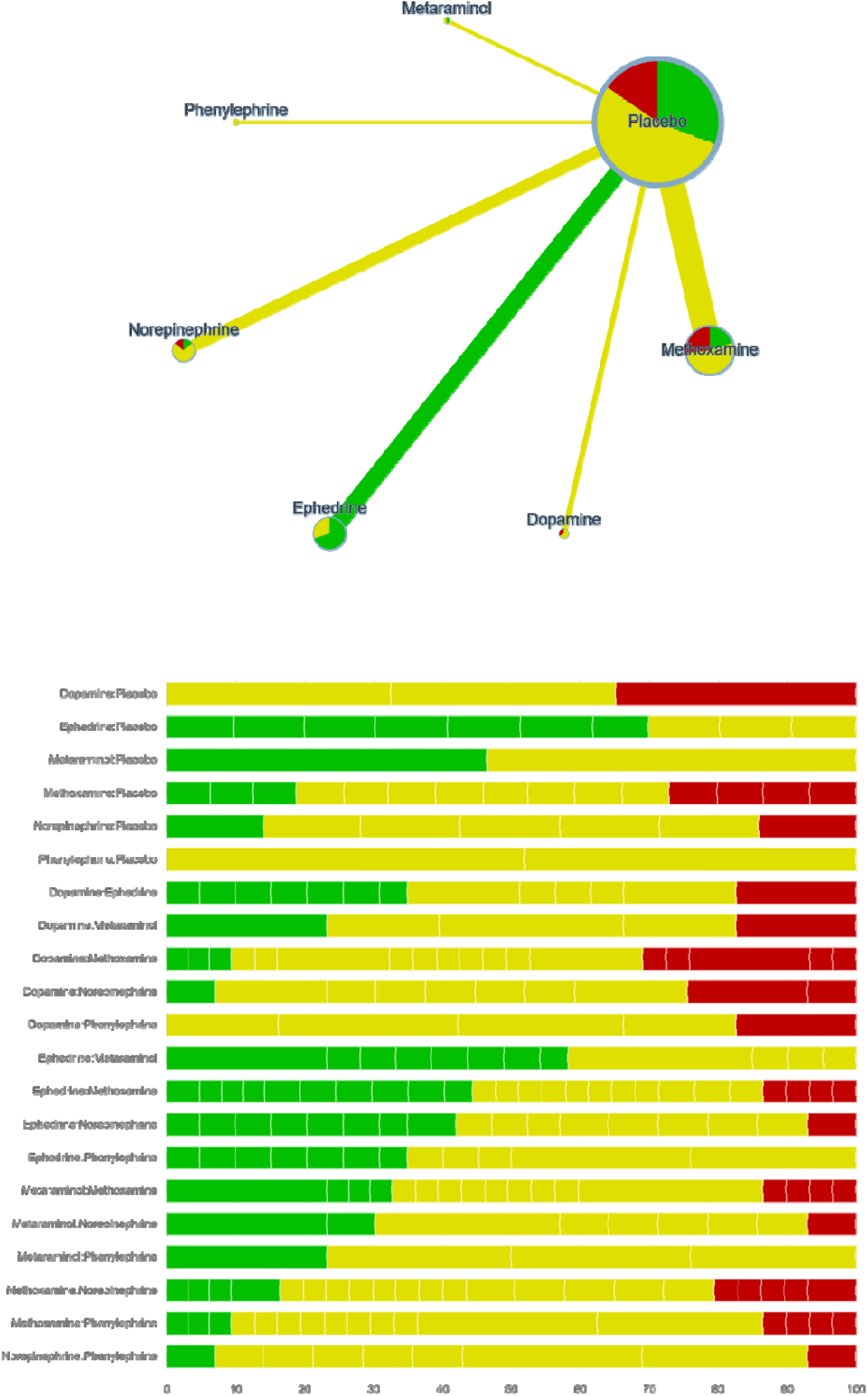
Risk of bias contribution by intervention group in Post-induction HR

**Figure S9.8:** Overall risk of bias by treatment comparison in Post-induction HR

